# Cardiovascular risk prediction in type 2 diabetes: a comparison of 22 risk scores in primary care setting

**DOI:** 10.1101/2020.10.08.20209015

**Authors:** K Dziopa, F W Asselbergs, J Gratton, N Chaturvedi, A F Schmidt

## Abstract

**Objective:** To compare performance of general and diabetes specific cardiovascular risk prediction scores in type 2 diabetes patients (T2DM).

**Design:** Cohort study.

**Setting:** Scores were identified through a systematic review and included irrespective of predicted outcome, or inclusion of T2DM patients. Performance was assessed using data from routine practice.

**Participants:** A contemporary representative sample of 203,172 UK T2DM patients (age ≥ 18 years).

**Main outcome measures:** Cardiovascular disease (CVD i.e., coronary heart disease and stroke) and CVD+ (including atrial fibrillation and heart failure).

**Results:** We identified 22 scores: 11 derived in the general population, 9 in only T2DM patients, and 2 that excluded T2DM patients. Over 10 years follow-up, 63,000 events occurred. The RECODE score, derived in people with T2DM, performed best for both CVD (c-statistic 0.731 (0.728,0.734), and CVD+ (0.732 (0.729,0.735)). Overall, neither derivation population, nor original predicted outcome influenced performance. Calibration slopes (1 indicates perfect calibration) ranged from 0.38 (95%CI 0.37;0.39) to 1.05 (95%CI 1.03;1.07). A simple, population specific recalibration process considerably improved performance, ranging between 0.98 and 1.03. Risk scores performed badly in people with pre-existing CVD (c-statistic ∼0.55). Scores with more predictors did not perform better: for CVD+ QRISK3 (19 variables) c-statistic 0.69 (95%CI 0.68;0.69), compared to CHD Basic (8 variables) 0.71 (95%CI 0.70; 0.71).

**Conclusions:** CVD risk prediction scores performed well in T2DM, irrespective of derivation population and of original predicted outcome. Scores performed poorly in patients with established CVD. Complex scores with multiple variables did not outperform simple scores. A simple population specific recalibration markedly improved score performance and is recommended for future use.

## Introduction

Despite major advances in treatment, people with type 2 diabetes (T2DM) remain at high risk for cardiovascular disease (CVD), the main cause of morbidity and mortality in this population^1^. There is however considerable heterogeneity in risk^2^, supporting the need for risk-stratified management.

CVD treatment initiation and intensification are guided by risk prediction algorithms. The UK National Institute for Health and Care Excellence (NICE) guidelines pragmatically recommends the use of the QRISK2 risk prediction tool in people with and without diabetes. The American College of Cardiology/American Heart Association (ACC/AHA) recommends estimating the 10-year risk of CVD using the Atherosclerotic Cardiovascular Disease (ASCVD) risk score^3^. Contrary to this, the European Society of Cardiology (ESC) does not recommend a CVD risk-prediction tool, and instead stratifies patients into three categories based on risk factors including: presence of target organ damage, number of risk factors, diabetes duration and age^4^. With over 300+ published CVD risk prediction tools^5^, many of which have not been validated in T2DM patients, nor directly compared within the same patient population, it is unclear which CVD scores performs best in T2DM. Previous comparisons only partially addressed this question, due to either focusing on non-representative T2DM patient enrolled in drug trails^6^, focused on a relatively short follow-up^7^, or used a very modest sample of T2DM patients ^8^, and often focusing on a small subset of available scores utilized in clinical practice, without exploring performance to predict CVD outcomes more relevant for T2DM patients. Quite apart from the greater CVD risk, even at a given level of individual risk factors, it is evident that the initial presentation of CVD in T2DM differs from that of the general population, with greater representation of heart failure and of peripheral artery disease (PAD), while hemorrhagic strokes are less frequent ^9^. General population scores, and indeed many designed for people with diabetes, have focused largely on prediction of coronary heart disease (CHD) and stroke only.

Our aim was to quantify the validity of existing general population and T2DM risk scores in predicting standard CVD (CHD, stroke, PAD), as well as a broader definition of major CVD outcomes that includes heart failure (HF) and atrial fibrillation (AF) as these are frequent outcomes in diabetic populations. We performed a systematic review to identify CVD risk prediction scores, and subsequently validated these in a large, UK-based electronic health records dataset. We also performed key subgroup analyses stratifying by gender, age, and CVD history and treatment.

## METHODS

### Systematic Review

A comprehensive literature search for CVD risk assessment tools was performed using MEDLINE, focusing on publications between 30 June 2008 to 16 January 2019; see search strategy in Appendix Figure 1 (*Systematic review - search strategy section*). Two reviewers (K. Dziopa, J. Gratton) independently reviewed the identified titles and abstracts, followed by full-text papers. Publications before this date were searched for using a previous review^10^.

Risk prediction models were included if they: (1) were derived from prospective cohort studies or randomised trials; (2) were derived in general (with or without exclusion of people with diabetes) or diabetic populations; (3) reported a measure of performance, and assessed 10-year risk of CVD, stroke, CHD, AF, HF or any combination of these (4) contained sufficient information to be run in the validation dataset; see Appendix Figure 1. Information was extracted on the derivation population, the statistical model (e.g., Cox, logistic regression, Weibull), year of publication, type of CVD, follow-up time and predictor definitions. For presentation purposes, rules were grouped on their derivation outcome: CVD, CHD, or other (including stroke and heart failure (HF)).

### Diabetes patient cohort

A cohort of 203,172 T2DM patients (18 years or older and without AF at the time of diabetes diagnosis) was extracted from CALIBER (Cardiovascular disease research using Linked Bespoke studies and Electronic health Records), linking three English EHR (Electronic Health Records) sources: primary care records from the Clinical Practice Research Datalink (CPRD), Hospital Episodes Statistics (HES) and national death registration from the Office for National Statistics (ONS)^11^.

T2DM patients in this dataset were identified based on a CALIBER phenotyping algorithm (https://www.caliberresearch.org/portal/phenotypes): this uses a combination of a GP (general practitioner) diagnosis of T2DM or ICD10 (International Statistical Classification of Diseases and Related Health Problems) / Read code for T2DM, full definition of components is provided in Appendix Table 6.

### Cardiovascular outcomes

Patients were followed-up from their initial T2DM diagnosis until their first cardiovascular event, death, end of study (2018-02-05), or 10-year follow-up landmark; whichever occurred first. Subjects with a previous record of AF were excluded, due to the inability to differentiate between ongoing versus recurrent AF events in EHR. Subjects with any other preexisting CVD event were included and history of CVD was used as a subgroup indicator.

A CVD event was defined as the first occurrence of fatal or non-fatal myocardial infarction (MI), sudden cardiac death, ischemic heart disease, fatal or non-fatal stroke or PAD since diagnosis of T2DM. We additionally defined CVD as including heart failure (HF) and / or atrial fibrillation (AF): ‘CVD+AF+HF’. Stroke consisted of any kind of fatal or non-fatal stroke. Detailed endpoint definitions from the CALIBER research portal^11^ are provided in Appendix Table 5.

### Patient characteristics

The following patient characteristics and measurements were extracted (see Appendix Table 9): gender, age (years), smoking status, glycated hemoglobin (HbA_1c_), fasting plasma glucose (FPG), body mass index (BMI), HDL and LDL cholesterol, total cholesterol, triglycerides, systolic blood pressure (SBP) and diastolic blood pressure (DBP), urine albumin to creatinine ratio, serum creatinine, C reactive protein, total white blood cell count, and electrocardiogram (ECG) results. Baseline predictor values were defined as measurements recorded closest to baseline (T2DM diagnosis date) and no more than 1 year prior or 1 week after the date of diagnosis of diabetes. The impact of a more liberal time-windows is described in Appendix Table 10. Any predictor without a measurement within this time frame was defined as missing. Information was also collected on the presence or absence of: rheumatoid arthritis, renal disease, foot amputation, systemic lupus erythematosus, Human Immunodeficiency Virus (HIV) infection, mental disorders, microalbuminuria, erectile dysfunction, hypertension, and migraine at baseline. Prescriptions of the following drugs were extracted: anticoagulants, diuretics, corticosteroids, statins, and blood pressure lowering medication. Finally, information on social deprivation (Townsend score) and family history of CVD, CHD, MI and stroke were sourced.

### Statistical analysis

Models were evaluated on discrimination (using Harrell c-statistic^12^), calibration (calibration-in-the-large and calibration slope^13^); see appendix (page 18) for a brief description of these metrics. We note that for binary outcomes predicted at a single moment in time the c-statistic is identical to the area under a receiver operator characteristic (ROC) curve ^12^. These models were evaluated both before, and after model recalibration, where a model’s intercept and slope is updated to adapt a risk score to a different populations, a similar but distinct outcome, or both. Here the available risk scores were independently recalibrated to predict all six of the CVD endpoints described. To prevent model overfitting, recalibration was performed in a 10% (20,317) independent training sample, which is an ample sample size to estimate the two coefficients (the intercept and slope) necessary for model recalibration. The remaining 90% (182,855) of the dataset was used to compare like-with-like model performance of the un-calibrated and recalibrated models.

Missing variables (presented in Table 1 and Appendix Table 7 - 9) were imputed using multiple imputation^14^. Imputation variables were selected using the procedure described in^15^, guarding against imputation while at the same time maximizing predictive accuracy. Moreover, the procedure eliminates predictors whose proportion of usable cases fails to meet a minimum value (here 0.5). Imputation specific results were combined using Rubin’s rules^16^.

**Table 1.**
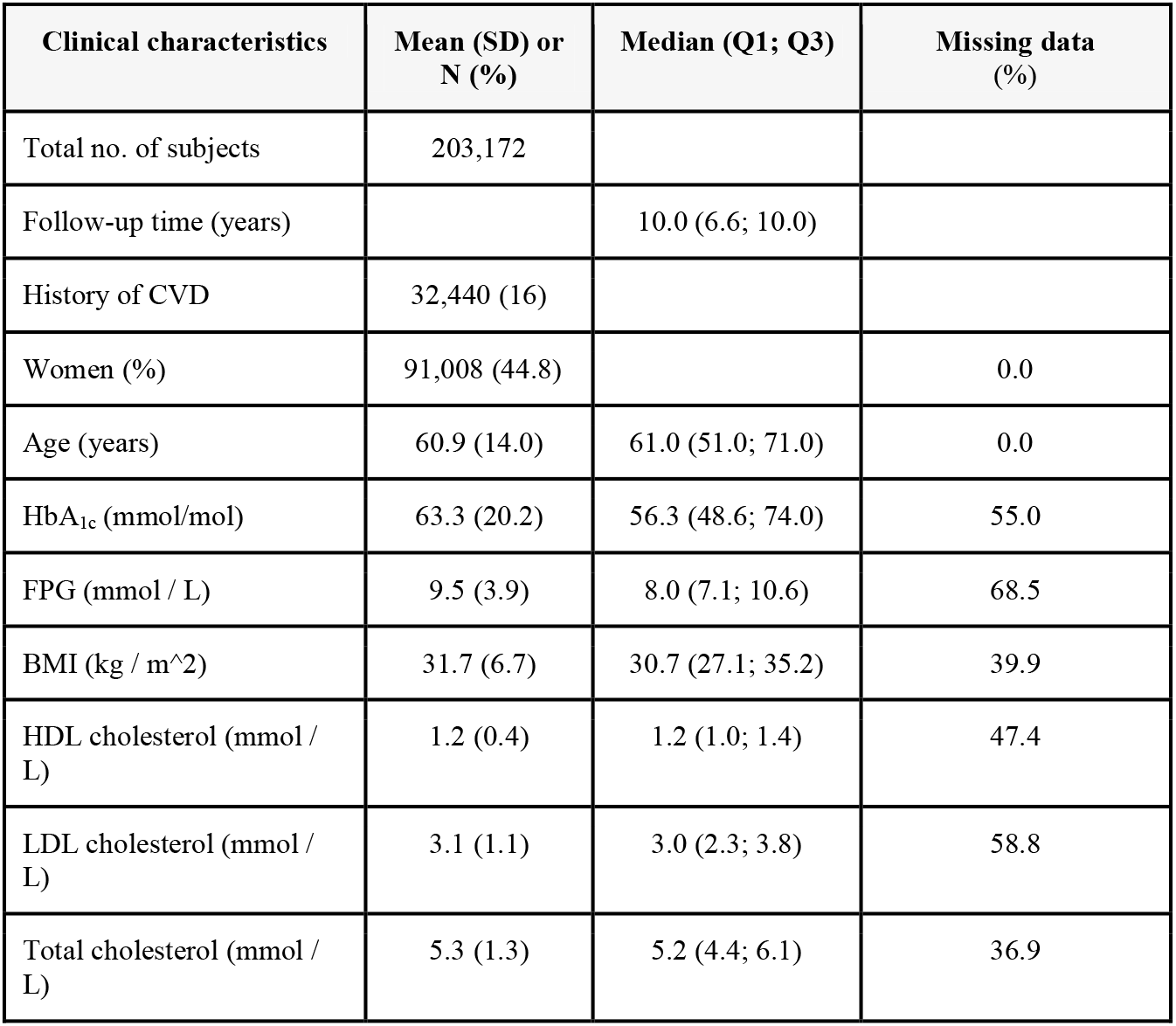
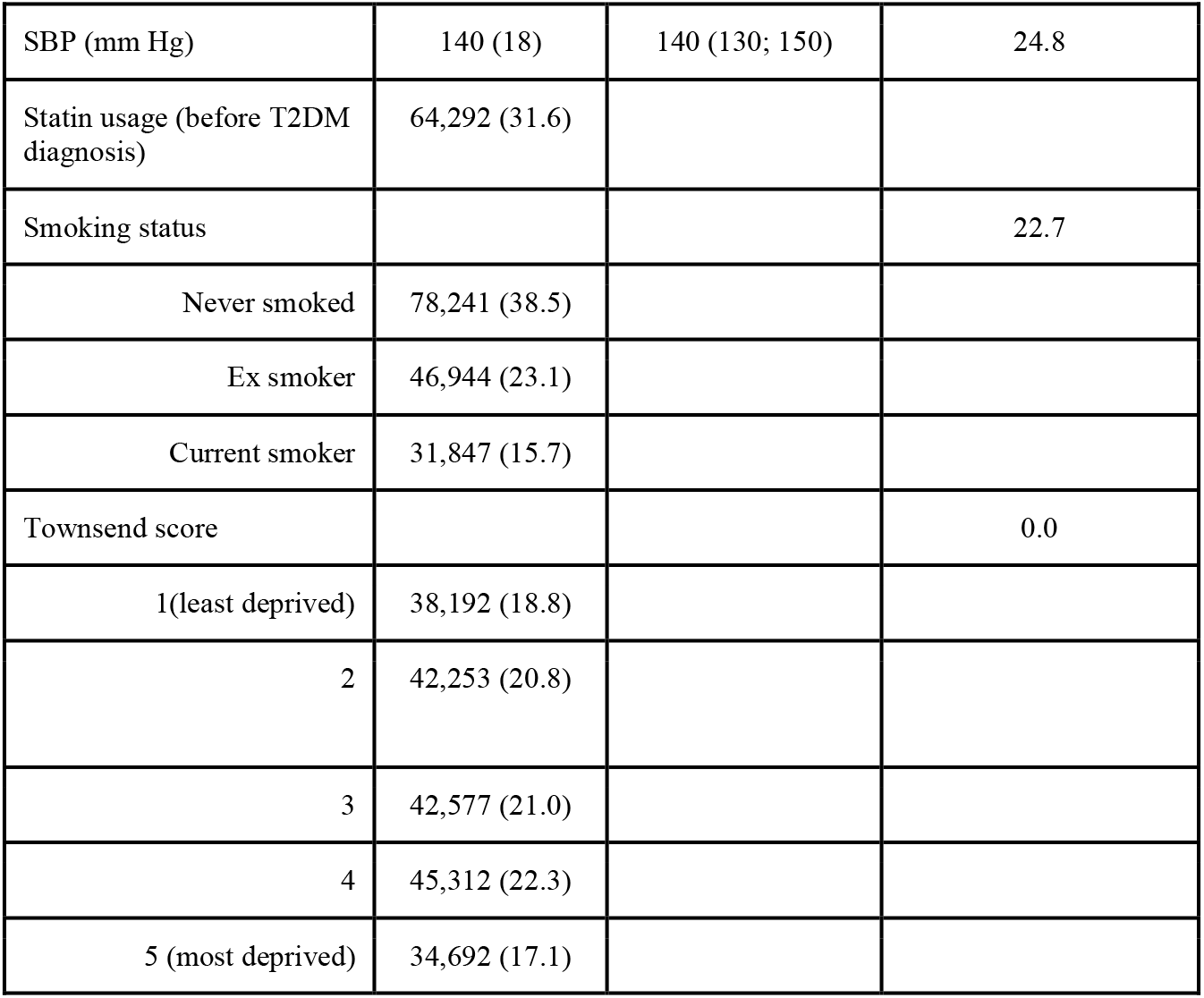
Clinical characteristics of prediction score variables around the time (1 year before - 1 week after) of diagnosis of T2DM.

| Clinical characteristics | Mean (SD) or N (%) | Median (Q1; Q3) | Missing data (%) |
| --- | --- | --- | --- |
| Total no. of subjects | 203,172 |  |  |
| Follow-up time (years) |  | 10.0 (6.6; 10.0) |  |
| History of CVD | 32,440 (16) |  |  |
| Women (%) | 91,008 (44.8) |  | 0.0 |
| Age (years) | 60.9 (14.0) | 61.0 (51.0; 71.0) | 0.0 |
| HbA<sub>1c</sub> (mmol/mol) | 63.3 (20.2) | 56.3 (48.6; 74.0) | 55.0 |
| FPG (mmol / L) | 9.5 (3.9) | 8.0 (7.1; 10.6) | 68.5 |
| BMI (kg / m<sup>2</sup>) | 31.7 (6.7) | 30.7 (27.1; 35.2) | 39.9 |
| HDL cholesterol (mmol / L) | 1.2 (0.4) | 1.2 (1.0; 1.4) | 47.4 |
| LDL cholesterol (mmol / L) | 3.1 (1.1) | 3.0 (2.3; 3.8) | 58.8 |
| Total cholesterol (mmol / L) | 5.3 (1.3) | 5.2 (4.4; 6.1) | 36.9 |
| SBP (mm Hg) | 140 (18) | 140 (130; 150) | 24.8 |
| Statin usage (before T2DM diagnosis) | 64,292 (31.6) |  |  |
| Smoking status |  |  | 22.7 |
| Never smoked | 78,241 (38.5) |  |  |
| Ex smoker | 46,944 (23.1) |  |  |
| Current smoker | 31,847 (15.7) |  |  |
| Townsend score |  |  | 0.0 |
| 1 (least deprived) | 38,192 (18.8) |  |  |
| 2 | 42,253 (20.8) |  |  |
| 3 | 42,577 (21.0) |  |  |
| 4 | 45,312 (22.3) |  |  |
| 5 (most deprived) | 34,692 (17.1) |  |  |

The above described analyses were performed on the overall sample, and on subgroups stratified on CVD history at T2DM diagnosis (absent vs present), gender (male vs female), age (four similarly sized categories) and statins usage at T2DM diagnosis (statin naïve vs statin user). These subgroups were specifically selected based on prior knowledge: among T2DM patients CVD risk increases more in women than in men; CVD risk increases with age, statin therapy reduces risk of CVD in adults at increased CVD risk without prior CVD events; preexisting CVD increases risk of recurrent events.

Discrimination was assessed using the c-statistic, calibration using calibration-in-the-large (CIL) and calibration slope (CS), and by calculating 95% confidence intervals (95%CI). To guard against over-optimism all estimates were calculated using the test data, independent form the training data used for potential model recalibration. All analyses were carried out in R, version 3.6.1. Calibration plots were generated using the *ggplot2* package^17^, statistics calculated using *Hmisc*^18^, and forest plots using the *metafor* package^19^.

## RESULTS

The systematic review retrieved 1,171 potentially relevant articles, of which 42 were retained after title and abstract screening. The majority were excluded due to poor or no validation and lacking performance metrics. After screening the full-text, we excluded 14 publications due to short follow-up time (less than 10 years), 4 used unavailable predictors in the CALIBER database, 2 did not provide enough details to implement, 2 did not report internal validation results, 3 were point risk scores, 1 was not published in English, and 1 did not present a new risk score (Appendix Figure 18). Finally, we included 15 publications reporting 22 different risk score models that reported 10 year risk of developing any kind of CVD with sufficient information to be run in the CALIBER database. Only two of the included scores were published before 2000 (Framingham 1991^20^, Framingham 1998^21^); Appendix Table 3.

Out of 22 identified CVD risk prediction models, 8 were derived in T2DM subjects alone (DARTS^22^, UKPDS 56^23^, UKPDS 68 C-HF and Stroke^24^, UKPDS 82 C-HF and CHD^25^, CHS Basic and Advanced^26^), 2 excluded T2DM subjects (SCORE CHD and CVD^27^), and 12 scores enrolled both non-T2DM as well as T2DM patients (Finrisk Stroke, CHD and CVD^28^, Framingham 1991 fatal CHD, CVD and Stroke^20^, Framingham 1998^21^, QRISK 2 ^29^, QRISK 3 ^30^, ASCVD ^3^, RECODE ^31^, and Reynolds Risk ^32 33^). Ten rules were designed to predict CVD, 7 CHD, 3 stroke, and 2 HF, see Appendix Table 4.

All of the risk scores incorporated classic CVD risk factors, such as age, sex, blood pressure and smoking status. Twenty risk scores included information about lipids. The scores that included a proportion of T2DM patients typically included T2DM (presence/absence) as a predictor, but did not include diabetes-specific risk factors such as diabetes duration, and glycaemic status (which were often used in T2DM specific scores). The total number of predictors taken into account for different risk prediction models ranged from 6 (SCORE ^27^) to about 19 (QRISK 3 ^30^); see Appendix Figure 2, and Appendix Table 4.

Baseline characteristics of included patients are presented in Table 1 and Appendix Table 7 – 9. Average age was around 60 years (SD: 14.0), 9,108 (45%) of participants were women. Just under a fifth (32,440 (16%)) had a previous history of CVD, and 64,292 (32%) were on statins.

### Number of and timing of CVD events

During a median follow up of 10.0 years (see Appendix Table 10), 63,000 (31.07%) T2DM patients suffered CVD, AF, or HF events, of these 51,636 (25.46%) had a CVD event, 40,242 (19.84%) CHD, 20,506 (10.11%) AF, (16,993;8.38%) HF and (10,413; 5.14%) stroke (see Figure 1 for Kaplan-Meier estimates).

**Figure 1.**
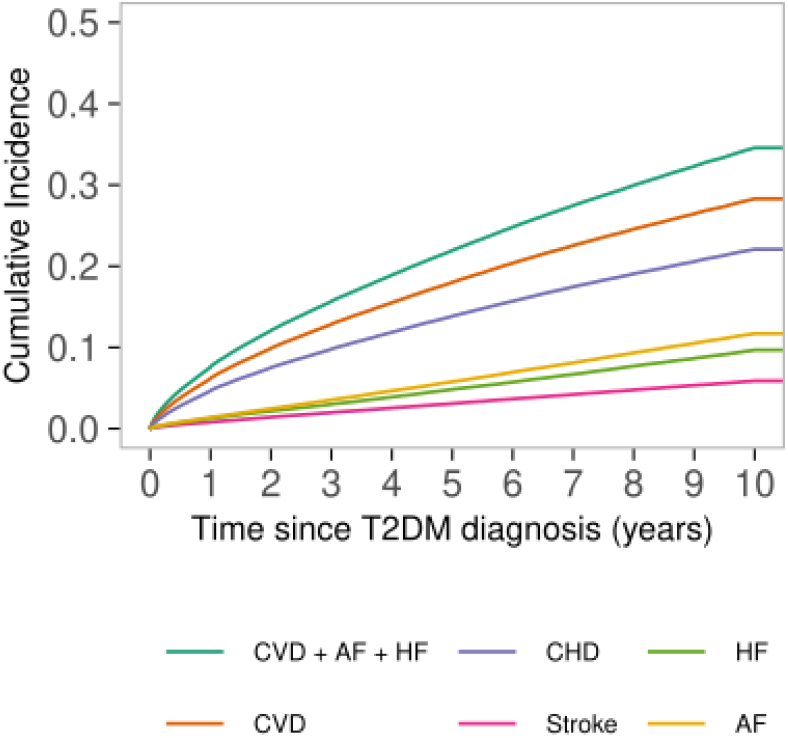
Kaplan-Meier estimates of the 10-years cumulative incidence of CVD after a T2DM diagnosis. N.b. follow-up time was censored if a subject did not experience a CVD event during the 10-year follow-up time. Cumulative incidence rate (CIR) within the 10 years time frame was the highest for the combination of the outcomes including CVD, AF and HF.

### Predicting cardiovascular risk in T2DM patients

We found little difference between analyses using complete-case data (Appendix Figures 4-5, and Appendix Tables 12-13) and multiple imputation and hence present the later in the main text.

Most models could accurately predict CVD (CS: from 0.38 to 1.05, CIL from −0.17 to 2.76) (Figure 2, and Appendix Table 14), even models designed to predict stroke and/or HF did not underperform substantially compared to CVD derived models. The scores almost uniformly under-estimated the risk of CVD+AF+HF, the exception being the Framingham 1991 CVD score, which systematically overestimated risk.

**Figure 2.**
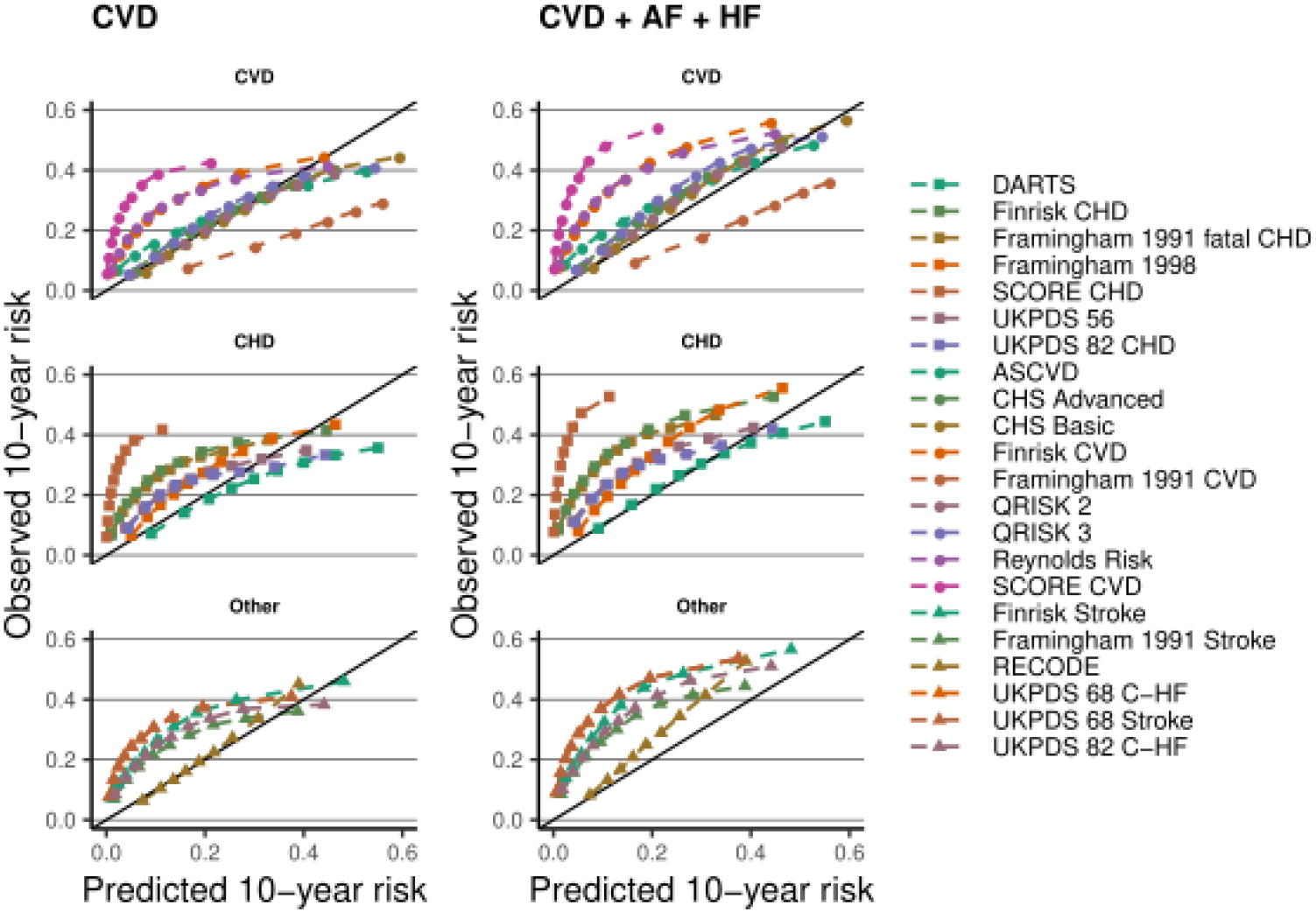
Calibration plots of 22 prediction rules for 10-years CVD risk, applied toT2DM patients. n.b. Estimates based on imputed data. Depicted performance is based on 90% of the data used for external validation. The observed 10-years risk is (y-axes) plotted against the average predicted 10-year risk (x-axis) within groups defined by quintiles of predicted risk. The columns indicate the type of CVD the scores were evaluated against. Scores were grouped by the derivation outcomes CVD, CHD, or other (including Stroke, C-HF)). The diagonal line reflects perfect calibration.

The CHD Basic (CS: 0.80 CIL: −0.17), ASCVD (CS: 0.41 CIL: −0.15), and QRISK2 (CS: 0.67 CIL: −0.17) models (originally derived to predict *any* CVD) generally showed near perfect calibration, for both CVD, and CVD+AF+HF. Focusing on scores not originally intended to predict CVD, we found that the DARTS score (a CHD score) could accurately predict both CVD (CS: 0.50 (95%CI 0.48; 0.51), CIL: −0.53 (95%CI −0.55; −0.52)), and CVD+AF+HF (CS: 0.59 (95%CI 0.57; 0.60), CIL: −0.21 (95%CI - 0.22; −0.20)), for the “other group” (including stroke and HF derived scores) we found RECODE (CS: 1.05 (95%CI 1.03; 1.07), CIL: 0.08 (95%CI 0.07; 0.09)) for CVD and (CS: 1.10 (95%CI 1.08; 1.12), CIL: 0.39 (95%CI 0.38; 0.40)) for CVD+AF+HF performed well (Figure 2). Despite observing reasonable external calibration, models had more difficulty discriminating between subjects who experienced an event within 10-years and those who remained event free: the c-statistic was typically around 0.68. The Framingham 1991 CVD risk score and UKPDS risk scores were amongst the worst performers (Figure 3). With a c-statistic of 0.73 (95%CI 0.73; 0.73) the RECODE rule outperformed the others (interaction p-values < 0.001). Similar, patterns of discrimination were observed when attempting to predict CVD+AF+HF, with the latter combined endpoint showing a slightly improved c-statistics (closer to 0.70). The discriminatory performance of these 22 rules in classifying CHD, stroke, AF, and HF is presented in Appendix Figures 14-15.

**Figure 3.**
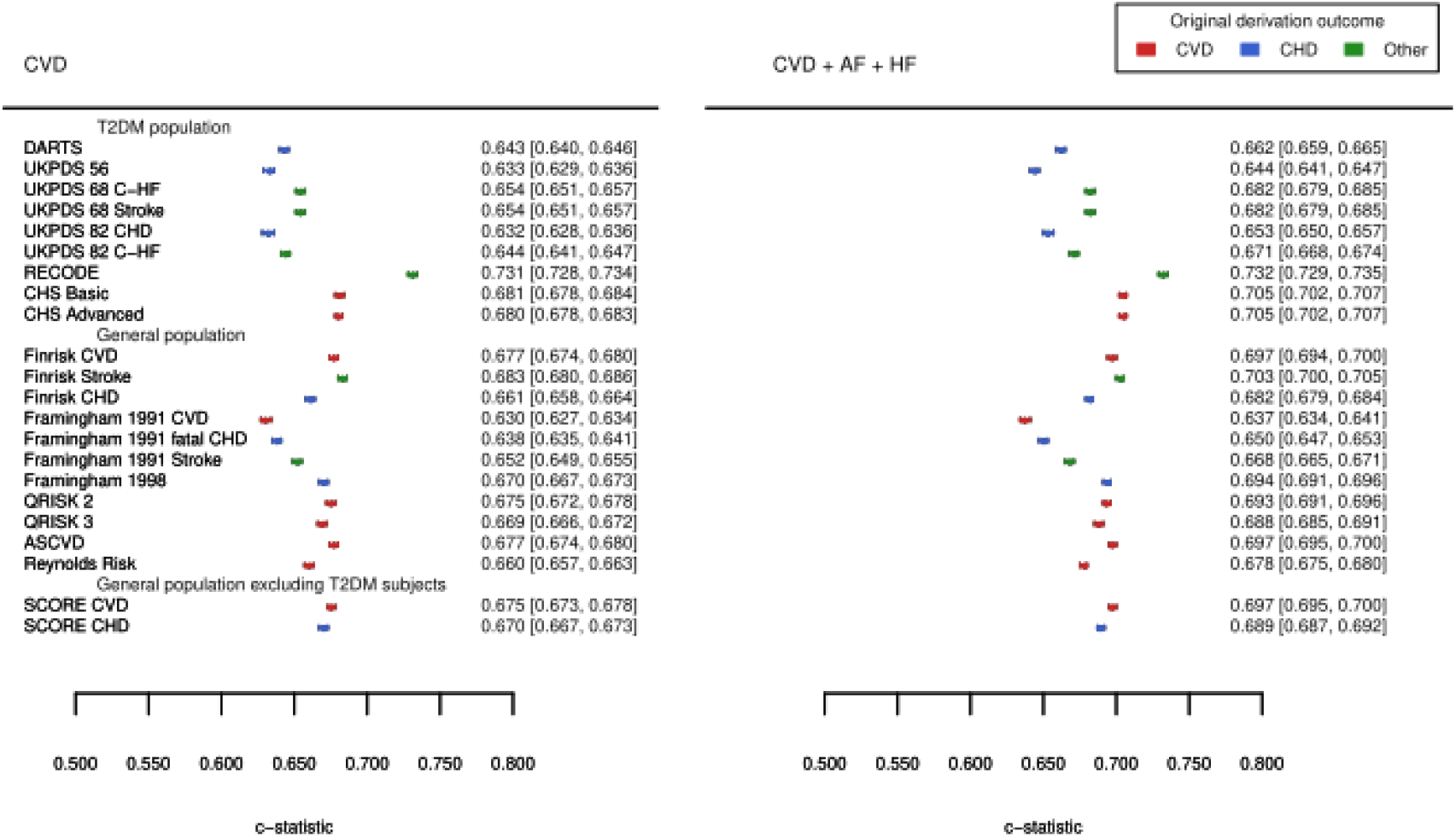
C-statistics (discrimination) of 22 CVD risk prediction tools externally validated in a UK-based T2DM sample split by the derivation population and the reported type of CVD outcome. n.b. Point estimates are presented alongside 95%CI. Results were based on imputed data and based on 90% of the data used for external validation.

We observed that scores with a large number of predictors did not necessarily outperform scores with fewer variables: QRISK3 (19 variables) CVD+AF+HF c-statistic 0.69 (95%CI 0.68;0.69), compared to 0.70 (95%CI 0.70; 0.70) for ASCVD (9 variables) and 0.71 (95%CI 0.70; 0.71) for CHD Basic (8 variables); with similar results for the CVD only outcome. Similarly, while the in T2DM patient derived RECODE did outperform the remaining 21 scores, but a restriction of a derivation population only to T2DM patients did not seem to generally improve discrimination (Figure 3).

### Performance after recalibration

Recalibrating the 22 models in the 10% training dataset considerably improved performance (Appendix Figure 8-11, and Appendix Tables 13, 15), with most rules showing near perfect calibration in the remaining 90% of the data used for model evaluation (the test set). Given that most of these 22 rules were not designed to predict stroke, AF, or HF it was somewhat surprising to see that recalibration markedly improved performance for these endpoints as well, with Figure 4 showing near perfect agreement between predicted and observed risk. For example, after recalibration, QRISK3 could predict HF (CS: 0.95 95%CI 0.87; 1.04) (Figure 6) and AF (CS: 0.97 95%CI 0.91; 1.04) remarkably well.

**Figure 4.**
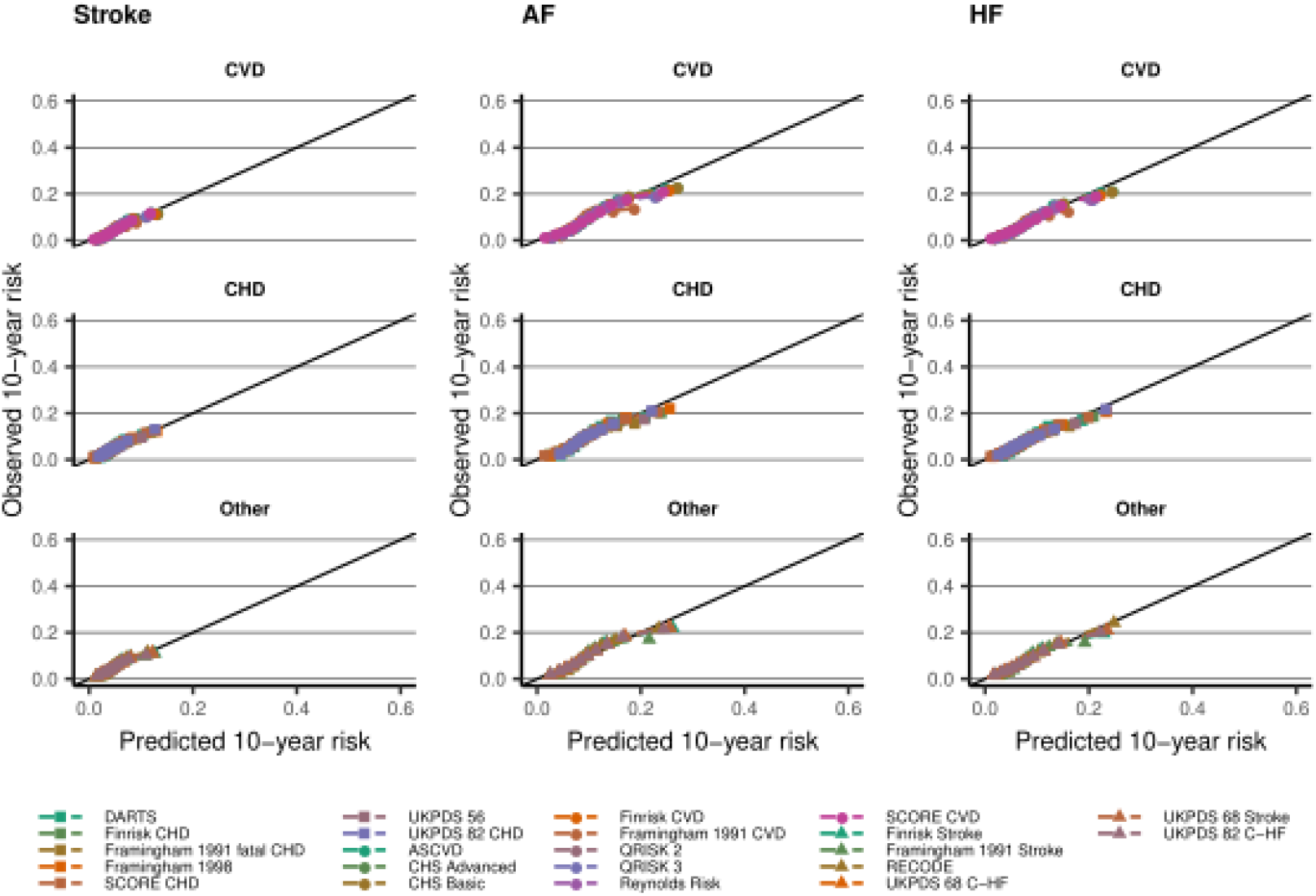
Calibration plots after recalibrating 22 prediction rules for 10-years CVD risk, applied toT2DM patients. n.b. Estimates based on imputed data. Depicted performance is based on 90% of the data used for external validation, independent of the 10 hold-out sample used to recalibrate the models. The observed 10-years risk is (y-axes) plotted against the average predicted 10-year risk (x-axis) within groups defined by quintiles of predicted risk. The columns indicate the type of CVD the scores were evaluated against. Scores were grouped by the derivation outcomes CVD, CHD, or other (including Stroke, C-HF)). The diagonal line reflects perfect calibration.

**Figure 5.**
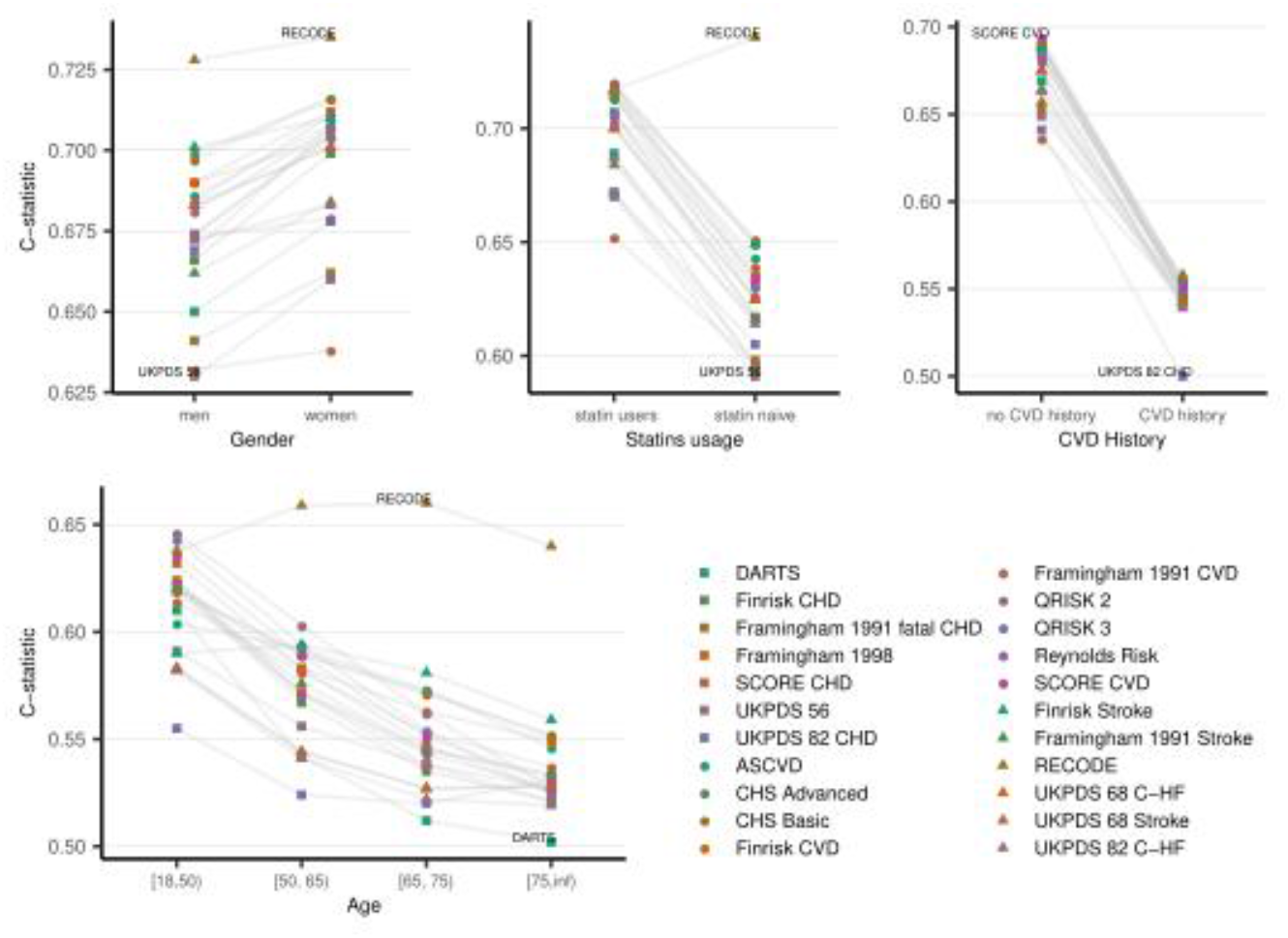
Discrimination (c-statistic) for all included risk scores against CVD + AF + HF outcome for the imputed dataset stratified by gender (men vs women), statins (statin naïve vs statin users), CVD history at the baseline (absent vs present), and age subgroups. n.b. Results were based on imputed data and based on 90% of the data used for external validation. Point estimates and 95%CI are presented in Appendix Table 16 – 19

**Figure 6.**
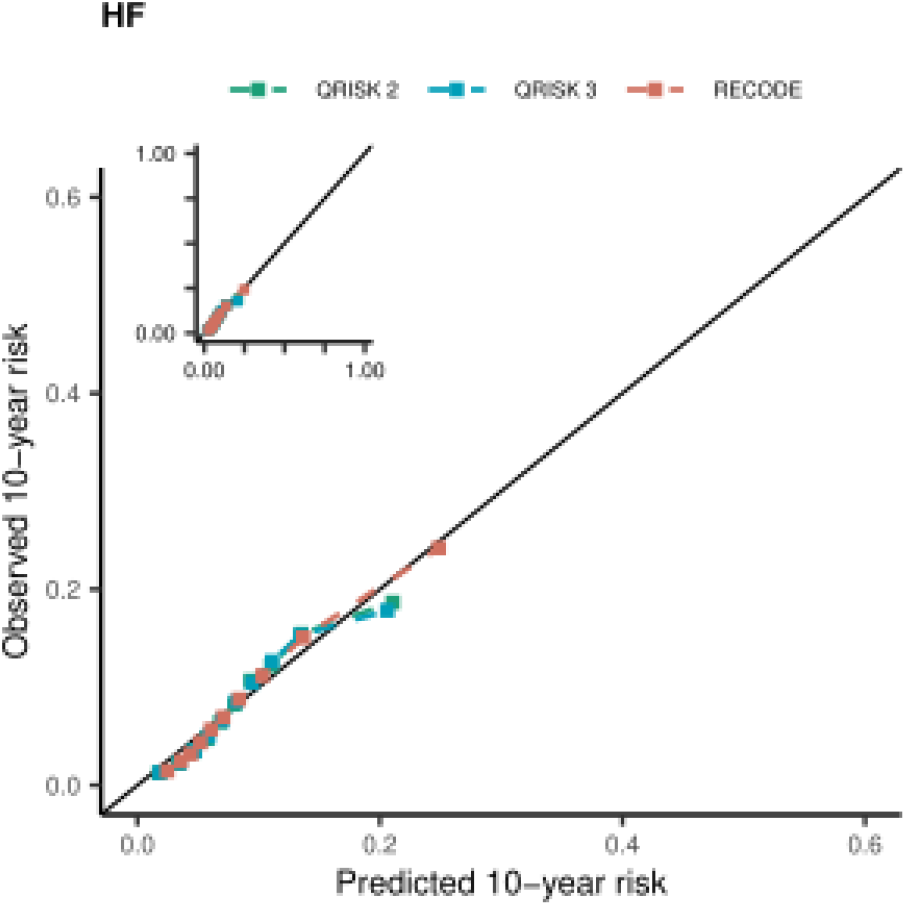
Calibration plots after recalibrating 3 prediction rules (QRISK2, QRISK3, RECODE) for 10-years HF risk, applied to T2DM patients. n.b. Estimates based on imputed data. Depicted performance is based on 90% of the data used for external validation, independent of the 10 hold-out sample used to recalibrate the models. The observed 10-years risk is (y-axes) plotted against the average predicted 10-year risk (x-axis) within groups defined by quintiles of predicted risk. The scores were evaluated against HF outcome.

### Subgroup analyses

The discriminative ability of the scores decreased with age (Figure 5, Appendix Figure 16), with some even failing to discriminate at all in older age groups: for example the UKPDS 82 C-HF c-statistic was 0.50 (95%CI 0.49; 0.51) for CVD (see Appendix Table 17). It was generally more difficult to accurately identify CVD, or CVD+AF+HF in men than in women (Figure 5), however the RECODE performed similarly by sex: c-statistic 0.73 (95%CI 0.72; 0.73) in men for both CVD and CVD+AF+HF, compared to 0.73 (95%CI 0.73;0.74) in women for CVD, and 0.73 (05%CI 0.73;0.74) in women for CVD+AF+HF; interaction p-values: 0.98 for both endpoints. Performance was markedly poorer in statin naïve versus statin user groups, again with the exception of RECODE which performed similarly in both groups (Figure 5). Performance was markedly poorer in those with established CVD versus those without CVD (Figure 5). The best performing score for patients without CVD at T2DM diagnosis was the SCORE CVD rule: c-statistic of 0.67 (95%CI 0.67;0.67) for 10-year CVD risk; and 0.69 (95%CI 0.69;0.70) for 10-year CVD+AF+HF risk; ASCVD, Finrisk CVD, and SCORE CHD achieved similar results. The c-statistic for people with established CVD was around 0.5 for all of the risk scores; the highest c-statistic of 0.54 (95%CI 0.53;0.55) for 10-year CVD risk was from the UKPDS 82 CHD, and 0.56 (95%CI 0.55;0.57) for CVD+AF+HF obtained from RECODE (Appendix Table 19).

## DISCUSSION AND CONCLUSION

We identified, and subsequently validated 22 cardiovascular risk prediction tools for a range of macrovascular endpoints in an English primary care cohort of 203,172 people with T2DM. We report a number of unique findings. Firstly, the UK recommended QRISK2 score performed comparatively well for CVD (c-statistic of 0.68 95%CI 0.67;0.68), and CVD combined with HF and AF (c-statistic of 0.69 95%CI 0.69;0.70), and for the individual endpoints AF, HF, CHD and any stroke. Secondly, diabetes specific scores do not appear superior to scores derived for the general population. Thirdly, scores performed universally poorly for T2DM patients with established CVD (c-statistics close to 0.50; random classification). Fourthly, scores with many additional features did not outperform those with fewer, and more readily available (in primary care) predictors. Finally, a simple recalibration step can markedly improve score performance, repurposing scores intended to predict *any* CVD or CHD to accurately predict stroke, AF and HF risk (see Figure 4).

We externally evaluated two widely used risk prediction scores in the UK (QRISK2 and QRISK3), with a good discriminatory ability in the general population: with c-statistics for QRISK2 of 0.82 in women, and 0.79 in men, and for QRIKS3 0.88 in women, and 0.86 in men. With a c-statistic below 0.70, we show that performance of both scores was markedly decreased in T2DM patients. This poor performance is surprising given that the QRISK scores were derived in a similar, but independent, sample of English patients, and used the same electronic healthcare infrastructure. Despite this, QRISK2 and QRISK3 did not outperform non-UK based scores such as RECODE, or CHS, and we note the RECODEs overall superior discriminative ability. The difference in discrimination between RECODE (0.73 95%CI 0.73; 0.74 for CVD+AF+HF) and QRISK2 (0.69 95%CI 0.69; 0.70) and QRISK3 (0.69 95%CI 0.69; 0.69) was statistically significant (interaction p-value < 0.001 for CVD and CVD+AF+HF), depending on the public health and policy implications, this modest increase might be sufficient to be considered as an alternative to the widely used QRISK rules in the UK or in other countries.

Predictive performance of the risk scores was markedly poorer in patients with pre-existing CVD at the time of T2DM diagnosis (c-statistic around 0.5). Most of the prediction models were developed in subjects without clinical manifestations of CVD, and were not validated in people with established CVD. Moreover, these risk scores lack predictors that are of particular importance in patients with established disease, such as time since first diagnosis of CVD, history of CVD and renal function^34^. Thus a risk score in a T2DM population with prevalent CVD needs to be developed. Despite recalibrating the scores in an independent training sample, calibration was still far from perfect in those with established CVD (Appendix Table 19), further underlining the scope for improvement in this patient subgroup.

The necessity for a T2DM specific score has often been made^7^ and revolves around the need to account for excess risk unexplained by conventional risk factors, and the desire to include diabetes specific variables such as HbA1c and diabetes duration, which are known observationally to predict CVD risk ^35^. It is suggested that a T2DM specific score can better deal with exposure and outcome associations specific to T2DM patients^22^. Despite these argument, we did not observe a clear benefit of T2DM specific rules (including T2DM specific variables) compared to scores derived in samples with a mixture of T2DM patients and the general population, or even actively excluding T2DM patients. This suggest that in the presence of other risk factors, which are often correlated among themselves, variables such as T2DM duration and HbA1c do not markedly contribute to model performance. Similarly, we note that despite comparing risk prediction tools derived across more than three decades, where health and healthcare has generally improved, the almost uniform performance of these scores in the present contemporary sample of T2DM illustrates that this healthcare changes did not affect external performance.

Due to the inherent limitations of EHR data some predictor variables were infrequently measured (e.g., CRP), which we attempted to address through multiple imputation. Possibly this reliance on imputed data biased study results, however we did not observe a meaningful difference in performance between complex model such as the QRISK3 (requiring 19 variables) and more straightforward models such as the SCORE (requiring only 8 variables), indeed with 13 predictors RECODE’s relatively good performance is unlikely explained by missing data. Furthermore, while disease histories and medication history could be readily extracted from before the time of T2DM diagnosis, measured risk factors, such as blood pressure, were extracted using a window of 12 months before and one week after diagnosis. While this does reflect data availability in real-world settings, medical professionals intending to use the risk prediction tools will likely actively measure key variables, especially if this is readily obtained, such as BMI and blood pressure. Thus, we may have underestimated true performance of risk prediction scores in an ideal setting.

The calibration (agreement between observed and predicted risk) was generally reasonable and could readily be improved by recalibrating the models in an independent training set. This recalibration was also successful in repurposing models to predict endpoints outside their intended use. Here we reiterate that all performance metrics, on discrimination (c-statistic) and calibration, were estimated in an independent test dataset fairly assessing performance without the over-optimism observed when calculating these metrics in the same training data used to recalibrate (or when deriving a model de novo). The near optimal calibration additionally highlights that the recalibrated models were not overfitted and utilized a sufficiently larger training sample, which would result in over- or under-estimating of the true risk in an independent test dataset. This is perhaps most clearly shown by comparing the calibration plots presented by van der Leeuw ^8^ derived in a small sample of 584 T2DM patients to the calibration observed in the current analysis using more than 20,000 T2DM patients in Figures 2,4, and 6. Previous studies have typically focused on *any* CVD or individual endpoints such as CHD and *any* stroke, here we show that such models can accurately be used to identify T2DM patients at increased risk for the composite CVD+HF+AF (AF and HF occur much more frequently in T2DM patients^36^). While we showed reasonable out-of-the-box calibration, recalibration improved performance to near perfect agreement and we propose that recalibration is more frequently considered before applying any model to local settings. Given the modest sample size (a few hundred cases) required to accurately recalibrate a model^37^, combined with the increased availability of EHR data, such recalibration could be readily applied by health care commissioners at a local level. To facilitate such recalibration we have appended a straightforward computer application in https://gitlab.com/cvd_in_t2dm/recalibration.

In summary, we show that risk scores derived in the general population work equally well in people with T2DM, even for the wider spectrum of CVD including heart failure and atrial fibrillation that occur more frequently in T2DM patients. In the UK, there appears little reason to move away from the pragmatic choice of the QRISK2 prediction tool. Just under a fifth of all patients with T2DM have a pre-existing CVD diagnosis, prediction tools universally performed badly in this population, explanations and solutions for this require further investigation. Finally, we show that recalibration of a given tool for the population of interest markedly improves performance and should be employed more widely.

## Data Availability

This study was carried out as part of the CALIBER programme. CALIBER, led from the UCL Institute of Health Informatics, is a research resource consisting of anonymised, coded variables extracted from linked electronic health records, methods and tools, specialised infrastructure, and training and support. This study is based in part on data from the Clinical Practice Research Datalink obtained under licence from the UK Medicines and Healthcare products Regulatory Agency. The data is provided by patients and collected by the NHS as part of their care and support. The interpretation and conclusions contained in this study are those of the author/s alone. The interpretation and conclusions contained in this study are those of the author/s alone. Copyright (2020), re-used with the permission of The Health & Social Care Information Centre. All rights reserved.

## Author contributions

FWA, and AFS contributed to the idea and design of the study. KD, JG conducted the literature search. KD prepared dataset for analysis and implemented the risk scores, KD and AFS conducted data analysis and created figures. KD wrote the manuscript with support from FWA, NC, AFS. provided critical input on the analyses and the drafted manuscript.

## Acknowledgements

KD is supported by NPIF programme grant MR/S502522/1. FA is supported by UCL Hospitals NIHR Biomedical Research Centre. JG is supported by BHF grant FS/17/70/33482. AFS is supported by BHF grant PG/18/5033837 and the UCL BHF Research Accelerator AA/18/6/34223. NC is supported by a MRC Unit grant MRC_UU_00019/1.

This study was carried out as part of the <u>CALIBER © programme</u>. CALIBER, led from the UCL Institute of Health Informatics, is a research resource consisting of anonymised, coded variables extracted from linked electronic health records, methods and tools, specialised infrastructure, and training and support. This study is based in part on data from the Clinical Practice Research Datalink obtained under licence from the UK Medicines and Healthcare products Regulatory Agency. The data is provided by patients and collected by the NHS as part of their care and support. The interpretation and conclusions contained in this study are those of the author/s alone. The interpretation and conclusions contained in this study are those of the author/s alone. Copyright © (2020), re-used with the permission of The Health & Social Care Information Centre. All rights reserved.

## Approvals

The study was approved by the MHRA (UK) Independent Scientific Advisory Committee [17_155], under Section 251 (NHS Social Care Act 2006).

## Prior postings and presentations

This study and its results have not been published previously.

