## Appendix for "Cardiovascular risk prediction in type 2 diabetes: a comparison of 22 risk scores in primary care setting"

**Systematic review - search strategy**

The search strategy focused on key words including CVD, type 2 diabetes, risk assessment or risk score and names of known risk scores. Appendix Figure 1 shows the search query to MEDLINE that we used as an extension of the search query previously performed [1]. In order to limit the number of irrelevant articles, we altered most of the Mesh headings to only search for major entries, excluding subheadings. The search query was limited to articles published between 2008-06-30 and 2019-01-16. Reference lists of papers and validation publications such as [1], [2], [3] were screened to identify additional publications reporting risk prediction models.

**Appendix Figure 1 Search query used for literature search. Search query was limited to articles published between 2008-06-30 and 2019-01-16.**

(diabetes mellitus, **type** 2[Majr:NoExp] **OR** diabetes mellitus **type** 2[Title/Abstract] **OR** Diabetic Angiopathies[Majr:NoExp] **OR** diabetic angiopath*[Title/**Abstract**])

**AND**

(risk assessment[Majr:NoExp] **OR** risk assessment*[Title/**Abstract**] **OR** predictive value **of** tests[Majr] **OR** "predictive value of test*"[Title/**Abstract**] **OR** sensitivity **and** specificity[Majr] **OR** reproducibility **of** results[MeSH Terms] **OR** risk score*[Title/**Abstract**] **OR** risk calculat*[Title/**Abstract**] **OR** risk engine* **OR** risk equation*[Title/**Abstract**] **OR** risk algorithm*[Title/**Abstract**] **OR** risk table*[Title/**Abstract**] **OR** risk predict*[Title/**Abstract**] **OR** risk **function***[Title/**Abstract**] **OR** framingham risk*[Title/**Abstract**] **OR** framingham equation*[Title/**Abstract**] **OR** framingham model*[Title/**Abstract**] **OR** PROCAM[Title/**Abstract**] **OR** decode[Title/**Abstract**] **OR** ukpds[Title/**Abstract**] **OR** copenhagen risk*[Title/**Abstract**] **OR** British Regional Heart Study[Title/**Abstract**] **OR** Swedish National Diabetes Register[Title/**Abstract**] **OR** Tayside[Title/**Abstract**] **OR** qrisk[Title/**Abstract**] **OR** Reynolds risk[Title/**Abstract**] **OR** Dundee risk[Title/**Abstract**] **OR** brhs score*[Title/**Abstract**] **OR** British Family Heart Study[Title/**Abstract**] **OR** **New** Zealand chart[Title/**Abstract**] **OR** seven countries study[Title/**Abstract**] **OR** Progetto CUORE[Title/**Abstract**]) **AND** (Cardiovascular Diseases[MeSH Terms] **OR** cardiovascular disease*[Title/**Abstract**] **OR** cardiovascular disorder*[Title/**Abstract**] **OR** Coronary Artery Disease[MeSH Terms] **OR** coronary artery disease[Title/**Abstract**] **OR** coronary artery disorder*[Title/**Abstract**] **OR** Stroke[MeSH Terms] **OR** stroke[Title/**Abstract**])

**Appendix Table 1 Abbreviations used in the appendix**

| Abbreviation | Full text |
|---|---|
| C-HF | Congestive Heart Failure |
| CVD | Cardiovascular disease |
| CHD | Coronary Heart Disease |
| HF | Heart Failure |
| AF | Atrial Fibrillation |
| MI | Myocardial Infarction |
| T2DM | Type 2 diabetes |
| HbA1c | Glycated haemoglobin |
| SBP | Systolic Blood Pressure |
| DBP | Diastolic Blood Pressure |
| ECG | Electrocardiography |
| HIV | Human Immunodeficiency Virus |
| Weibull AFT model | Weibull Accelerated Failure Time model |
| Cox's PH model | Cox Proportional-Hazards Model |
| CS | Calibration Slope |
| CIL | Calibration-in-the-large |

### Outlier Removal

#### Appendix Table 2  Summary of applied cut-offs for the continuous variables.

| Clinical characteristics | Valid data ranges (applied cut-offs) |
|---|---|
| Age (years) | [18.0, 100.0] |
| HbA1c (mmol / mol) | [20.0, 120.0] |
| HDL cholesterol (mmol / L) | [0.0, 20.0] |
| Total cholesterol (mmol / L) | [0.0, 20.0] |
| SBP (mm Hg) | [70, 250] |
| DBP (mm Hg) | [40, 150] |
| Urine albumin / creatinine ratio (mg / mmol) | [0.0, 200] |
| Serum creatinine (micromol / L) | [0.0 - 900] |

**Appendix Table 3 Characteristics of the evaluated CVD risk scores.**

| Name | Presented models (number of predictors$ Appendix Table 8) | Year of publication | Derivation sample | Derived with/ without T2DM subjects | Statistical model | Reported type of predicted CVD | Evaluated type of CVD |
|---|---|---|---|---|---|---|---|
| FINRISK [4] | Finrisk Stroke (8)<br>Finrisk CHD (8)<br>Finrisk CVD (8) | 2016 | National FINRISK Study:<br>random population samples of individuals 30 to 64 years of age - North Karelia (Finland); n = 9,391 men and 10,056 women | with T2DM (diabetes included as a risk factor) | Regression model | CHD (MI, unstable angina pectoris, death from CHD), Stroke (infractions hemorrhage), CVD | Stroke, CHD, CVD |
| DARTS (Diabetes Audit and Research in Tayside, Scotland) [5] | DARTS (10) | 2006 | Tayside, Scotland UK, T2DM without history of CHD | with T2DM (100% T2DM subjects) | Weibull AFT model | First major CHD event (fatal or non-fatal acute MI or CHD death) | CHD |
| Framingham 1991 [6] | Framingham 1991 Fatal CHD (8)<br>Framingham 1991 CVD (8)<br>Framingham 1991 Stroke (8) | 1991 | 5,573 US white men and women aged 30 - 74 years without cancer and CVD | with T2DM (diabetes included as a risk factor) | Weibull AFT model | Myocardial infarction, CHD, fatal CHD, stroke, CVD, death from CVD | CHD, CVD, Stroke |
| Framingham 1998 [7] | Framingham 1998 (7) | 1998 | 5,345 US white men and women aged 30 - 74 | with T2DM (diabetes included as a risk factor) | Cox's PH model | CHD (angina pectoris, recognised and unrecognised MI, coronary insufficiency, CHD death) Hard CHD included total CHD without angina pectoris | CHD |
| QRISK 2 [8] | QRISK 2 (13) | 2008 | 2.3 million people, aged 35 - 74, England and Wales, people without previous CVD | with T2DM (diabetes included as a risk factor) | Cox's PH model | CHD, stroke, or transient ischemic attack; not peripheral | CVD |
| QRISK 3 [9] | QRISK 3 (19) | 2017 | 7.89 million people, aged 25 -84 years, without CHD and not prescribed statins | with T2DM (diabetes included as a risk factor) | Cox's PH model | CVD defined as a composite outcome of coronary heart disease, ischaemic stroke, or transient ischaemic attack | CVD |
| SCORE [10] | SCORE CHD (6)<br>SCORE CVD (6) | 2003 | 205,178 European men and women, aged 19 - 80 years without history of MI | without T2DM | Weibull PH model | CHD, CVD, CVD mortality | CHD, CVD |

| | | | | | | | |
|---|---|---|---|---|---|---|---|
| UKPDS 56 [11] | UKPDS 56 (9) | 2001 | 4,540 people from U.K. Prospective Diabetes Study, aged 25 - 65 years, without MI, angina or heart failure | with T2DM (100% T2DM subjects) | Cox's PH model | Fatal or non-fatal MI or sudden death | CHD |
| UKPDS 68 [12] | UKPDS 68 C-HF (5)<br>UKPDS 68 Stroke (10) | 2004 | 3,641 people with diagnosed diabetes type 2, aged 25 - 65 years | with T2DM (100% T2DM subjects) | Weibull PH model | CVD: MI (fatal and non-fatal myocardial infarction and sudden death), IHD, CHF, Cerebrovascular disease: first non-fatal stroke | HF, Stroke |
| UKPDS 82 [13] | UKPDS 82 C-HF (10)<br>UKPDS 82 CHD (15) | 2013 | 5,102 people with diagnosed diabetes type 2, age 25 - 65 years | with T2DM (100% T2DM subjects) | Weibull PH model | MI: acute myocardial infarction or sudden death; Stroke: non-fatal stroke or fatal stroke); chronic ischemic heart disease (IHD); Congestive heart failure (CHF) | HF, CHD |
| ASCVD Pooled Cohort Risk Equations [14] | ASCVD (9) | 2013 | 24,626 people, white and African-American, aged 40 - 79 years, without CVD | with T2DM (diabetes included as a risk factor) | Cox's PH model | ASCVD event (CHD death, nonfatal myocardial infarction, or fatal or nonfatal stroke) | CVD |
| RECODE [15] | RECODE (13) | 2017 | 9,635 people from Action to Control Cardiovascular Risk in Diabetes Study | with T2DM (100% T2DM subjects) | Cox PH model | MI, stroke, CHF, cardiovascular mortality | CVD + HF |
| Reynolds Risk ** [16] [17] | Reynolds Risk (9) | 2007, 2008 | Reynolds Risk Score for Men: 10,724 healthy men without diabetic and CVD, aged over 50 years<br>Reynolds Risk Score for Women: 24,558 healthy women, aged over 45 years | with T2DM (diabetes included as a risk factor) | Cox PH model | CVD, CHD, MI, stroke, coronary revascularization, cardiovascular death | CVD |
| CHS (Cardiovascular Health Study) * [18] | CHS Basic (8)<br>CHS Advanced (9) | 2013 | 782 people with diabetes, without CVD, CHF and AF, aged over 65 years | with T2DM (100% T2DM subjects) | Cox PH model | MI, stroke and death | CVD |

*- based on the implementation provided from the Utrecht research team

**- Combined two risk scores: Reynolds Risk Score for Men, Reynolds Risk Score for Women

**Appendix Table 4 Predictor variables included in cardiovascular prediction models. (*) Type 2 diabetes means either a risk factor or T2DM derivation population of a risk score.**

| | DARTS [5](a) | UKPDS 56 [11](a) | UKPDS 68 [12](a) | UKPDS 82 [13](a) | RECODE [15](a) | CHS [18](a) | FINRISK [4](b) | Framingham 1991 [6] (b) | Framingham 1998 [7](b) | QRISK 2 [8](b) | QRISK 3 [9](b) | ASCVD [14](b) | Reynolds Risk [16][17](b) | SCORE [10](c) | **Number of publications** |
|---|---|---|---|---|---|---|---|---|---|---|---|---|---|---|---|
| **Age** | ✓ | ✓ | ✓ | ✓ | ✓ | ✓ | ✓ | ✓ | ✓ | ✓ | ✓ | ✓ | ✓ | ✓ | 14 |
| **Sex** | ✓ | ✓ | ✓ | ✓ | ✓ | | ✓ | ✓ | ✓ | ✓ | ✓ | ✓ | ✓ | ✓ | 13 |
| **Smoking status** | ✓ | ✓ | ✓ | ✓ | ✓ | ✓ | ✓ | ✓ | ✓ | ✓ | ✓ | ✓ | ✓ | ✓ | 14 |
| **SBP or DBP** | ✓ | ✓ | ✓ | ✓ | ✓ | ✓ | ✓ | ✓ | ✓ | ✓ | ✓ | ✓ | ✓ | ✓ | 14 |
| **Total cholesterol** | ✓ | ✓ | ✓ | | ✓ | ✓ | ✓ | ✓ | ✓ | ✓ | ✓ | ✓ | ✓ | ✓ | 13 |
| **HDL cholesterol** | | ✓ | ✓ | ✓ | ✓ | ✓ | ✓ | ✓ | ✓ | ✓ | | ✓ | ✓ | ✓ | 12 |
| **LDL cholesterol** | | | | ✓ | | | | | | | | | | | 1 |
| **Type 2 diabetes*** | ✓ | ✓ | ✓ | ✓ | ✓ | ✓ | ✓ | ✓ | ✓ | ✓ | ✓ | ✓ | ✓ | | 13 |
| **Duration of diabetes** | ✓ | | | ✓ | | | | | | | | | | | 2 |
| **$HbA_{1c}$** | ✓ | ✓ | ✓ | ✓ | ✓ | | | | | | | | | | 5 |
| **Treated hypertension** | ✓ | | | | ✔ | | | | | ✓ | ✓ | ✓ | | | 5 |
| **Height** | ✔ | | | | | | | | | | | | | | 1 |
| **Left Ventricular Hypertrophy (LVH) ECG** | | | | | | | | ✔ | | | | | | | 1 |

| | | | | | | | | | | | | | | | |
|---|---|---|---|---|---|---|---|---|---|---|---|---|---|---|---|
| **Ethnicity** | | ✔ | | ✔ | ✔ | | | | | ✔ | ✔ | ✔ | | | 6 |
| **BMI** | | | ✔ | ✔ | | | | | | ✔ | | | | | 3 |
| **AF** | | | ✔ | ✔ | | | | | | ✔ | ✔ | | | | 4 |
| **Townsend Social Deprivation Score** | | | | | | | | | | ✔ | ✔ | | | | 2 |
| **Rheumatoid Arthritis (RA)** | | | | | | | | | | ✔ | ✔ | | | | 2 |
| **Family history of CVD** | | | | | | | | | | | ✔ | | ✔ | | 2 |
| **Type 1 diabetes** | | | | | | | | | | | ✔ | | | | 1 |
| **Chronic kidney disease (CKD)** | | | | | | | | | | | ✔ | | | | 1 |
| **Migraine** | | | | | | | | | | | ✔ | | | | 1 |
| **Corticosteroid use** | | | | | | | | | | | ✔ | | | | 1 |
| **Systemic lupus** | | | | | | | | | | | ✔ | | | | 1 |
| **Atypical antipsychotic** | | | | | | | | | | | ✔ | | | | 1 |
| **Severe mental illness** | | | | | | | | | | | ✔ | | | | 1 |
| **History of CVD** | | | ✔ | ✔ | ✔ | | ✔ | | | | | | | | 4 |
| **Peripheral vascular disease (PVD)** | | | | ✔ | | | | | | | | | | | 1 |
| **White Blood Count (WBC)** | | | | ✔ | | | | | | | | | | | 1 |
| **Presence of micro- or macro albuminuria** | | | | ✔ | | | | | | | | | | | 1 |

| | | | | | | | | | | | | | | | |
|---|---|---|---|---|---|---|---|---|---|---|---|---|---|---|---|
| **Serum Creatinine** | | | | | ✔ | ✔ | | | | | | | | | 2 |
| **Urine albumin to creatinine ratio** | | | | | ✔ | | | | | | | | | | 1 |
| **High-sensitivity C-reactive protein (hsCRP)** | | | | | | ✔ | | | | | | | ✔ | | 2 |
| **Use of glucose-lowering medications** | | | | | | ✔ | | | | | | | | | 1 |
| **Use of statins or anticoagulants** | | | | | ✔ | | | | | | | | | | 1 |
| **Total number of predictors** | 10 | 9 | 11 | 16 | 13 | 9 | 8 | 8 | 7 | 13 | 19 | 9 | 9 | 6 | |

n.b Derivation population: (a) T2DM population, (b) General population, (c) General population excluding T2DM subjects

**Appendix Figure 2 Number of predictors used in each of the CVD risk scores**

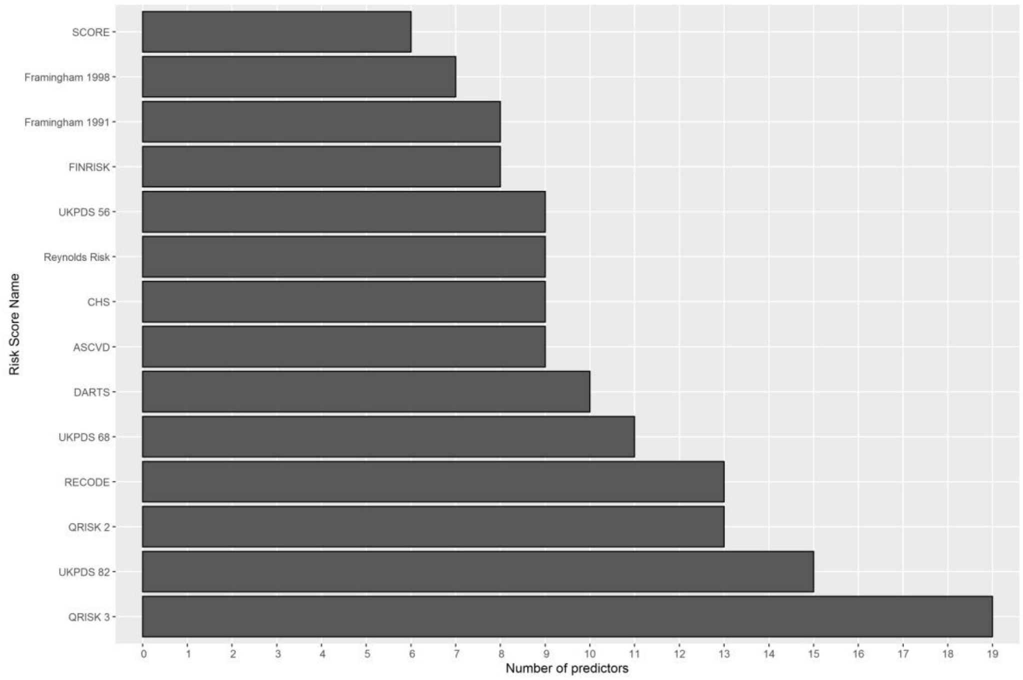

**Appendix Table 5 Outcomes definitions based on CALIBER dataset.**

| Type of cardiovascular disease | Link to CALIBER research platform |
|---|---|
| Fatal and non fatal MI | https://www.caliberresearch.org/portal/show/phenotype_mi<br><br>https://www.caliberresearch.org/portal/show/phenotype_chd_nos |
| Fatal or non fatal Stroke | https://www.caliberresearch.org/portal/show/phenotype_stroke_intracerebral_haem,<br>https://www.caliberresearch.org/portal/show/phenotype_stroke_ischaemic,<br>https://www.caliberresearch.org/portal/show/phenotype_stroke_nos,<br>https://www.caliberresearch.org/portal/show/phenotype_stroke_subarachnoid |
| Fatal peripheral vascular disease | https://www.caliberresearch.org/portal/show/phenotype_pad |
| Sudden Cardiac Death | https://www.caliberresearch.org/portal/show/phenotype_scd |
| HF | https://www.caliberresearch.org/portal/show/phenotype_hf |
| AF | https://www.caliberresearch.org/portal/show/phenotype_af |
| Ischemic stroke | https://www.caliberresearch.org/portal/show/phenotype_stroke_ischaemic<br><br>https://www.caliberresearch.org/portal/show/phenotype_stroke_nos |
| Haemorrhagic stroke | https://www.caliberresearch.org/portal/show/phenotype_stroke_intracerebral_haem,<br><br>https://www.caliberresearch.org/portal/show/phenotype_stroke_subarachnoid |

**Appendix Table 6 T2DM definition based on CALIBER dataset. This is a composite of GP diagnosis of diabetes, diabetes diagnoses from primary and secondary care.**

| Definition / Data Source | Link to CALIBER research platform |
|---|---|
| **Diabetes Phenotype definition** | https://www.caliberresearch.org/portal/show/phenotype_diabetes (extract patients with type 2 diabetes) |
| GP diagnosis of diabetes | https://www.caliberresearch.org/portal/show/diabdiag_gprd |
| Diabetes diagnosis (primary care) | https://www.caliberresearch.org/portal/show/dm_gprd |
| Diabetes diagnosis (secondary care) | https://www.caliberresearch.org/portal/show/dm_hes<br>ICD-10 codes: E10-13, O240-243, G590, G632, H280, H360, M142, N083 |

**Appendix Table 7 Baseline characteristics of T2DM patients at the time of diagnosis.**

| **Clinical characteristics** | **Available sample (%)** |
|---|---|
| Total no. of subjects | 203,172 |
| Rheumatoid arthritis | 2,313 (1.1) |
| Renal disease | 12,145 (6.0) |
| Exercise ECG results | 6,909 (3.4) |
| Anticoagulants use | 710 (0.3) |
| Diuretic use | 64,524 (31.8) |
| Corticosteroids use | 11,347 (5.6) |
| Foot amputation | 155 (0.1) |
| Systemic lupus erythematosus | 218 (0.1) |
| HIV positive | 131 (0.1) |
| Mental disorders | 1,399 (0.7) |
| Microalbuminuria | 3 (0.0) |
| Erectile dysfunction disorder | 8,406 (4.1) |
| Erectile dysfunction drugs | 7,638 (3.8) |
| Hypertension | 139,463 (68.6) |
| Atrial Fibrillation | 20,503 (10.1) |
| Migraine | 920 (0.5) |
| Treated hypertension (blood lowering medication) | 99,527 (49.0) |
| Family history of CVD | 52,949 (26.1) |
| Family history of CHD | 439 (0.2) |
| Family history of MI | 57 (0.0) |
| Family history of Stroke | 426 (0.2) |

**Appendix Table 8 Ethnic origin of T2DM patients.**

| Clinical characteristics | Available sample (%) |
|---|---|
| Total no. of subjects | 203,172 |
| White British | 45,010 (22.2) |
| White Irish | 1,160 (0.6) |
| White Other | 3,608 (1.8) |
| White NOS | 8,496 (4.2) |
| Mixed White and Black | 335 (0.2) |
| Mixed White and Asian | 144 (0.1) |
| Mixed Asian and Black | 90 (0.0) |
| Mixed Other | 275 (0.1) |
| Mixed NOS | 31 (0.0) |
| Indian | 4403 (2.2) |
| Pakistani | 2,335 (1.1) |
| Bangladeshi | 926 (0.5) |
| Other Asian | 2,235 (1.1) |
| Black Caribbean | 1,893 (0.9) |
| Black African | 2,612 (1.3) |
| Black Other | 740 (0.4) |
| Chinese | 414 (0.2) |
| Other Ethnic Group | 2,118 (1.0) |
| Ethnic Group not specified | 126,347 (62.2) |

**Appendix Table 9 Clinical characteristics of the continuous variables around the time (1 year before - 1 week after) of diagnosis of T2DM.**

| Clinical characteristics | Mean (SD) or N (%) | Median (Q1; Q3) | Missing data (% of total no. of subjects) |
|---|---|---|---|
| Total no. of subjects | 203,172 | | |
| Total cholesterol (mmol / L) | 5.3 (1.3) | 5.2 (4.4; 6.1) | 36.9 |
| Triglycerides (serum and plasma) (mmol / L) | 2.4 (2.2) | 1.9 (1.3; 2.7) | 47.8 |
| DBP (mm Hg) | 81 (11) | 80 (75; 88) | 24.8 |
| Urine albumin to creatinine ratio | 3.8 (11.6) | 1.1 (0.6; 2.5) | 89.3 |
| Serum creatinine (micromol / L) | 85.7 (26.9) | 83.0 (71.0; 96.0) | 32.8 |
| C reactive protein (mg / L) | 12.6 (25.9) | 6.0 (3.0; 12.0) | 91.6 |
| Total white blood cell count (10^9 / L) | 7.7 (2.3) | 7.4 (6.2; 8.8) | 52.2 |

**Appendix Table 10 Comparison of clinical characteristics of the continuous variables around the time (1 year before - 1 week after) and (1 year before - 3 months after) of diagnosis of T2DM.**

| | **Timeframe: 1 year before and 1 week after the diagnosis of T2DM** | | | **Timeframe: 1 year before and 3 months after the diagnosis of T2DM** | | |
|---|---|---|---|---|---|---|
| **Clinical characteristics** | **Mean (SD) or N (%)** | **Median (Q1; Q3)** | **Missing data** (% of total no. of subjects) | **Mean (SD) or N (%)** | **Median (Q1; Q3)** | **Missing data** (% of total no. of subjects) |
| Total no. of subjects | 203,172 | | | 203,172 | | |
| Follow-up time (years) | | 10.0 (6.6; 10.0) | | | 10.0 (6.6; 10.0) | |
| Women (%) | 9,108 (44.8) | | 0.0 | 91,008 (44.8) | | 0.0 |
| Age (years) | 60.9 (14.0) | 61.0 (51.0; 71.0) | 0.0 | 60.9 (14.0) | 61.0 (51.0; 71.0) | 0.0 |
| HbA1c (mmol/mol) | 63.3 (20.2) | 56.3 (48.6; 74.0) | 55.0 | 59.9 (18.3) | 54.1 (47.5; 68.3) | **39.0** |
| FPG (mmol / L) | 9.5 (3.9) | 8.0 (7.1; 10.6) | 68.5 | 9.2 (3.7) | 7.9 (7.0; 10.1) | 67.0 |
| BMI (kg / m^2) | 31.7 (6.7) | 30.7 (27.1; 35.2) | 39.9 | 31.3 (6.5) | 30.4 (26.8; 34.7) | **29.0** |
| HDL cholesterol (mmol / L) | 1.2 (0.4) | 1.2 (1.0; 1.4) | 47.4 | 1.2 (0.4) | 1.2 (1.0; 1.4) | **41.0** |
| LDL cholesterol (mmol / L) | 3.1 (1.1) | 3.0 (2.3; 3.8) | 58.8 | 2.9 | 2.8 (2.2; 3.6) | **53.0** |
| SBP (mm Hg) | 140 (18) | 140 (130; 150) | 24.8 | 137 (17) | 136 (126; 145) | **20.0** |

**Appendix Table 11 The 10-year incidence of cardiovascular disease after a type 2 diabetes diagnosis.**

| Type of cardiovascular disease | No. events (%) |
|---|---|
| Total no. of subjects | 203,172 |
| CVD, HF, or AF | 63,000 (31.07) |
| CVD | 51,636 (25.46) |
| CHD | 40,243 (19.84) |
| Any stroke | 10,413 (5.14) |
| HF | 16,993 (8.38) |
| AF | 20,506 (10.11) |

n.b. One patient could have more than one event, but the event type is unique.

**Appendix Figure 3 UpSet plot [19] presenting the intersection between different outcomes.**

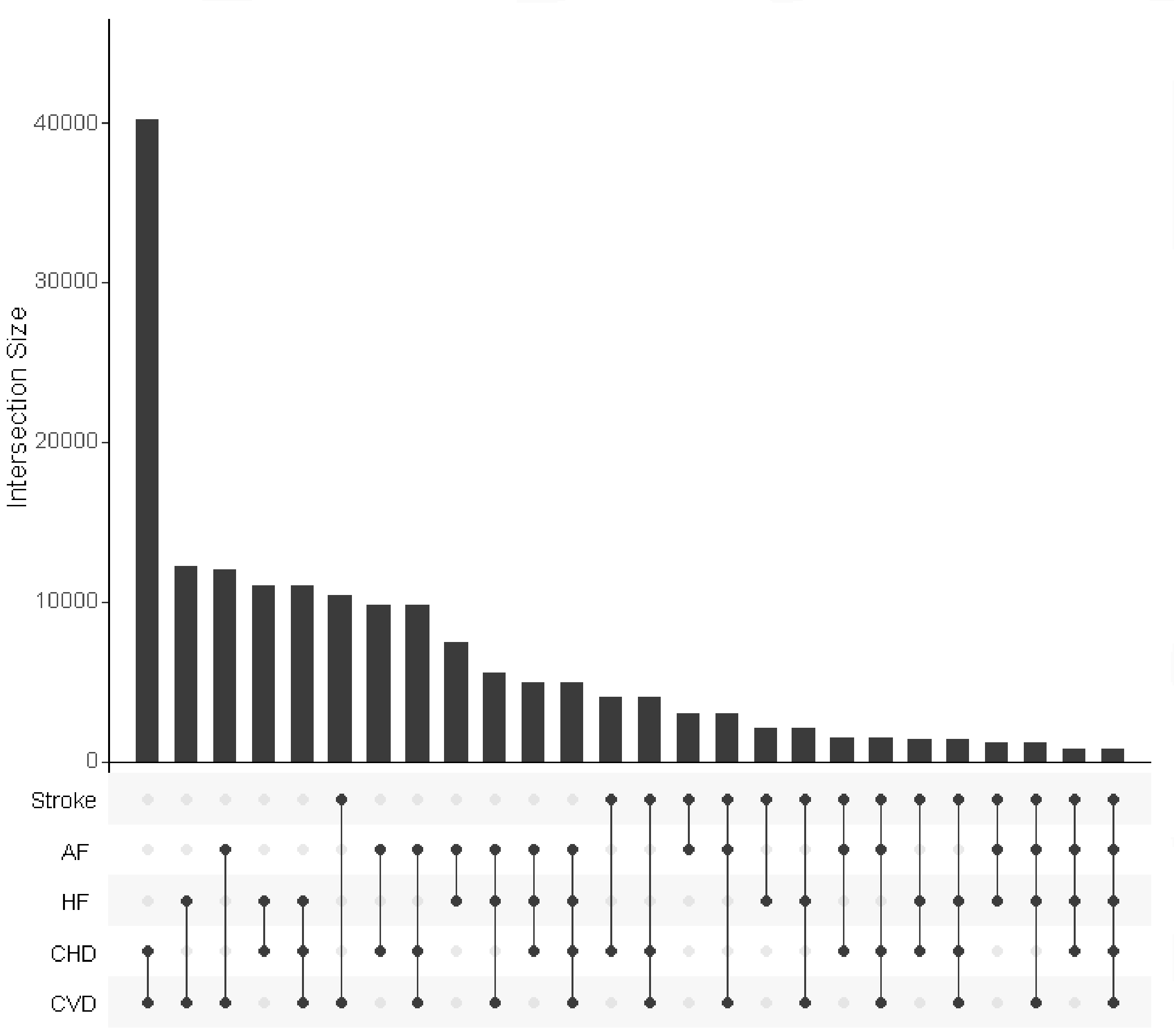

n.b. UpSet plots the intersections of a set as a matrix, shown at the bottom of bar plot. Each column corresponds to a set, and each row in a matrix corresponds to one segment in a Venn diagram. Cells are either empty (light-gray), indicating that this set is not part of that intersection, or filled, showing that the set is participating in the intersection.

**Calibration and Discrimination**

The calibration process describes the agreement between the predicted 10-year risk and the observed 10-year risk. Calibration-in-the-large statistics compare the mean predicted risk and mean observed risks. Calibration-in-the-large statistics of value 0 and calibration slopes of value 1 indicate good model's calibration.

The discrimination process describes model's ability to distinguish patients who will develop the CVD event and patients who will not get the CVD event. The discrimination was assessed by calculating Harrell C statistic, a C statistic of value 1 denotes perfect discrimination, and a value of 0.5 denotes almost random prediction.

**Calibration**

**Appendix Figure 4: Calibration plots of CVD prediction rules calculated based on a complete-case analysis**

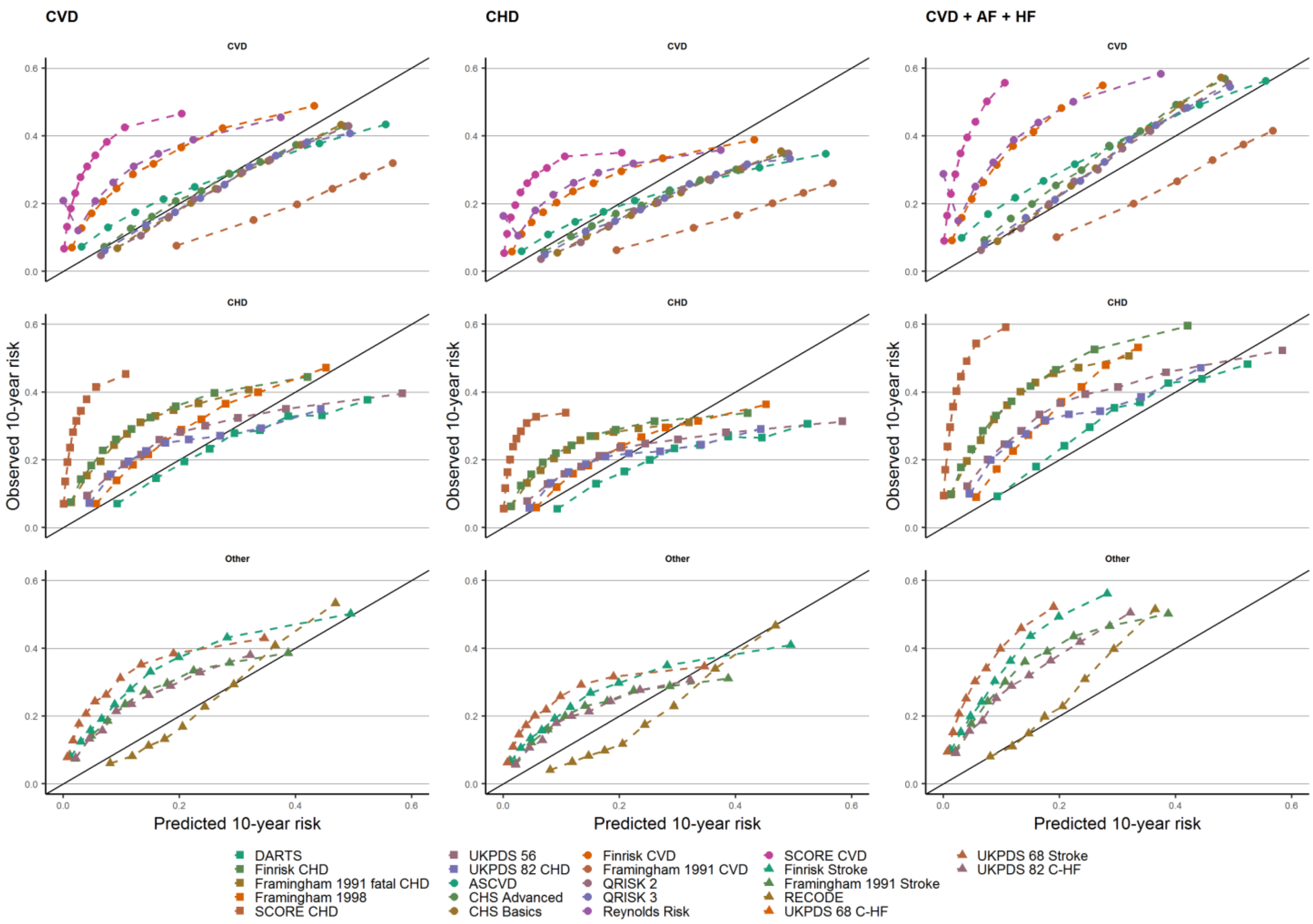

n.b. estimates based on complete-cases from UK sample of T2DM patients. The observed 10-years risk is (y-axes) plotted against the average predicted 10-year risk (x-axis) within groups defined by quintiles of predicted risk. The columns indicate the type of CVD the scores were evaluated against (CVD, CHD, CVD + AF + HF). Scores were grouped by the derivation outcomes CVD, CHD, or other (including Stroke, C-HF)). The diagonal line reflects perfect calibration.

**Appendix Figure 5: Calibration plots of CVD prediction rules calculated based on a complete-case analysis**

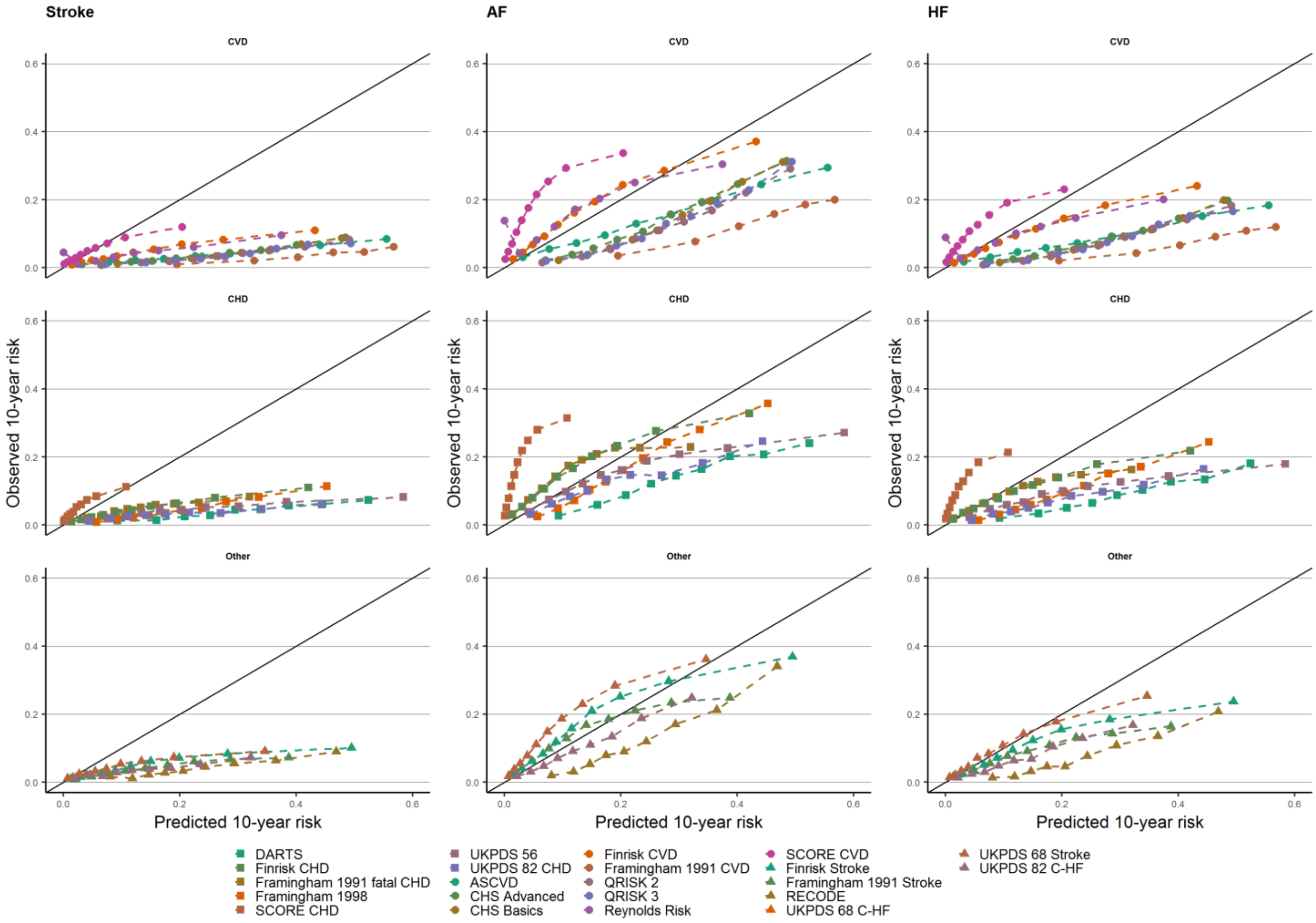

n.b. estimates based on complete-cases from UK sample of T2DM patients. The observed 10-years risk is (y-axes) plotted against the average predicted 10-year risk (x-axis) within groups defined by quintiles of predicted risk. The columns indicate the type of CVD the scores were evaluated against (Stroke, AF, HF). Scores were grouped by the derivation outcomes CVD, CHD, or other (including Stroke, C-HF)). The diagonal line reflects perfect calibration.

**Appendix Figure 6**: **Calibration plots of CVD prediction rules applied in an UK sample of T2DM patients.**

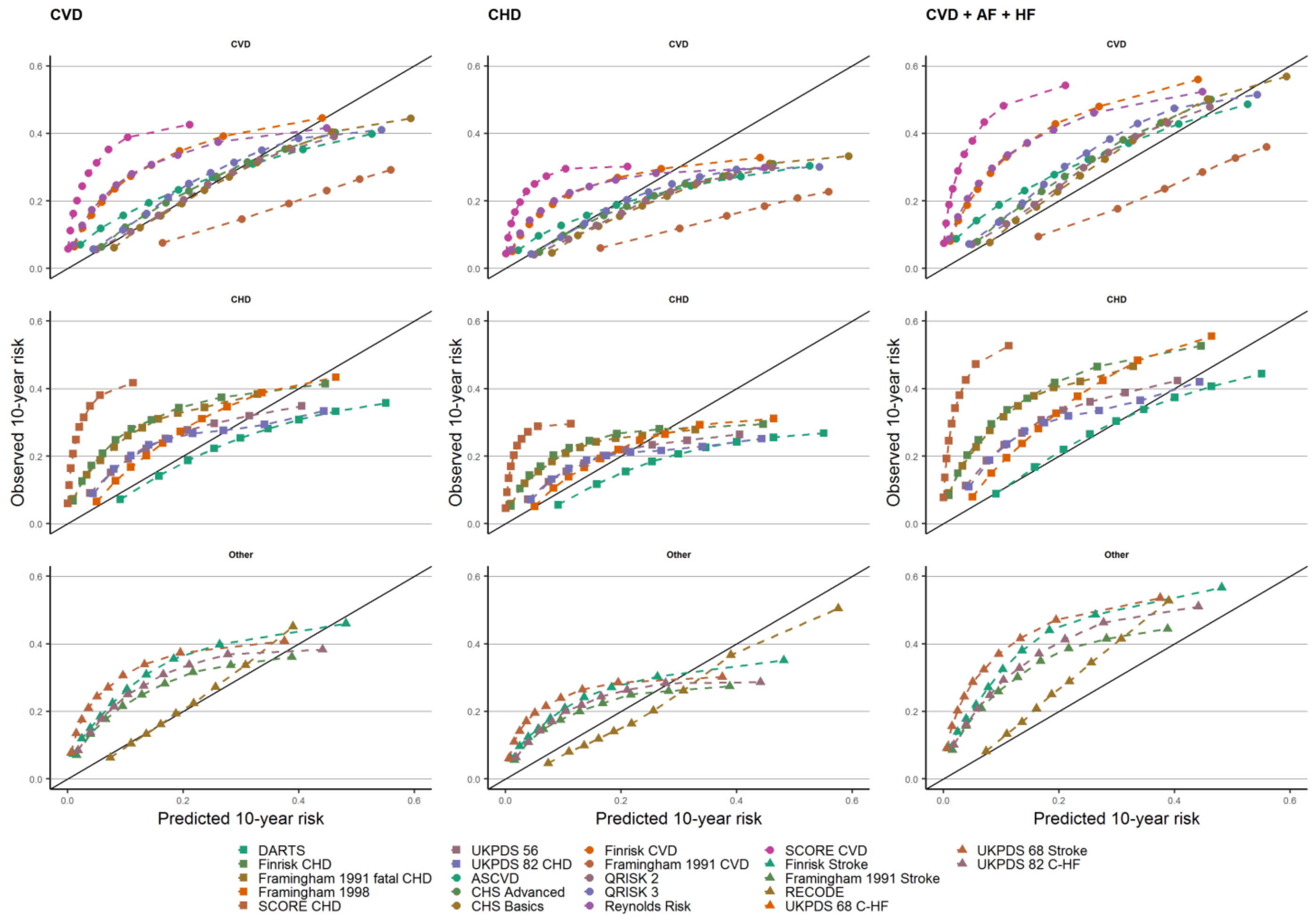

n.b. estimates based on imputed data. The observed 10-years risk is (y-axes) plotted against the average predicted 10-year risk (x-axis) within groups defined by quintiles of predicted risk. The columns indicate the type of CVD the scores were evaluated against (CVD, CHD, CVD + AF + HF). Scores were grouped by the derivation outcomes CVD, CHD, or other (including Stroke, C-HF)). The diagonal line reflects perfect calibration.

**Appendix Figure 7**: **Calibration plots of CVD prediction rules applied in an UK sample of T2DM patients.**

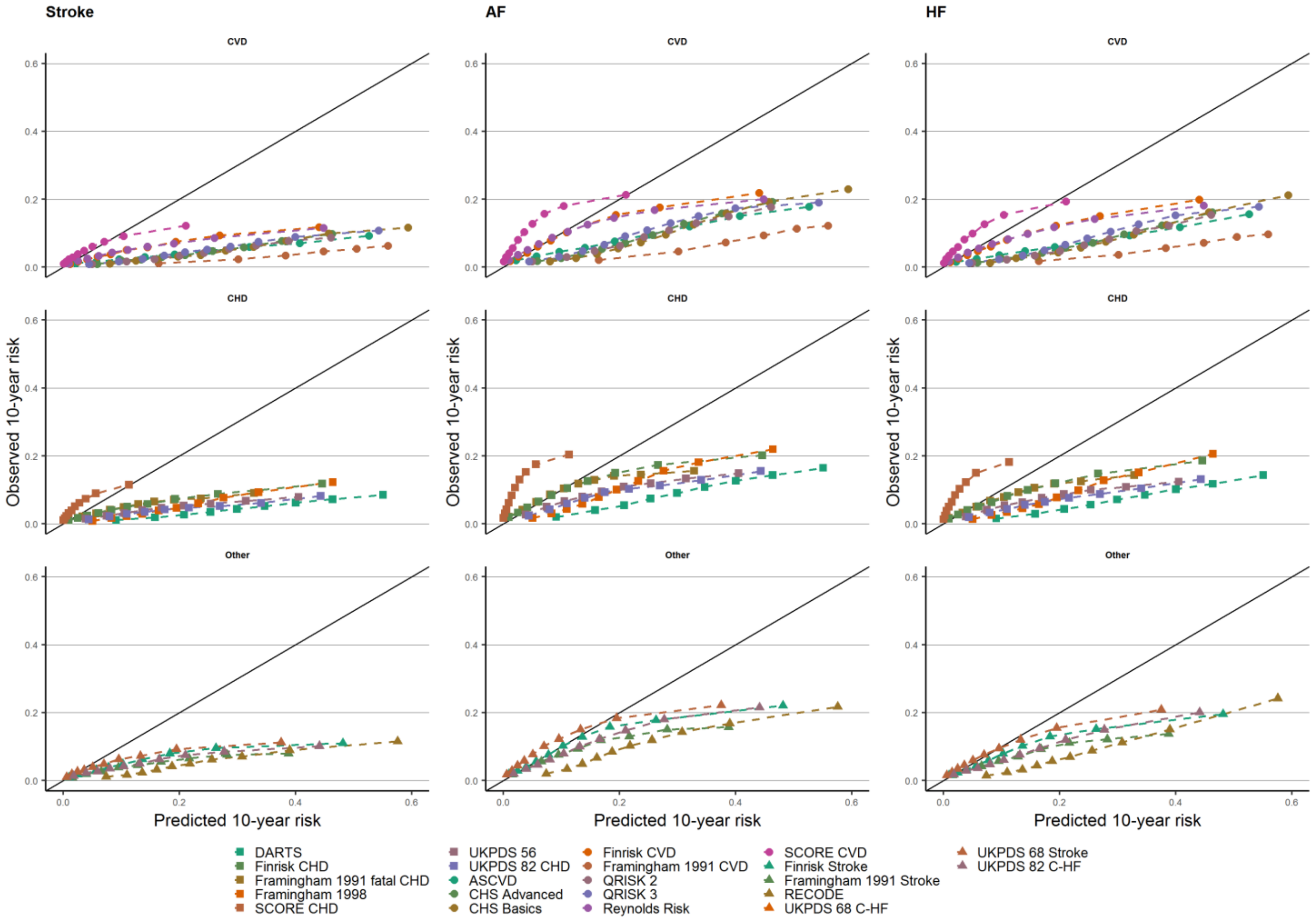

n.b. estimates based on imputed data. The observed 10-years risk is (y-axes) plotted against the average predicted 10-year risk (x-axis) within groups defined by quintiles of predicted risk. The columns indicate the type of CVD the scores were evaluated against (Stroke, AF, HF). Scores were grouped by the derivation outcomes CVD, CHD, or other (including Stroke, C-HF)). The diagonal line reflects perfect calibration.

**Performance after recalibration**

**Appendix Figure 8: Calibration plots after recalibration of the cardiovascular diseases based on a complete-case analysis.**

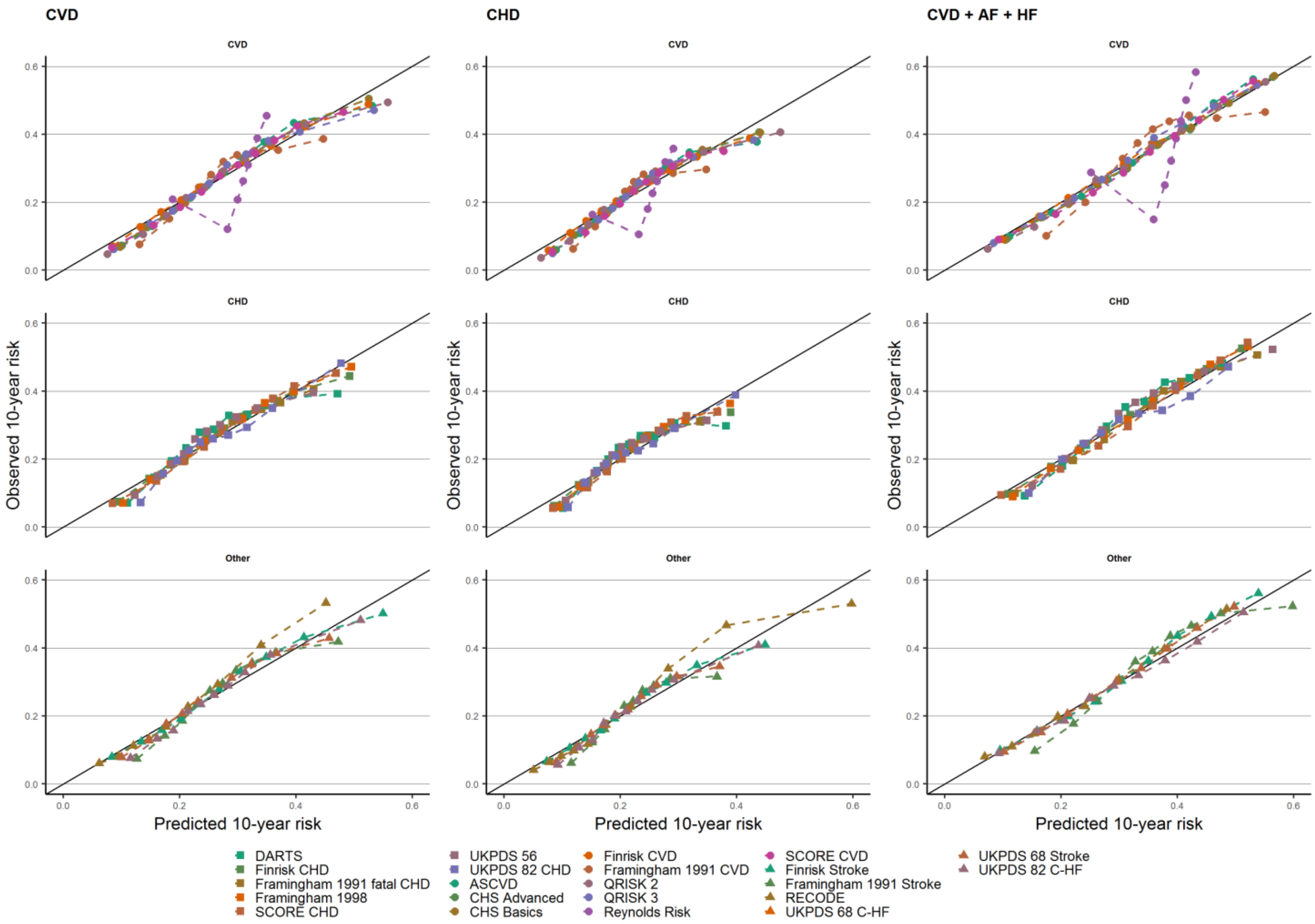

n.b. estimates based on complete-case analysis. The observed 10-years risk is (y-axes) plotted against the average predicted 10-year risk (x-axis) within groups defined by quintiles of predicted risk. The columns indicate the type of CVD the scores were evaluated against (CVD, CHD, CVD + AF + HF). Scores were grouped by the derivation outcomes CVD, CHD, or other (including Stroke, C-HF)). The diagonal line reflects perfect calibration.

**Appendix Figure 9: Calibration plots after recalibration of the cardiovascular diseases based on a complete-case analysis.**

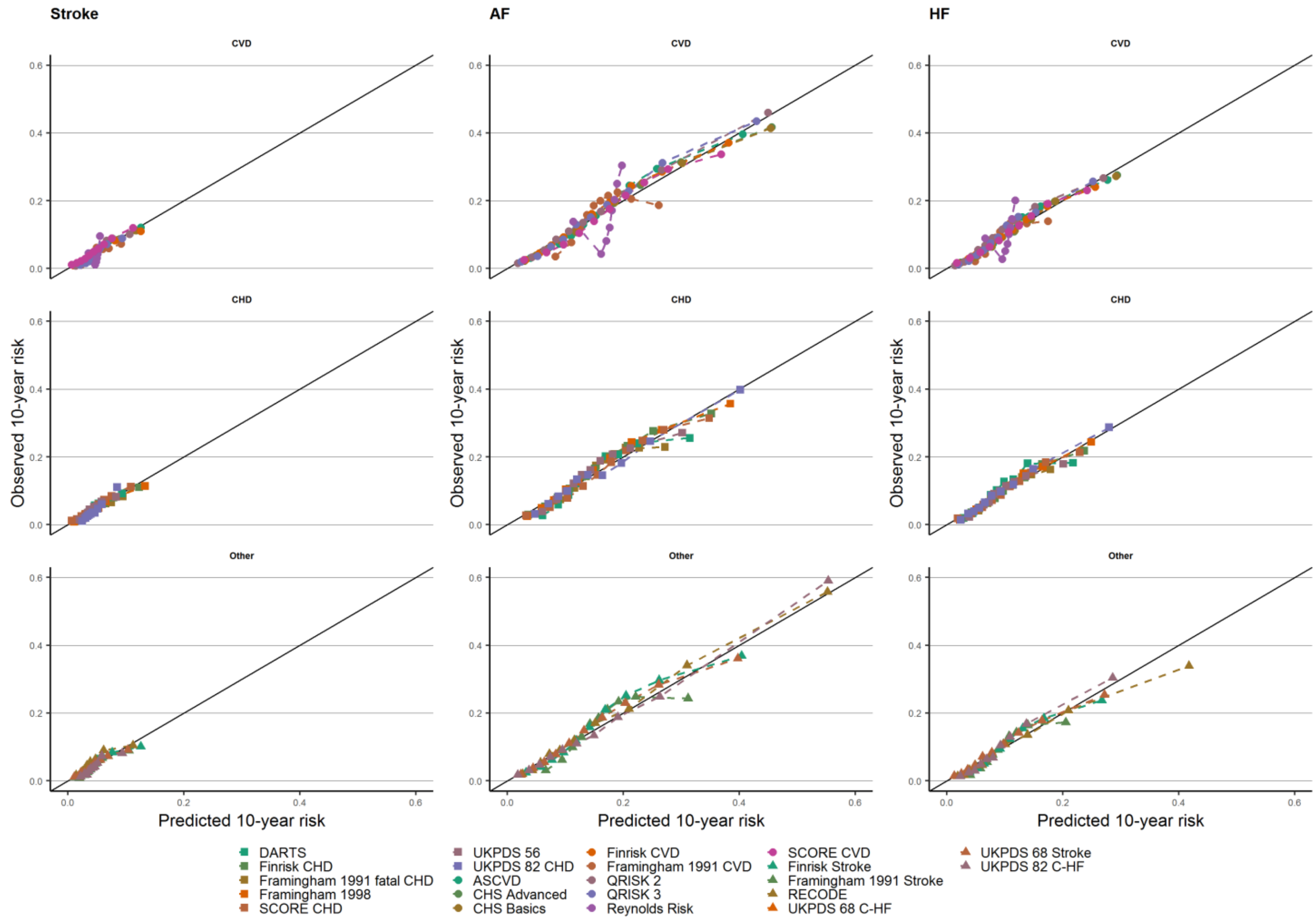

n.b. estimates based on complete-case analysis. The observed 10-years risk is (y-axes) plotted against the average predicted 10-year risk (x-axis) within groups defined by quintiles of predicted risk. The columns indicate the type of CVD the scores were evaluated against (Stroke, AF, HF). Scores were grouped by the derivation outcomes CVD, CHD, or other (including Stroke, C-HF)). The diagonal line reflects perfect calibration.

**Appendix Figure 10: Calibration plots after recalibration of the CVD prediction rules calculated using the imputed datasets.**

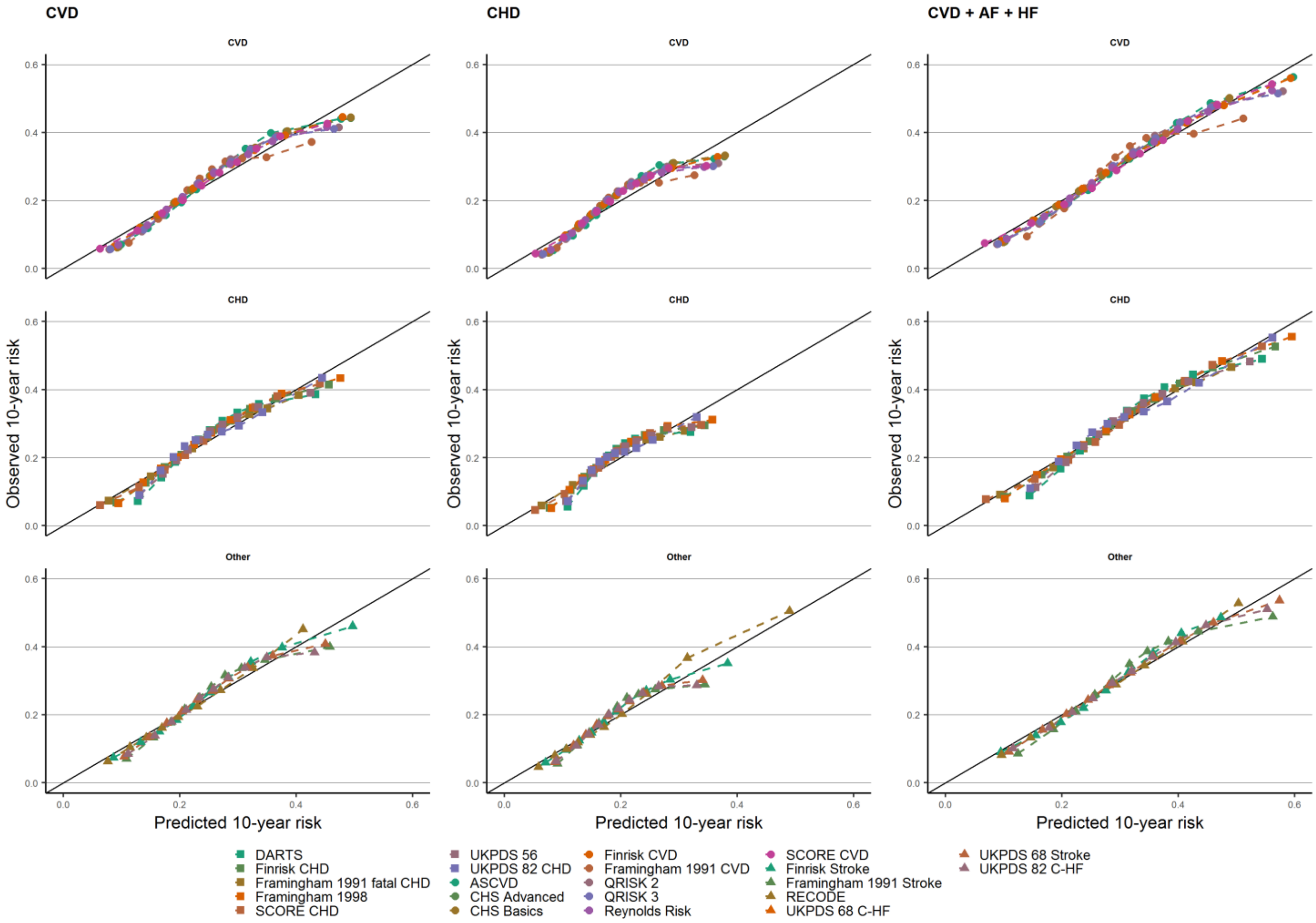

n.b. estimates based on imputed data. The observed 10-years risk is (y-axes) plotted against the average predicted 10-year risk (x-axis) within groups defined by quintiles of predicted risk. The columns indicate the type of CVD the scores were evaluated against (CVD, CHD, CVD + AF + HF). Scores were grouped by the derivation outcomes CVD, CHD, or other (including Stroke, C-HF)). The diagonal line reflects perfect calibration.

**Appendix Figure 11: Calibration plots after recalibration of the CVD prediction rules calculated using the imputed datasets.**

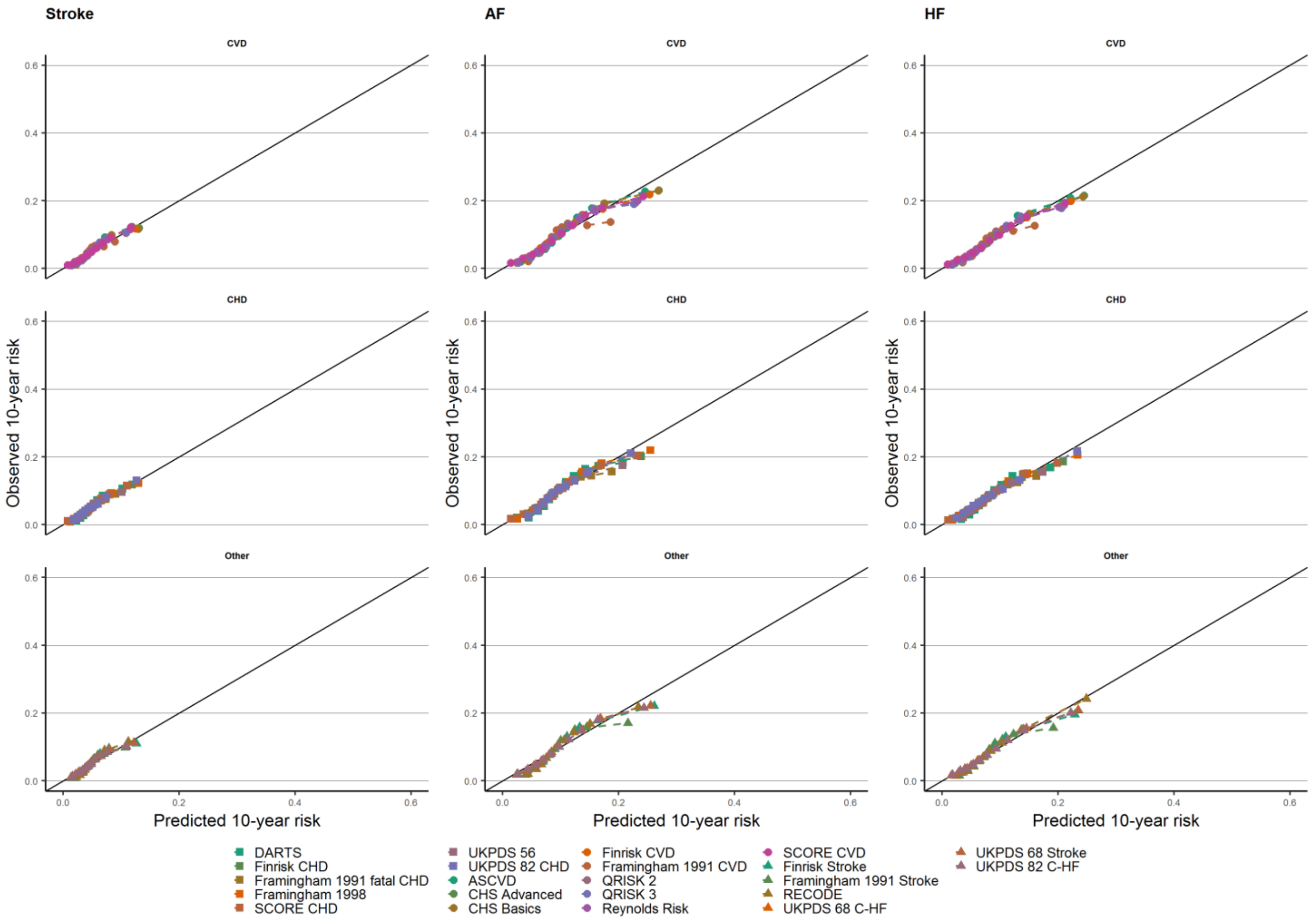

n.b. estimates based on imputed data. The observed 10-years risk is (y-axes) plotted against the average predicted 10-year risk (x-axis) within groups defined by quintiles of predicted risk. The columns indicate the type of CVD the scores were evaluated against (Stroke, AF, HF). Scores were grouped by the derivation outcomes CVD, CHD, or other (including Stroke, C-HF)). The diagonal line reflects perfect calibration.

**Performance before recalibration based on a complete-case analysis.**

Discrimination

Most of the risk scores has a moderate discrimination in CALIBER dataset, the C-statistic ranged from 0.62 (*Reynolds Risk*) to 0.77 (*RECODE*) for CVD outcome; from 0.62 to 0.77 (*RECODE*) for CHD outcome; from 0.62 (*Reynolds Risk*) to 0.80 (*RECODE*) for CVD, AF and HF outcomes; from 0.58 (Reynolds Risk) to 0.70 (*Framingham 1998, ASCVD*) for Stroke outcome; from 0.61 to 0.80 (*UKPDS 82 C-HF*) for AF outcome; from 0.60 (*Reynolds Risk*) to 0.78 (RECODE) for HF outcome.

**Appendix Figure 12: Forest plot presenting discrimination of the risk prediction models against CVD, CHD and CVD + AF + HF outcomes. The figure displays the C-statistic and 95% CI. The C-statistic was calculated based on a complete-case analysis.**

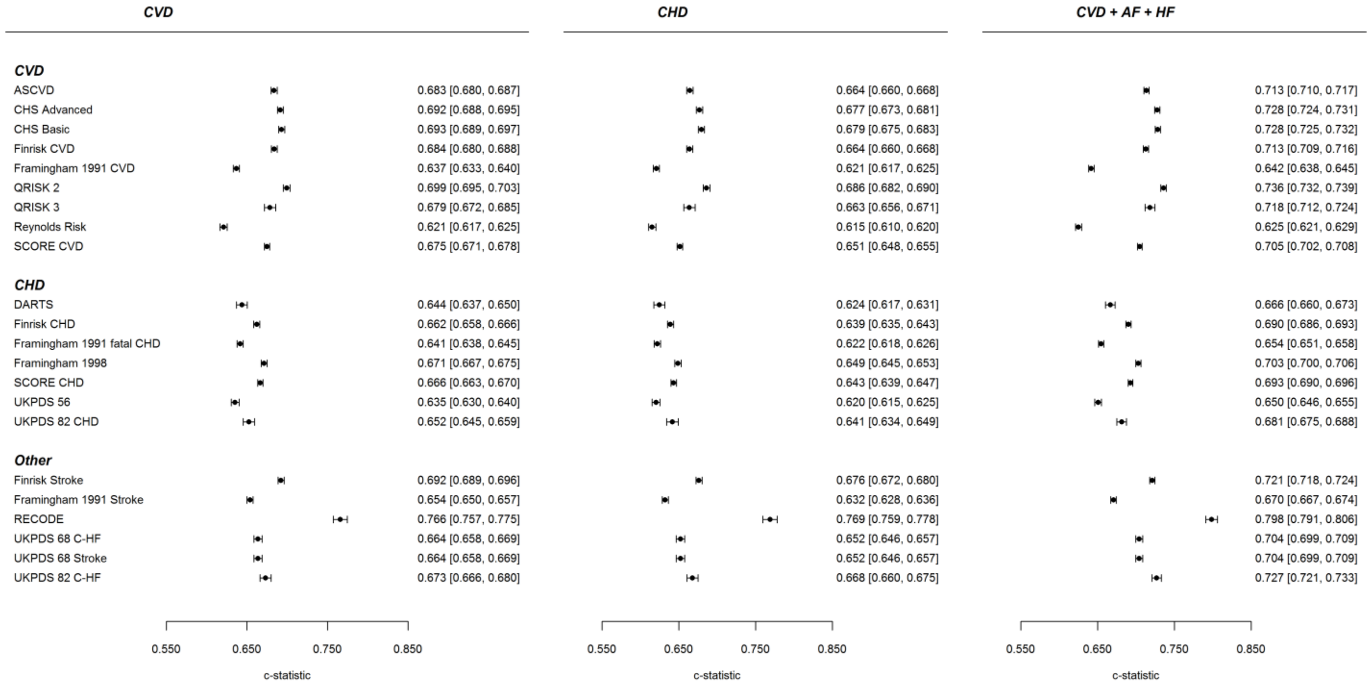

**Appendix Figure 13: Forest plot presenting discrimination of the risk prediction models against Stroke, AF and HF outcomes. The figure displays the C-statistic and 95% CI. The C-statistic was calculated based on a complete-case analysis.**

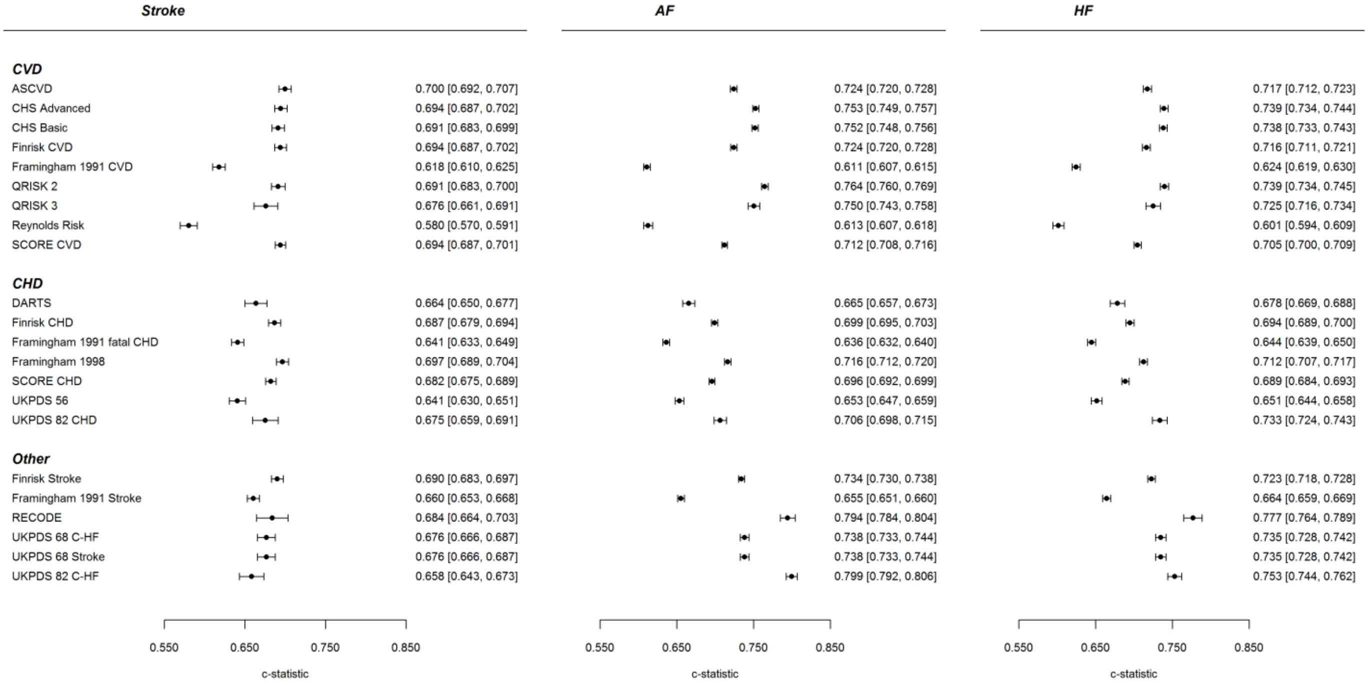

**Appendix Figure 14: Forest plot presenting discrimination of the risk prediction models against CVD, CHD and CVD + AF + HF outcomes. The figure displays the C-statistic and 95% CI. The C-statistic was calculated accounting for missing data using multiple imputation.**

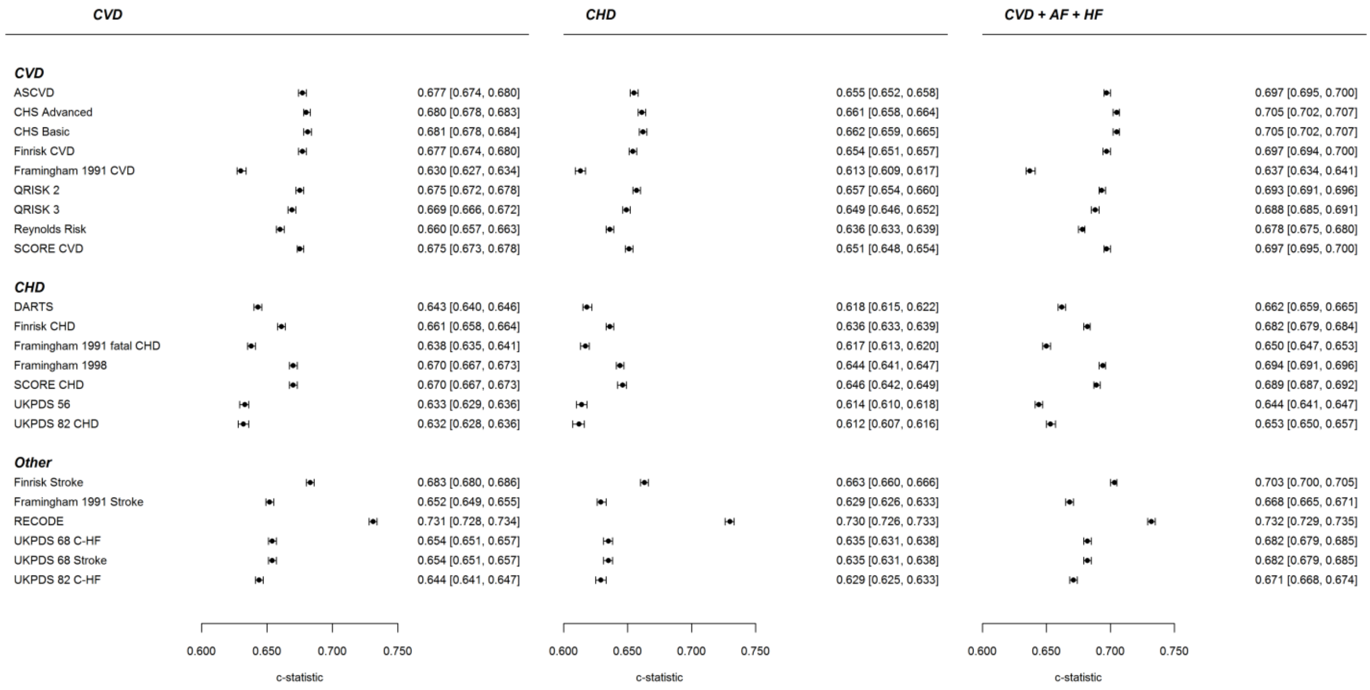

**Appendix Figure 15: Forest plot presenting discrimination of the risk prediction models against Stroke, AF and HF outcomes. The figure displays the C-statistic and 95% CI. The C-statistic was calculated accounting for missing data using multiple imputation.**

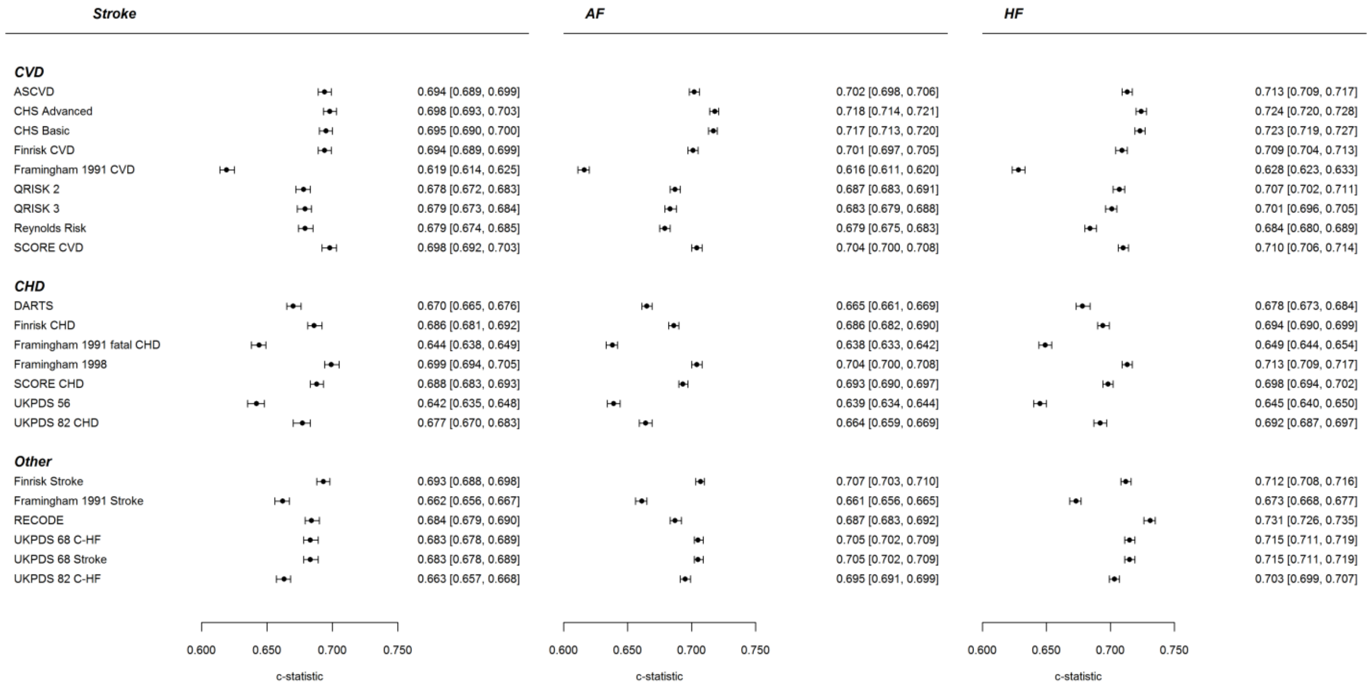

**Appendix Table 12: Calibration (calibration-in-the-large (CIL) and calibration slope (CS)) for all included risk score against CVD, CHD, CVD + AF + HF, Stroke, AF and HF outcomes based on a complete-case analysis.**

| | CVD | | CHD | | CVD + AF + HF | | Stroke | | AF | | HF | |
|---|---|---|---|---|---|---|---|---|---|---|---|---|
| | CS | CIL | CS | CIL | CS | CIL | CS | CIL | CS | CIL | CS | CIL |
| ASCVD | 0.471 (0.459;0.483) | -0.182 (-0.199;-0.165) | 0.399 (0.387;0.411) | -0.536 (-0.554;-0.518) | 0.599 (0.587;0.612) | 0.318 (0.302;0.334) | 0.424 (0.404;0.444) | -2.703 (-2.736;-2.669) | 0.573 (0.558;0.587) | -0.996 (-1.016;-0.977) | 0.515 (0.499;0.531) | -1.696 (-1.720;-1.672) |
| CHS Advanced | 0.772 (0.754;0.790) | -0.108 (-0.124;-0.092) | 0.697 (0.679;0.716) | -0.420 (-0.437;-0.403) | 0.974 (0.956;0.992) | 0.329 (0.315;0.344) | 0.740 (0.705;0.774) | -2.324 (-2.355;-2.292) | 1.054 (1.031;1.077) | -0.851 (-0.870;-0.833) | 0.960 (0.934;0.987) | -1.458 (-1.480;-1.435) |
| CHS Basic | 0.911 (0.890;0.932) | -0.177 (-0.192;-0.161) | 0.828 (0.806;0.850) | -0.478 (-0.494;-0.461) | 1.141 (1.119;1.162) | 0.245 (0.230;0.259) | 0.857 (0.816;0.897) | -2.336 (-2.367;-2.305) | 1.233 (1.206;1.260) | -0.896 (-0.914;-0.878) | 1.125 (1.094;1.156) | -1.487 (-1.509;-1.464) |
| Finrisk CVD | 0.613 (0.598;0.627) | 0.933 (0.917;0.949) | 0.536 (0.521;0.551) | 0.610 (0.593;0.627) | 0.734 (0.720;0.748) | 1.394 (1.379;1.409) | 0.632 (0.603;0.661) | -1.360 (-1.391;-1.329) | 0.763 (0.745;0.781) | 0.155 (0.136;0.173) | 0.726 (0.704;0.747) | -0.477 (-0.499;-0.454) |
| Framingham 1991 CVD | 0.576 (0.558;0.594) | -1.314 (-1.329;-1.298) | 0.499 (0.481;0.518) | -1.616 (-1.632;-1.600) | 0.598 (0.581;0.615) | -0.887 (-0.901;-0.873) | 0.488 (0.452;0.524) | -3.507 (-3.538;-3.477) | 0.447 (0.427;0.468) | -2.048 (-2.066;-2.029) | 0.506 (0.480;0.531) | -2.645 (-2.667;-2.623) |
| QRISK 2 | 0.798 (0.777;0.818) | -0.228 (-0.246;-0.211) | 0.721 (0.700;0.742) | -0.522 (-0.541;-0.504) | 1.042 (1.021;1.064) | 0.190 (0.174;0.206) | 0.624 (0.588;0.660) | -2.506 (-2.541;-2.470) | 1.154 (1.126;1.181) | -0.997 (-1.017;-0.976) | 0.915 (0.887;0.944) | -1.591 (-1.616;-1.566) |
| QRISK 3 | 0.723 (0.688;0.758) | -0.226 (-0.255;-0.197) | 0.642 (0.607;0.678) | -0.508 (-0.538;-0.477) | 0.959 (0.923;0.995) | 0.198 (0.171;0.225) | 0.582 (0.518;0.647) | -2.559 (-2.620;-2.497) | 1.084 (1.038;1.129) | -0.962 (-0.996;-0.927) | 0.869 (0.819;0.919) | -1.598 (-1.640;-1.556) |
| Reynolds Risk | 0.096 (0.091;0.101) | 1.482 (1.464;1.501) | 0.098 (0.093;0.104) | 1.122 (1.103;1.141) | 0.089 (0.084;0.093) | 2.027 (2.009;2.046) | 0.032 (0.022;0.043) | -1.030 (-1.065;-0.995) | 0.069 (0.063;0.075) | 0.580 (0.559;0.601) | 0.059 (0.051;0.066) | -0.077 (-0.102;-0.052) |
| SCORE CVD | 0.486 (0.475;0.497) | 2.217 (2.203;2.232) | 0.417 (0.405;0.428) | 1.872 (1.856;1.887) | 0.577 (0.566;0.588) | 2.695 (2.681;2.709) | 0.546 (0.522;0.569) | -0.106 (-0.133;-0.078) | 0.618 (0.604;0.633) | 1.386 (1.369;1.403) | 0.591 (0.573;0.608) | 0.775 (0.755;0.795) |
| DARTS | 0.578 (0.547;0.610) | -0.411 (-0.439;-0.384) | 0.491 (0.458;0.523) | -0.706 (-0.736;-0.677) | 0.694 (0.663;0.724) | 0.005 (-0.021;0.030) | 0.585 (0.526;0.644) | -2.628 (-2.684;-2.572) | 0.628 (0.591;0.666) | -1.200 (-1.234;-1.167) | 0.657 (0.614;0.701) | -1.736 (-1.776;-1.697) |
| Finrisk CHD | 0.531 (0.517;0.546) | 0.974 (0.958;0.989) | 0.448 (0.433;0.462) | 0.651 (0.635;0.668) | 0.639 (0.625;0.653) | 1.432 (1.417;1.447) | 0.606 (0.577;0.635) | -1.317 (-1.349;-1.286) | 0.661 (0.643;0.679) | 0.198 (0.179;0.216) | 0.640 (0.619;0.661) | -0.433 (-0.456;-0.411) |
| Framingham 1991 | 0.576 | 0.949 | 0.498 | 0.649 | 0.621 | 1.376 | 0.592 | -1.222 (-1.252;- | 0.565 | 0.222 | 0.606 | -0.368 (-0.390;- |

| | | | | | | | | | | | | |
|---|---|---|---|---|---|---|---|---|---|---|---|---|
| fatal CHD | (0.558;0.593) | (0.934;0.964) | (0.480;0.517) | (0.633;0.665) | (0.605;0.637) | (1.362;1.391) | (0.554;0.631) | 1.191) | (0.543;0.586) | (0.204;0.240) | (0.579;0.634) | 0.346) |
| Framingham 1998 | 0.845 (0.823;0.866) | 0.366 (0.351;0.381) | 0.717 (0.695;0.739) | 0.072 (0.056;0.088) | 1.039 (1.018;1.060) | 0.781 (0.767;0.795) | 0.949 (0.906;0.992) | -1.787 (-1.817;-1.756) | 1.079 (1.052;1.105) | -0.350 (-0.368;-0.332) | 1.048 (1.016;1.080) | -0.935 (-0.957;-0.913) |
| SCORE CHD | 0.490 (0.478;0.501) | 2.847 (2.833;2.861) | 0.420 (0.408;0.432) | 2.510 (2.494;2.525) | 0.569 (0.557;0.580) | 3.313 (3.300;3.327) | 0.546 (0.521;0.571) | 0.570 (0.543;0.597) | 0.600 (0.585;0.616) | 2.035 (2.018;2.052) | 0.576 (0.558;0.595) | 1.436 (1.416;1.456) |
| UKPDS 56 | 0.469 (0.451;0.488) | 0.205 (0.186;0.225) | 0.408 (0.388;0.427) | -0.102 (-0.123;-0.081) | 0.541 (0.523;0.559) | 0.657 (0.638;0.675) | 0.436 (0.401;0.471) | -2.081 (-2.121;-2.041) | 0.511 (0.489;0.533) | -0.542 (-0.565;-0.518) | 0.494 (0.468;0.520) | -1.162 (-1.191;-1.134) |
| UKPDS 82 CHD | 0.504 (0.480;0.529) | 0.035 (0.006;0.064) | 0.454 (0.428;0.479) | -0.266 (-0.297;-0.236) | 0.653 (0.628;0.679) | 0.498 (0.471;0.525) | 0.458 (0.416;0.500) | -2.471 (-2.533;-2.409) | 0.665 (0.636;0.695) | -0.749 (-0.784;-0.715) | 0.713 (0.679;0.748) | -1.393 (-1.435;-1.351) |
| Finrisk Stroke | 0.583 (0.570;0.596) | 0.924 (0.908;0.940) | 0.522 (0.509;0.536) | 0.590 (0.572;0.607) | 0.704 (0.691;0.717) | 1.399 (1.384;1.414) | 0.536 (0.511;0.561) | -1.454 (-1.486;-1.423) | 0.712 (0.696;0.729) | 0.117 (0.098;0.136) | 0.658 (0.639;0.677) | -0.540 (-0.563;-0.517) |
| Framingham 1991 Stroke | 0.423 (0.411;0.435) | 0.413 (0.397;0.429) | 0.350 (0.337;0.362) | 0.075 (0.058;0.092) | 0.484 (0.472;0.496) | 0.890 (0.875;0.905) | 0.389 (0.368;0.411) | -2.051 (-2.084;-2.018) | 0.395 (0.381;0.409) | -0.410 (-0.429;-0.390) | 0.412 (0.396;0.429) | -1.081 (-1.104;-1.057) |
| RECODE | 1.068 (1.020;1.116) | -0.075 (-0.115;-0.035) | 1.031 (0.981;1.080) | -0.395 (-0.438;-0.352) | 1.426 (1.372;1.480) | 0.418 (0.381;0.455) | 0.513 (0.442;0.584) | -2.331 (-2.411;-2.251) | 1.183 (1.128;1.238) | -0.750 (-0.797;-0.704) | 0.983 (0.925;1.041) | -1.401 (-1.457;-1.344) |
| UKPDS 68 C-HF | 0.490 (0.473;0.508) | 1.371 (1.350;1.393) | 0.450 (0.431;0.468) | 1.062 (1.038;1.085) | 0.642 (0.624;0.659) | 1.836 (1.815;1.857) | 0.503 (0.467;0.540) | -1.004 (-1.048;-0.959) | 0.749 (0.725;0.773) | 0.563 (0.537;0.589) | 0.731 (0.703;0.759) | -0.048 (-0.080;-0.017) |
| UKPDS 68 Stroke | 0.490 (0.473;0.508) | 1.371 (1.350;1.393) | 0.450 (0.431;0.468) | 1.062 (1.038;1.085) | 0.642 (0.624;0.659) | 1.836 (1.815;1.857) | 0.503 (0.467;0.540) | -1.004 (-1.048;-0.959) | 0.749 (0.725;0.773) | 0.563 (0.537;0.589) | 0.731 (0.703;0.759) | -0.048 (-0.080;-0.017) |
| UKPDS 82 C-HF | 0.480 (0.458;0.502) | 0.523 (0.494;0.552) | 0.451 (0.429;0.474) | 0.211 (0.180;0.242) | 0.783 (0.757;0.809) | 1.001 (0.973;1.028) | 0.347 (0.310;0.385) | -2.131 (-2.195;-2.066) | 1.045 (1.011;1.079) | -0.294 (-0.329;-0.258) | 0.644 (0.615;0.674) | -0.976 (-1.019;-0.932) |

**Appendix Table 13: Calibration after performing recalibration (calibration-in-the-large (CIL) and calibration slope (CS)) for all included risk score against CVD, CHD, CVD + AF + HF, Stroke, AF and HF outcomes based on a complete-case analysis.**

| | CVD | | CHD | | CVD + AF + HF | | Stroke | | AF | | HF | |
|---|---|---|---|---|---|---|---|---|---|---|---|---|
| | CS | CIL | CS | CIL | CS | CIL | CS | CIL | CS | CIL | CS | CIL |
| ASCVD | 0.988 (0.963;1.014) | -0.007 (-0.022;0.008) | 0.951 (0.922;0.980) | -0.016 (-0.033;-0.000) | 1.007 (0.986;1.028) | -0.004 (-0.019;0.011) | 0.987 (0.940;1.033) | -0.067 (-0.098;-0.036) | 0.998 (0.972;1.023) | 0.023 (0.005;0.042) | 0.942 (0.912;0.971) | -0.014 (-0.036;0.009) |
| CHS Advanced | 1.068 (1.043;1.092) | -0.006 (-0.022;0.009) | 1.052 (1.024;1.080) | -0.028 (-0.044;-0.012) | 1.032 (1.013;1.052) | -0.035 (-0.050;-0.020) | 1.031 (0.983;1.079) | 0.008 (-0.023;0.039) | 1.000 (0.978;1.022) | -0.033 (-0.052;-0.014) | 1.060 (1.031;1.089) | -0.079 (-0.102;-0.057) |
| CHS Basic | 1.064 (1.040;1.089) | -0.006 (-0.022;0.009) | 1.050 (1.022;1.078) | -0.028 (-0.044;-0.012) | 1.029 (1.010;1.048) | -0.035 (-0.050;-0.020) | 1.030 (0.981;1.079) | 0.007 (-0.024;0.038) | 0.996 (0.974;1.017) | -0.033 (-0.052;-0.014) | 1.056 (1.027;1.085) | -0.079 (-0.102;-0.057) |
| Finrisk CVD | 0.974 (0.951;0.997) | -0.004 (-0.019;0.011) | 0.971 (0.944;0.998) | 0.003 (-0.013;0.019) | 1.001 (0.981;1.020) | 0.014 (0.000;0.029) | 0.944 (0.901;0.988) | -0.020 (-0.051;0.011) | 1.024 (0.999;1.048) | 0.042 (0.024;0.060) | 1.054 (1.022;1.085) | -0.033 (-0.055;-0.011) |
| Framingham 1991 CVD | 1.072 (1.038;1.105) | -0.015 (-0.029;-0.000) | 1.138 (1.096;1.181) | -0.004 (-0.019;0.012) | 1.064 (1.034;1.094) | -0.008 (-0.022;0.005) | 1.037 (0.961;1.114) | -0.081 (-0.112;-0.051) | 1.030 (0.982;1.078) | 0.005 (-0.012;0.023) | 1.117 (1.061;1.174) | -0.014 (-0.036;0.007) |
| QRISK 2 | 0.976 (0.951;1.001) | -0.039 (-0.056;-0.023) | 0.939 (0.912;0.966) | -0.041 (-0.058;-0.023) | 1.002 (0.981;1.023) | -0.022 (-0.039;-0.006) | 1.011 (0.952;1.069) | -0.023 (-0.057;0.012) | 1.009 (0.986;1.033) | 0.045 (0.024;0.066) | 0.965 (0.934;0.995) | 0.078 (0.053;0.103) |
| QRISK 3 | 0.938 (0.893;0.983) | -0.014 (-0.042;0.014) | 0.995 (0.940;1.050) | -0.007 (-0.036;0.023) | 0.965 (0.929;1.002) | 0.022 (-0.005;0.050) | 1.264 (1.123;1.404) | -0.152 (-0.211;-0.092) | 1.015 (0.972;1.058) | 0.054 (0.020;0.089) | 1.017 (0.958;1.075) | 0.035 (-0.007;0.077) |
| Reynolds Risk | 0.968 (0.918;1.019) | -0.032 (-0.048;-0.017) | 1.007 (0.952;1.063) | -0.033 (-0.050;-0.016) | 0.924 (0.876;0.972) | -0.015 (-0.030;-0.000) | 0.599 (0.407;0.791) | -0.030 (-0.063;0.004) | 0.893 (0.814;0.972) | -0.007 (-0.026;0.012) | 0.788 (0.688;0.887) | 0.052 (0.029;0.075) |
| SCORE CVD | 1.091 (1.066;1.116) | -0.001 (-0.015;0.012) | 1.150 (1.118;1.181) | -0.010 (-0.024;0.004) | 1.069 (1.049;1.089) | -0.016 (-0.029;-0.004) | 0.963 (0.921;1.004) | 0.102 (0.075;0.128) | 1.074 (1.049;1.100) | -0.050 (-0.066;-0.034) | 1.062 (1.030;1.093) | -0.020 (-0.040;-0.001) |
| DARTS | 0.913 (0.863;0.962) | 0.024 (-0.002;0.051) | 0.905 (0.845;0.965) | 0.002 (-0.026;0.030) | 0.951 (0.908;0.993) | 0.000 (-0.025;0.025) | 1.069 (0.962;1.176) | 0.030 (-0.025;0.085) | 1.008 (0.948;1.068) | -0.046 (-0.078;-0.013) | 0.937 (0.874;0.999) | 0.063 (0.025;0.102) |
| Finrisk CHD | 0.967 (0.941;0.993) | -0.004 (-0.019;0.011) | 0.959 (0.928;0.990) | 0.003 (-0.013;0.018) | 1.001 (0.979;1.023) | 0.014 (-0.000;0.028) | 0.937 (0.892;0.982) | -0.021 (-0.052;0.009) | 1.012 (0.985;1.039) | 0.040 (0.022;0.058) | 1.044 (1.009;1.078) | -0.034 (-0.055;-0.012) |
| Framingham 1991 fatal CHD | 1.050 (1.018;1.082) | -0.014 (-0.029;0.000) | 1.099 (1.059;1.139) | -0.004 (-0.019;0.012) | 1.058 (1.030;1.086) | -0.008 (-0.022;0.006) | 1.026 (0.959;1.093) | -0.081 (-0.112;-0.051) | 1.039 (0.999;1.078) | 0.006 (-0.012;0.023) | 1.106 (1.056;1.155) | -0.014 (-0.036;0.007) |

| | | | | | | | | | | | | |
|---|---|---|---|---|---|---|---|---|---|---|---|---|
| Framingham 1998 | 1.034 (1.007;1.060) | -0.005 (-0.020;0.010) | 1.048 (1.016;1.081) | 0.005 (-0.011;0.021) | 1.051 (1.030;1.073) | 0.003 (-0.011;0.018) | 0.942 (0.899;0.985) | -0.069 (-0.099;-0.038) | 0.990 (0.966;1.015) | 0.020 (0.002;0.038) | 1.046 (1.014;1.078) | -0.001 (-0.023;0.021) |
| SCORE CHD | 1.085 (1.059;1.111) | -0.002 (-0.016;0.011) | 1.151 (1.118;1.185) | -0.011 (-0.025;0.003) | 1.061 (1.040;1.082) | -0.018 (-0.031;-0.005) | 0.927 (0.884;0.970) | 0.100 (0.073;0.127) | 1.070 (1.043;1.097) | -0.052 (-0.068;-0.036) | 1.040 (1.007;1.073) | -0.022 (-0.041;-0.003) |
| UKPDS 56 | 1.001 (0.961;1.040) | 0.040 (0.022;0.059) | 0.976 (0.929;1.023) | 0.034 (0.015;0.054) | 0.967 (0.935;0.999) | 0.037 (0.020;0.054) | 1.078 (0.990;1.165) | 0.022 (-0.017;0.060) | 0.970 (0.928;1.012) | 0.036 (0.013;0.058) | 0.980 (0.929;1.032) | -0.011 (-0.039;0.016) |
| UKPDS 82 CHD | 1.111 (1.057;1.166) | -0.050 (-0.077;-0.024) | 1.073 (1.013;1.133) | -0.031 (-0.060;-0.003) | 1.079 (1.038;1.121) | -0.038 (-0.063;-0.012) | 1.451 (1.318;1.584) | -0.118 (-0.176;-0.060) | 1.005 (0.961;1.050) | -0.042 (-0.075;-0.009) | 1.034 (0.984;1.083) | 0.010 (-0.030;0.050) |
| Finrisk Stroke | 0.975 (0.953;0.998) | -0.003 (-0.018;0.012) | 0.977 (0.951;1.003) | 0.004 (-0.012;0.020) | 0.996 (0.977;1.015) | 0.015 (0.001;0.029) | 0.949 (0.904;0.994) | -0.020 (-0.051;0.010) | 1.026 (1.003;1.050) | 0.042 (0.024;0.060) | 1.054 (1.024;1.085) | -0.033 (-0.055;-0.011) |
| Framingham 1991 Stroke | 1.073 (1.042;1.104) | -0.011 (-0.026;0.004) | 1.101 (1.062;1.140) | -0.001 (-0.016;0.015) | 1.061 (1.035;1.087) | -0.004 (-0.018;0.009) | 1.060 (1.001;1.119) | -0.076 (-0.106;-0.045) | 0.995 (0.960;1.030) | 0.010 (-0.008;0.028) | 1.078 (1.035;1.120) | -0.010 (-0.031;0.012) |
| RECODE | 0.974 (0.930;1.018) | 0.063 (0.023;0.104) | 0.972 (0.926;1.019) | -0.001 (-0.045;0.042) | 1.047 (1.007;1.086) | 0.079 (0.040;0.118) | 0.786 (0.677;0.894) | 0.171 (0.093;0.249) | 0.968 (0.923;1.013) | 0.041 (-0.007;0.090) | 0.859 (0.808;0.910) | -0.145 (-0.203;-0.087) |
| UKPDS 68 C-HF | 1.052 (1.014;1.090) | 0.009 (-0.011;0.030) | 1.093 (1.048;1.138) | 0.010 (-0.011;0.032) | 1.042 (1.013;1.071) | 0.021 (0.002;0.040) | 0.955 (0.885;1.024) | -0.012 (-0.055;0.031) | 0.966 (0.935;0.996) | 0.028 (0.002;0.053) | 0.939 (0.903;0.975) | 0.040 (0.009;0.070) |
| UKPDS 68 Stroke | 1.052 (1.014;1.090) | 0.009 (-0.011;0.030) | 1.093 (1.048;1.138) | 0.010 (-0.011;0.032) | 1.042 (1.013;1.071) | 0.021 (0.002;0.040) | 0.955 (0.885;1.024) | -0.012 (-0.055;0.031) | 0.966 (0.935;0.996) | 0.028 (0.002;0.053) | 0.939 (0.903;0.975) | 0.040 (0.009;0.070) |
| UKPDS 82 C-HF | 1.048 (0.999;1.096) | -0.045 (-0.072;-0.018) | 1.017 (0.966;1.068) | -0.026 (-0.054;0.003) | 1.048 (1.013;1.082) | -0.027 (-0.053;-0.001) | 1.090 (0.972;1.207) | -0.120 (-0.178;-0.062) | 1.089 (1.054;1.124) | -0.025 (-0.060;0.011) | 1.066 (1.017;1.115) | 0.015 (-0.025;0.056) |

**Appendix Table 14 Calibration (calibration-in-the-large (CIL) and calibration slope (CS)) for all included risk score against CVD, CHD, CVD + AF + HF, Stroke, AF and HF outcomes based on imputed dataset.**

| Risk Score Name | **CVD** | | **CHD** | | **CVD + AF + HF** | | **Stroke** | | **AF** | | **HF** | |
|---|---|---|---|---|---|---|---|---|---|---|---|---|
| | **CS** | **CIL** | **CS** | **CIL** | **CS** | **CIL** | **CS** | **CIL** | **CS** | **CIL** | **CS** | **CIL** |
| ASCVD | 0.411 (0.402;0.419) | -0.153 (-0.167;-0.139) | 0.338 (0.33;0.347) | -0.573 (-0.588;-0.558) | 0.489 (0.48;0.497) | 0.213 (0.199;0.227) | 0.378 (0.364;0.391) | -2.49 (-2.514;-2.466) | 0.425 (0.413;0.436) | -1.587 (-1.605;-1.569) | 0.448 (0.436;0.46) | -1.846 (-1.866;-1.827) |
| CHS Advanced | 0.677 (0.664;0.689) | -0.087 (-0.099;-0.075) | 0.586 (0.574;0.599) | -0.457 (-0.47;-0.444) | 0.799 (0.787;0.811) | 0.237 (0.226;0.248) | 0.707 (0.685;0.729) | -2.131 (-2.153;-2.11) | 0.797 (0.779;0.814) | -1.349 (-1.366;-1.333) | 0.833 (0.814;0.852) | -1.575 (-1.593;-1.558) |
| CHS Basic | 0.8 (0.786;0.815) | -0.165 (-0.176;-0.153) | 0.699 (0.684;0.714) | -0.522 (-0.534;-0.509) | 0.94 (0.926;0.954) | 0.146 (0.135;0.158) | 0.828 (0.801;0.854) | -2.15 (-2.172;-2.129) | 0.94 (0.919;0.96) | -1.386 (-1.402;-1.37) | 0.982 (0.959;1.004) | -1.606 (-1.624;-1.589) |
| Finrisk CVD | 0.543 (0.532;0.553) | 0.913 (0.899;0.927) | 0.461 (0.45;0.471) | 0.521 (0.506;0.535) | 0.619 (0.609;0.629) | 1.258 (1.245;1.271) | 0.584 (0.565;0.604) | -1.215 (-1.238;-1.192) | 0.61 (0.595;0.625) | -0.411 (-0.429;-0.393) | 0.641 (0.624;0.658) | -0.644 (-0.663;-0.625) |
| Framingham 1991 CVD | 0.514 (0.499;0.529) | -1.379 (-1.394;-1.364) | 0.441 (0.426;0.456) | -1.743 (-1.759;-1.727) | 0.543 (0.529;0.557) | -1.061 (-1.075;-1.046) | 0.463 (0.438;0.487) | -3.393 (-3.416;-3.369) | 0.445 (0.425;0.465) | -2.621 (-2.64;-2.601) | 0.492 (0.469;0.514) | -2.844 (-2.864;-2.823) |
| QRISK 2 | 0.666 (0.652;0.681) | -0.174 (-0.187;-0.161) | 0.579 (0.563;0.596) | -0.538 (-0.551;-0.524) | 0.757 (0.744;0.771) | 0.144 (0.131;0.157) | 0.595 (0.562;0.629) | -2.196 (-2.218;-2.174) | 0.665 (0.642;0.688) | -1.418 (-1.435;-1.4) | 0.742 (0.713;0.77) | -1.642 (-1.66;-1.623) |
| QRISK 3 | 0.682 (0.667;0.698) | 0.04 (0.028;0.053) | 0.582 (0.565;0.6) | -0.319 (-0.332;-0.305) | 0.777 (0.763;0.792) | 0.354 (0.341;0.366) | 0.637 (0.6;0.673) | -1.961 (-1.983;-1.939) | 0.689 (0.664;0.713) | -1.189 (-1.206;-1.172) | 0.755 (0.725;0.785) | -1.411 (-1.429;-1.393) |
| Reynolds Risk | 0.466 (0.455;0.476) | 0.917 (0.902;0.932) | 0.386 (0.375;0.397) | 0.524 (0.509;0.54) | 0.53 (0.52;0.539) | 1.262 (1.248;1.276) | 0.505 (0.487;0.524) | -1.233 (-1.256;-1.209) | 0.507 (0.492;0.521) | -0.415 (-0.433;-0.396) | 0.526 (0.51;0.542) | -0.651 (-0.671;-0.631) |
| SCORE CVD | 0.434 (0.426;0.443) | 2.147 (2.133;2.16) | 0.369 (0.361;0.378) | 1.74 (1.726;1.755) | 0.492 (0.484;0.5) | 2.509 (2.496;2.522) | 0.503 (0.485;0.52) | -0.034 (-0.057;-0.011) | 0.523 (0.51;0.535) | 0.784 (0.767;0.802) | 0.545 (0.531;0.559) | 0.546 (0.528;0.565) |
| DARTS | 0.495 (0.481;0.509) | -0.532 (-0.546;-0.518) | 0.389 (0.373;0.405) | -0.902 (-0.916;-0.887) | 0.588 (0.574;0.602) | -0.211 (-0.224;-0.197) | 0.488 (0.451;0.526) | -2.606 (-2.631;-2.582) | 0.505 (0.482;0.529) | -1.803 (-1.822;-1.785) | 0.541 (0.509;0.573) | -2.034 (-2.054;-2.014) |
| Finrisk CHD | 0.476 (0.466;0.486) | 0.913 (0.898;0.928) | 0.394 (0.383;0.404) | 0.52 (0.504;0.535) | 0.547 (0.537;0.557) | 1.259 (1.244;1.273) | 0.545 (0.526;0.564) | -1.225 (-1.248;-1.201) | 0.543 (0.529;0.557) | -0.416 (-0.435;-0.397) | 0.573 (0.556;0.589) | -0.651 (-0.67;-0.631) |
| Framingham 1991 | 0.498 | 0.893 | 0.426 | 0.531 | 0.538 | 1.212 | 0.54 | -1.1 (-1.123;- | 0.519 | -0.337 (-0.355;- | 0.562 | -0.557 (-0.577;- |

| | | | | | | | | | | | | |
|---|---|---|---|---|---|---|---|---|---|---|---|---|
| fatal CHD | (0.485;0.511) | (0.877;0.909) | (0.412;0.439) | (0.515;0.547) | (0.526;0.55) | (1.197;1.227) | (0.513;0.567) | 1.076) | (0.499;0.539) | 0.318) | (0.539;0.585) | 0.537) |
| Framingham 1998 | 0.773 (0.758;0.788) | 0.319 (0.306;0.331) | 0.638 (0.622;0.654) | -0.035 (-0.048;-0.022) | 0.909 (0.894;0.924) | 0.628 (0.616;0.639) | 0.889 (0.86;0.917) | -1.655 (-1.677;-1.633) | 0.912 (0.89;0.934) | -0.894 (-0.91;-0.878) | 0.968 (0.945;0.992) | -1.113 (-1.131;-1.095) |
| SCORE CHD | 0.444 (0.435;0.452) | 2.763 (2.749;2.776) | 0.38 (0.37;0.389) | 2.366 (2.352;2.381) | 0.496 (0.487;0.504) | 3.115 (3.102;3.129) | 0.508 (0.489;0.526) | 0.63 (0.607;0.653) | 0.526 (0.512;0.54) | 1.432 (1.415;1.45) | 0.544 (0.529;0.56) | 1.199 (1.181;1.218) |
| UKPDS 56 | 0.433 (0.421;0.444) | 0.146 (0.129;0.163) | 0.36 (0.348;0.372) | -0.235 (-0.253;-0.218) | 0.483 (0.471;0.494) | 0.478 (0.463;0.494) | 0.422 (0.401;0.442) | -1.975 (-2.002;-1.948) | 0.426 (0.409;0.443) | -1.159 (-1.18;-1.138) | 0.443 (0.425;0.462) | -1.395 (-1.417;-1.372) |
| UKPDS 82 CHD | 0.425 (0.412;0.438) | 0.041 (0.028;0.054) | 0.349 (0.335;0.362) | -0.349 (-0.363;-0.334) | 0.515 (0.502;0.528) | 0.379 (0.367;0.392) | 0.492 (0.474;0.509) | -2.136 (-2.16;-2.113) | 0.461 (0.446;0.475) | -1.296 (-1.314;-1.278) | 0.578 (0.562;0.593) | -1.538 (-1.557;-1.519) |
| Finrisk Stroke | 0.522 (0.513;0.532) | 0.944 (0.931;0.957) | 0.452 (0.442;0.462) | 0.541 (0.527;0.555) | 0.596 (0.587;0.605) | 1.297 (1.284;1.309) | 0.517 (0.5;0.535) | -1.253 (-1.275;-1.23) | 0.568 (0.555;0.582) | -0.422 (-0.439;-0.404) | 0.589 (0.575;0.604) | -0.663 (-0.682;-0.644) |
| Framingham 1991 Stroke | 0.383 (0.372;0.394) | 0.364 (0.347;0.38) | 0.312 (0.302;0.323) | -0.049 (-0.067;-0.031) | 0.436 (0.425;0.446) | 0.724 (0.708;0.739) | 0.355 (0.339;0.371) | -1.932 (-1.961;-1.903) | 0.363 (0.351;0.376) | -1.049 (-1.071;-1.027) | 0.39 (0.376;0.404) | -1.303 (-1.327;-1.279) |
| RECODE | 1.052 (1.034;1.07) | 0.078 (0.066;0.09) | 1.001 (0.983;1.02) | -0.284 (-0.297;-0.271) | 1.098 (1.081;1.115) | 0.392 (0.381;0.403) | 0.6 (0.574;0.625) | -1.956 (-1.978;-1.933) | 0.672 (0.651;0.692) | -1.169 (-1.185;-1.152) | 0.875 (0.852;0.899) | -1.395 (-1.413;-1.377) |
| UKPDS 68 C-HF | 0.424 (0.412;0.435) | 1.371 (1.351;1.391) | 0.361 (0.35;0.372) | 0.967 (0.945;0.989) | 0.516 (0.504;0.529) | 1.726 (1.707;1.744) | 0.486 (0.468;0.504) | -0.824 (-0.859;-0.789) | 0.559 (0.542;0.576) | 0.006 (-0.022;0.034) | 0.592 (0.575;0.61) | -0.235 (-0.264;-0.205) |
| UKPDS 68 Stroke | 0.424 (0.412;0.435) | 1.371 (1.351;1.391) | 0.361 (0.35;0.372) | 0.967 (0.945;0.989) | 0.516 (0.504;0.529) | 1.726 (1.707;1.744) | 0.486 (0.468;0.504) | -0.824 (-0.859;-0.789) | 0.559 (0.542;0.576) | 0.006 (-0.022;0.034) | 0.592 (0.575;0.61) | -0.235 (-0.264;-0.205) |
| UKPDS 82 C-HF | 0.479 (0.458;0.499) | 0.745 (0.723;0.767) | 0.421 (0.401;0.441) | 0.368 (0.344;0.392) | 0.586 (0.564;0.608) | 1.075 (1.054;1.095) | 0.516 (0.487;0.544) | -1.33 (-1.37;-1.291) | 0.643 (0.612;0.675) | -0.537 (-0.569;-0.506) | 0.68 (0.645;0.714) | -0.766 (-0.799;-0.733) |

**Appendix Table 15 Calibration after performing recalibration (calibration-in-the-large (CIL) and calibration slope (CS)) for all included risk score against CVD, CHD, CVD + AF + HF, Stroke, AF and HF outcomes accounting for missing data using multiple imputation.**

| | CVD | | CHD | | CVD + AF + HF | | Stroke | | AF | | HF | |
|---|---|---|---|---|---|---|---|---|---|---|---|---|
| | CS | CIL | CS | CIL | CS | CIL | CS | CIL | CS | CIL | CS | CIL |
| ASCVD | 1.012 (0.987;1.036) | 0.006 (-0.005;0.017) | 0.985 (0.958;1.012) | 0.028 (0.016;0.04) | 0.99 (0.967;1.013) | -0.013 (-0.023;-0.002) | 1.003 (0.929;1.077) | 0.021 (0;0.043) | 0.979 (0.939;1.02) | -0.034 (-0.05;-0.018) | 0.955 (0.91;0.999) | -0.034 (-0.051;-0.016) |
| CHS Advanced | 1.006 (0.988;1.024) | 0.006 (-0.005;0.017) | 0.993 (0.971;1.015) | 0.028 (0.016;0.04) | 0.991 (0.976;1.006) | -0.013 (-0.023;-0.002) | 0.99 (0.958;1.022) | 0.021 (0;0.042) | 1.002 (0.979;1.024) | -0.034 (-0.05;-0.019) | 0.958 (0.936;0.981) | -0.034 (-0.051;-0.017) |
| CHS Basic | 1.006 (0.987;1.024) | 0.006 (-0.005;0.017) | 0.993 (0.971;1.015) | 0.028 (0.016;0.04) | 0.991 (0.976;1.007) | -0.013 (-0.024;-0.002) | 0.987 (0.955;1.02) | 0.021 (0;0.042) | 0.999 (0.976;1.022) | -0.034 (-0.05;-0.019) | 0.961 (0.936;0.985) | -0.034 (-0.052;-0.017) |
| Finrisk CVD | 1.014 (0.991;1.038) | 0.008 (-0.003;0.019) | 0.981 (0.953;1.009) | 0.03 (0.018;0.042) | 1 (0.979;1.021) | -0.011 (-0.022;0) | 1.022 (0.981;1.063) | 0.024 (0.003;0.045) | 0.983 (0.956;1.011) | -0.031 (-0.047;-0.015) | 0.986 (0.954;1.017) | -0.03 (-0.048;-0.013) |
| Framingham 1991 CVD | 0.985 (0.945;1.025) | 0.007 (-0.004;0.019) | 0.942 (0.9;0.984) | 0.029 (0.017;0.041) | 0.988 (0.952;1.023) | -0.011 (-0.022;0) | 1.099 (1.017;1.18) | 0.022 (0.001;0.043) | 0.952 (0.895;1.01) | -0.031 (-0.047;-0.015) | 1.014 (0.945;1.084) | -0.031 (-0.048;-0.014) |
| QRISK 2 | 0.988 (0.952;1.025) | 0.01 (-0.002;0.021) | 0.95 (0.903;0.997) | 0.032 (0.019;0.044) | 0.988 (0.958;1.017) | -0.009 (-0.02;0.002) | 1.031 (0.943;1.118) | 0.025 (0.004;0.047) | 0.984 (0.927;1.041) | -0.028 (-0.045;-0.012) | 0.969 (0.891;1.046) | -0.027 (-0.045;-0.009) |
| QRISK 3 | 0.983 (0.944;1.022) | 0.008 (-0.004;0.019) | 0.939 (0.887;0.991) | 0.029 (0.017;0.042) | 0.981 (0.948;1.014) | -0.011 (-0.022;0) | 1.022 (0.925;1.119) | 0.023 (0.001;0.044) | 0.974 (0.906;1.042) | -0.031 (-0.048;-0.015) | 0.953 (0.868;1.039) | -0.031 (-0.049;-0.012) |
| Reynolds Risk | 1.014 (0.981;1.047) | 0.008 (-0.003;0.02) | 0.973 (0.937;1.009) | 0.03 (0.018;0.042) | 0.996 (0.967;1.025) | -0.01 (-0.02;0.001) | 1.036 (0.981;1.092) | 0.025 (0.003;0.046) | 0.966 (0.934;0.997) | -0.03 (-0.046;-0.014) | 0.972 (0.923;1.022) | -0.029 (-0.046;-0.012) |
| SCORE CVD | 1.016 (0.99;1.041) | 0.009 (-0.002;0.02) | 0.975 (0.946;1.003) | 0.03 (0.018;0.042) | 1.001 (0.979;1.022) | -0.01 (-0.02;0.001) | 1.042 (1.001;1.084) | 0.027 (0.006;0.048) | 0.98 (0.95;1.009) | -0.031 (-0.047;-0.015) | 0.965 (0.933;0.996) | -0.03 (-0.047;-0.012) |
| DARTS | 1.017 (0.946;1.088) | 0.008 (-0.003;0.019) | 0.987 (0.887;1.087) | 0.03 (0.018;0.041) | 1.009 (0.954;1.064) | -0.01 (-0.02;0.001) | 1.085 (0.929;1.24) | 0.026 (0.004;0.048) | 1.021 (0.933;1.109) | -0.029 (-0.045;-0.013) | 0.99 (0.897;1.083) | -0.028 (-0.045;-0.01) |
| Finrisk CHD | 1.009 (0.976;1.043) | 0.009 (-0.002;0.02) | 0.971 (0.931;1.01) | 0.031 (0.019;0.043) | 0.997 (0.969;1.026) | -0.009 (-0.02;0.002) | 1.034 (0.987;1.081) | 0.026 (0.005;0.047) | 0.973 (0.938;1.008) | -0.029 (-0.045;-0.013) | 0.981 (0.939;1.023) | -0.028 (-0.045;-0.01) |
| Framingham 1991 fatal | 0.989 | 0.007 (- | 0.936 | 0.029 | 0.993 | -0.011 (-0.022;0) | 1.117 | 0.023 | 0.966 | -0.031 (-0.047;- | 0.985 | -0.03 (-0.048;- |

| | | | | | | | | | | | | |
|---|---|---|---|---|---|---|---|---|---|---|---|---|
| CHD | (0.948;1.03) | 0.004;0.018) | (0.888;0.984) | (0.017;0.041) | (0.958;1.029) | | (1.039;1.195) | (0.002;0.044) | (0.913;1.018) | 0.015) | (0.929;1.041) | 0.013) |
| Framingham 1998 | 1.003 (0.98;1.026) | 0.007 (-0.004;0.018) | 0.974 (0.948;1.001) | 0.029 (0.017;0.041) | 0.992 (0.974;1.01) | -0.011 (-0.022;0) | 1.009 (0.971;1.046) | 0.024 (0.003;0.045) | 0.999 (0.974;1.024) | -0.031 (-0.047;-0.015) | 0.954 (0.926;0.981) | -0.03 (-0.048;-0.013) |
| SCORE CHD | 1.023 (0.995;1.051) | 0.009 (-0.002;0.02) | 0.978 (0.946;1.01) | 0.03 (0.018;0.043) | 1.008 (0.983;1.032) | -0.01 (-0.02;0.001) | 1.064 (1.015;1.112) | 0.027 (0.006;0.048) | 0.972 (0.939;1.005) | -0.03 (-0.046;-0.014) | 0.978 (0.942;1.014) | -0.029 (-0.046;-0.012) |
| UKPDS 56 | 1.033 (0.982;1.084) | 0.008 (-0.003;0.019) | 0.992 (0.923;1.061) | 0.029 (0.017;0.041) | 1.016 (0.971;1.062) | -0.01 (-0.021;0.001) | 1.027 (0.952;1.102) | 0.023 (0.002;0.044) | 0.95 (0.903;0.997) | -0.031 (-0.046;-0.015) | 0.998 (0.92;1.077) | -0.03 (-0.047;-0.013) |
| UKPDS 82 CHD | 0.989 (0.958;1.02) | 0.006 (-0.005;0.017) | 0.984 (0.939;1.03) | 0.027 (0.016;0.039) | 0.984 (0.956;1.011) | -0.013 (-0.024;-0.002) | 0.953 (0.914;0.992) | 0.023 (0.002;0.044) | 1.011 (0.972;1.05) | -0.033 (-0.048;-0.017) | 0.925 (0.891;0.959) | -0.034 (-0.051;-0.016) |
| Finrisk Stroke | 1.019 (1;1.038) | 0.006 (-0.005;0.017) | 0.991 (0.968;1.013) | 0.028 (0.016;0.04) | 1.003 (0.985;1.021) | -0.013 (-0.023;-0.002) | 1.016 (0.978;1.053) | 0.022 (0;0.043) | 0.996 (0.972;1.02) | -0.034 (-0.049;-0.018) | 0.99 (0.964;1.017) | -0.033 (-0.05;-0.016) |
| Framingham 1991 Stroke | 0.99 (0.955;1.026) | 0.008 (-0.004;0.019) | 0.957 (0.921;0.993) | 0.029 (0.017;0.041) | 0.984 (0.952;1.016) | -0.01 (-0.022;0.001) | 1.031 (0.954;1.108) | 0.023 (0.001;0.044) | 0.978 (0.92;1.037) | -0.031 (-0.047;-0.015) | 0.974 (0.908;1.039) | -0.03 (-0.048;-0.013) |
| RECODE | 1.026 (1.004;1.049) | 0.003 (-0.01;0.015) | 1.047 (1.017;1.077) | 0.026 (0.013;0.04) | 1.015 (0.995;1.035) | -0.017 (-0.029;-0.005) | 1.062 (0.973;1.151) | 0.021 (0;0.043) | 1.041 (0.991;1.091) | -0.035 (-0.051;-0.018) | 0.997 (0.952;1.042) | -0.036 (-0.055;-0.018) |
| UKPDS 68 C-HF | 1.008 (0.971;1.045) | 0.006 (-0.005;0.017) | 0.995 (0.952;1.038) | 0.028 (0.016;0.04) | 0.994 (0.964;1.025) | -0.012 (-0.023;-0.001) | 0.988 (0.94;1.035) | 0.023 (0.001;0.044) | 0.998 (0.962;1.034) | -0.032 (-0.049;-0.016) | 0.945 (0.913;0.977) | -0.032 (-0.049;-0.014) |
| UKPDS 68 Stroke | 1.008 (0.971;1.045) | 0.006 (-0.005;0.017) | 0.995 (0.952;1.038) | 0.028 (0.016;0.04) | 0.994 (0.964;1.025) | -0.012 (-0.023;-0.001) | 0.988 (0.94;1.035) | 0.023 (0.001;0.044) | 0.998 (0.962;1.034) | -0.032 (-0.049;-0.016) | 0.945 (0.913;0.977) | -0.032 (-0.049;-0.014) |
| UKPDS 82 C-HF | 1.013 (0.973;1.053) | 0.006 (-0.005;0.017) | 1.003 (0.947;1.06) | 0.028 (0.016;0.04) | 0.994 (0.961;1.028) | -0.012 (-0.023;-0.001) | 0.997 (0.938;1.056) | 0.022 (0.001;0.043) | 0.993 (0.951;1.035) | -0.033 (-0.049;-0.016) | 0.947 (0.911;0.982) | -0.032 (-0.049;-0.014) |

**Subgroup analysis: gender (male vs female), age stratification (4 bins defined based on age distribution), statins usage at the baseline (used vs not used) and CVD history at the baseline (absent vs present).**

**Appendix Table 16 Discrimination (c-statistic) and calibration (calibration-in-the-large (CIL) and calibration slope (CS)) for all included risk scores against CVD and CVD + AF + HF outcomes for the imputed dataset stratified by gender (male vs female).**

| Risk Score Name | Gender (male vs female) | CVD | | | CVD + AF + HF | | |
|---|---|---|---|---|---|---|---|
| | | C-stat | CS | CIL | C-stat | CS | CIL |
| ASCVD | male | 0.665 (0.662;0.669) | 1.025 (0.986;1.064) | -0.002 (-0.016;0.013) | 0.686 (0.682;0.689) | 1.003 (0.97;1.037) | 0.007 (-0.007;0.021) |
| | female | 0.686 (0.682;0.69) | 0.975 (0.931;1.02) | 0.031 (0.014;0.049) | 0.708 (0.704;0.712) | 0.939 (0.903;0.975) | 0.024 (0.007;0.04) |
| Framingham 1991 fatal CHD | male | 0.629 (0.624;0.633) | 1.01 (0.95;1.07) | 0 (-0.015;0.014) | 0.641 (0.637;0.645) | 0.992 (0.94;1.043) | 0.008 (-0.006;0.021) |
| | female | 0.65 (0.645;0.655) | 1.06 (0.972;1.148) | 0.034 (0.016;0.051) | 0.662 (0.657;0.666) | 1.01 (0.936;1.083) | 0.026 (0.009;0.042) |
| Framingham 1991 CVD | male | 0.623 (0.618;0.627) | 1.022 (0.965;1.078) | -0.001 (-0.016;0.013) | 0.632 (0.628;0.637) | 1.001 (0.955;1.047) | 0.007 (-0.007;0.021) |
| | female | 0.631 (0.626;0.636) | 1.109 (1.02;1.198) | 0.032 (0.015;0.05) | 0.638 (0.633;0.643) | 1.06 (0.989;1.132) | 0.024 (0.008;0.04) |
| Framingham 1991 Stroke | male | 0.648 (0.644;0.652) | 1.05 (0.998;1.102) | -0.002 (-0.016;0.013) | 0.662 (0.658;0.666) | 1.029 (0.993;1.064) | 0.007 (-0.008;0.021) |
| | female | 0.67 (0.665;0.674) | 1.081 (0.991;1.171) | 0.036 (0.019;0.053) | 0.684 (0.679;0.688) | 1.047 (0.976;1.118) | 0.028 (0.012;0.045) |
| CHS Advanced | male | 0.677 (0.673;0.68) | 1.04 (1.016;1.065) | -0.005 (-0.02;0.009) | 0.699 (0.695;0.702) | 1.016 (0.995;1.037) | 0.003 (-0.011;0.017) |
| | female | 0.692 (0.687;0.696) | 0.984 (0.957;1.011) | 0.029 (0.012;0.046) | 0.716 (0.713;0.72) | 0.957 (0.935;0.979) | 0.02 (0.004;0.037) |
| CHS Basic | male | 0.675 (0.672;0.679) | 1.04 (1.015;1.065) | -0.005 (-0.02;0.009) | 0.697 (0.694;0.701) | 1.015 (0.994;1.037) | 0.003 (-0.011;0.017) |
| | female | 0.691 (0.687;0.695) | 0.981 (0.953;1.009) | 0.029 (0.012;0.046) | 0.716 (0.712;0.719) | 0.956 (0.934;0.979) | 0.02 (0.004;0.037) |
| DARTS | male | 0.633 (0.629;0.637) | 1.037 (0.926;1.148) | -0.002 (-0.017;0.012) | 0.65 (0.646;0.654) | 1.022 (0.929;1.114) | 0.006 (-0.008;0.02) |
| | female | 0.658 (0.654;0.663) | 1.046 (0.972;1.121) | 0.025 (0.007;0.042) | 0.678 (0.673;0.682) | 0.994 (0.937;1.052) | 0.016 (-0.001;0.033) |
| Finrisk CHD | male | 0.646 (0.642;0.649) | 1.039 (0.984;1.094) | -0.001 (-0.016;0.013) | 0.666 (0.662;0.669) | 1.019 (0.973;1.065) | 0.007 (-0.006;0.021) |
| | female | 0.679 (0.674;0.683) | 1.021 (0.965;1.076) | 0.03 (0.013;0.048) | 0.699 (0.694;0.703) | 0.974 (0.928;1.021) | 0.022 (0.005;0.039) |
| Finrisk CVD | male | 0.668 (0.665;0.672) | 1.038 (1;1.076) | -0.002 (-0.017;0.012) | 0.69 (0.686;0.693) | 1.015 (0.984;1.047) | 0.006 (-0.008;0.02) |
| | female | 0.684 (0.679;0.688) | 1.016 (0.965;1.067) | 0.031 (0.013;0.048) | 0.704 (0.7;0.708) | 0.972 (0.928;1.017) | 0.023 (0.006;0.04) |

| | | | | | | | |
|---|---|---|---|---|---|---|---|
| Finrisk Stroke | male | 0.679 (0.675;0.682) | 1.04 (1.01;1.069) | -0.004 (-0.018;0.011) | 0.701 (0.697;0.704) | 1.016 (0.992;1.04) | 0.005 (-0.009;0.019) |
| | female | 0.689 (0.685;0.694) | 1.007 (0.96;1.054) | 0.032 (0.015;0.05) | 0.71 (0.706;0.714) | 0.969 (0.927;1.011) | 0.025 (0.008;0.041) |
| Framingham 1998 | male | 0.669 (0.665;0.673) | 1.041 (1.007;1.076) | -0.004 (-0.018;0.011) | 0.69 (0.686;0.693) | 1.021 (0.996;1.046) | 0.005 (-0.009;0.02) |
| | female | 0.689 (0.685;0.693) | 1.027 (0.988;1.065) | 0.034 (0.017;0.051) | 0.712 (0.708;0.715) | 0.985 (0.956;1.014) | 0.027 (0.01;0.043) |
| QRISK 2 | male | 0.665 (0.661;0.668) | 1.001 (0.949;1.053) | 0.002 (-0.012;0.016) | 0.681 (0.677;0.684) | 0.969 (0.925;1.013) | 0.011 (-0.003;0.025) |
| | female | 0.687 (0.682;0.691) | 1.006 (0.959;1.052) | 0.031 (0.013;0.049) | 0.707 (0.703;0.711) | 0.966 (0.929;1.002) | 0.023 (0.006;0.04) |
| QRISK 3 | male | 0.657 (0.653;0.661) | 0.99 (0.938;1.043) | 0.002 (-0.013;0.016) | 0.674 (0.67;0.677) | 0.966 (0.921;1.01) | 0.011 (-0.003;0.025) |
| | female | 0.684 (0.68;0.689) | 1.011 (0.961;1.06) | 0.03 (0.012;0.047) | 0.704 (0.7;0.708) | 0.967 (0.928;1.006) | 0.021 (0.004;0.038) |
| RECODE | male | 0.726 (0.722;0.73) | 1.017 (0.985;1.049) | 0 (-0.016;0.016) | 0.728 (0.724;0.732) | 1.007 (0.971;1.043) | 0.01 (-0.006;0.025) |
| | female | 0.73 (0.725;0.735) | 0.994 (0.945;1.043) | 0.034 (0.015;0.054) | 0.735 (0.73;0.739) | 0.982 (0.935;1.03) | 0.025 (0.007;0.043) |
| Reynolds Risk | male | 0.653 (0.65;0.657) | 1.035 (0.988;1.082) | 0 (-0.015;0.014) | 0.673 (0.67;0.677) | 1.019 (0.98;1.058) | 0.008 (-0.006;0.022) |
| | female | 0.66 (0.656;0.665) | 1.039 (0.951;1.127) | 0.03 (0.012;0.048) | 0.679 (0.675;0.684) | 0.987 (0.917;1.057) | 0.022 (0.005;0.04) |
| SCORE CHD | male | 0.654 (0.65;0.657) | 1.043 (0.999;1.086) | 0 (-0.015;0.014) | 0.674 (0.671;0.678) | 1.025 (0.989;1.061) | 0.008 (-0.006;0.022) |
| | female | 0.682 (0.678;0.686) | 1.026 (0.985;1.066) | 0.031 (0.013;0.048) | 0.705 (0.701;0.709) | 0.979 (0.946;1.012) | 0.023 (0.007;0.039) |
| SCORE CVD | male | 0.663 (0.659;0.667) | 1.041 (1.002;1.081) | -0.001 (-0.016;0.013) | 0.684 (0.681;0.688) | 1.023 (0.991;1.055) | 0.008 (-0.006;0.022) |
| | female | 0.688 (0.683;0.692) | 1.02 (0.983;1.057) | 0.032 (0.015;0.049) | 0.711 (0.707;0.715) | 0.978 (0.948;1.008) | 0.024 (0.008;0.04) |
| UKPDS 56 | male | 0.614 (0.609;0.618) | 1.015 (0.915;1.115) | 0 (-0.014;0.014) | 0.63 (0.625;0.634) | 1.017 (0.934;1.1) | 0.008 (-0.005;0.022) |
| | female | 0.642 (0.638;0.647) | 1.042 (0.944;1.14) | 0.028 (0.01;0.046) | 0.66 (0.655;0.664) | 0.988 (0.917;1.059) | 0.019 (0.002;0.037) |
| UKPDS 68 C-HF | male | 0.659 (0.655;0.663) | 1.043 (0.998;1.089) | -0.001 (-0.016;0.014) | 0.683 (0.679;0.687) | 1.024 (0.984;1.064) | 0.008 (-0.007;0.022) |
| | female | 0.674 (0.669;0.678) | 0.996 (0.961;1.031) | 0.033 (0.015;0.05) | 0.701 (0.697;0.705) | 0.965 (0.934;0.996) | 0.026 (0.009;0.042) |
| UKPDS 68 Stroke | male | 0.659 (0.655;0.663) | 1.043 (0.998;1.089) | -0.001 (-0.016;0.014) | 0.683 (0.679;0.687) | 1.024 (0.984;1.064) | 0.008 (-0.007;0.022) |
| | female | 0.674 (0.669;0.678) | 0.996 (0.961;1.031) | 0.033 (0.015;0.05) | 0.701 (0.697;0.705) | 0.965 (0.934;0.996) | 0.026 (0.009;0.042) |
| UKPDS 82 C-HF | male | 0.648 (0.644;0.653) | 1.055 (0.997;1.113) | -0.001 (-0.016;0.014) | 0.672 (0.668;0.677) | 1.022 (0.975;1.07) | 0.008 (-0.007;0.022) |
| | female | 0.656 (0.651;0.662) | 0.961 (0.918;1.004) | 0.029 (0.011;0.047) | 0.683 (0.678;0.687) | 0.938 (0.902;0.974) | 0.021 (0.003;0.038) |
| UKPDS 82 CHD | male | 0.651 (0.647;0.656) | 1.037 (0.977;1.096) | -0.001 (-0.016;0.013) | 0.669 (0.665;0.673) | 1.008 (0.954;1.061) | 0.007 (-0.007;0.022) |
| | female | 0.689 (0.683;0.695) | 0.952 (0.915;0.99) | 0.036 (0.018;0.053) | 0.707 (0.702;0.713) | 0.96 (0.925;0.996) | 0.027 (0.01;0.043) |

**Appendix Table 17 Discrimination (c-statistic) and calibration (calibration-in-the-large (CIL) and calibration slope (CS)) for all included risk scores against CVD and CVD + AF + HF outcomes for the imputed dataset stratified by age subgroups (1st bin from 18 to less than 50 years, 2nd bin from 50 to less than 65 years, 3rd bin from 65 to less than 75 years, and 4th bin from 75).**

| Risk Score Name | Age-group (years) | **CVD** | | | **CVD + AF + HF** | | |
|---|---|---|---|---|---|---|---|
| | | **C-stat** | **CS** | **CIL** | **C-stat** | **CS** | **CIL** |
| ASCVD | [18, 50) | 0.615 (0.605;0.625) | 1.248 (0.83;1.665) | 0.015 (-0.019;0.048) | 0.604 (0.595;0.613) | 1.286 (0.888;1.685) | 0.012 (-0.019;0.043) |
| | [50, 65) | 0.593 (0.588;0.599) | 0.941 (0.844;1.038) | -0.018 (-0.037;0.001) | 0.592 (0.587;0.597) | 0.953 (0.854;1.052) | -0.011 (-0.029;0.007) |
| | [65, 75) | 0.573 (0.567;0.579) | 1.062 (0.929;1.195) | -0.043 (-0.064;-0.023) | 0.573 (0.567;0.579) | 1.165 (0.993;1.338) | -0.043 (-0.062;-0.023) |
| | [75, 105] | 0.529 (0.522;0.536) | 0.63 (0.307;0.952) | 0.076 (0.054;0.098) | 0.546 (0.539;0.552) | 1.234 (0.877;1.591) | 0.057 (0.035;0.079) |
| | [18, 50) | 0.629 (0.619;0.639) | 1.094 (0.898;1.289) | 0.017 (-0.018;0.051) | 0.619 (0.61;0.628) | 1.118 (0.927;1.309) | 0.014 (-0.017;0.046) |
| | [50, 65) | 0.572 (0.566;0.578) | 1.073 (0.884;1.261) | -0.018 (-0.036;0.001) | 0.571 (0.565;0.576) | 1.062 (0.879;1.246) | -0.011 (-0.028;0.007) |
| | [65, 75) | 0.54 (0.533;0.546) | 0.967 (0.718;1.217) | -0.038 (-0.058;-0.018) | 0.538 (0.532;0.545) | 1.009 (0.698;1.319) | -0.038 (-0.057;-0.019) |
| Framingham 1991 fatal CHD | [75, 105] | 0.516 (0.508;0.523) | 1.212 (-0.17;2.593) | 0.076 (0.054;0.098) | 0.521 (0.514;0.528) | 0.922 (0.409;1.435) | 0.057 (0.035;0.079) |
| | [18, 50) | 0.623 (0.613;0.633) | 1.097 (0.951;1.243) | 0.017 (-0.017;0.052) | 0.614 (0.605;0.623) | 1.136 (0.958;1.313) | 0.015 (-0.017;0.046) |
| | [50, 65) | 0.583 (0.577;0.589) | 1.057 (0.881;1.233) | -0.017 (-0.036;0.002) | 0.581 (0.575;0.586) | 1.056 (0.882;1.231) | -0.01 (-0.028;0.008) |
| | [65, 75) | 0.557 (0.551;0.563) | 1.069 (0.895;1.243) | -0.038 (-0.058;-0.018) | 0.553 (0.547;0.559) | 1.081 (0.863;1.299) | -0.038 (-0.057;-0.018) |
| Framingham 1991 CVD | [75, 105] | 0.535 (0.528;0.542) | 1.092 (0.62;1.564) | 0.076 (0.054;0.099) | 0.537 (0.53;0.543) | 0.988 (0.663;1.314) | 0.058 (0.036;0.08) |
| | [18, 50) | 0.628 (0.619;0.638) | 1.023 (0.93;1.116) | 0.02 (-0.014;0.055) | 0.62 (0.611;0.63) | 1.061 (0.943;1.18) | 0.017 (-0.014;0.049) |
| | [50, 65) | 0.576 (0.57;0.582) | 1.132 (0.941;1.324) | -0.019 (-0.038;0) | 0.576 (0.57;0.583) | 1.087 (0.916;1.257) | -0.012 (-0.029;0.006) |
| | [65, 75) | 0.543 (0.537;0.55) | 1.125 (0.849;1.4) | -0.039 (-0.059;-0.018) | 0.545 (0.539;0.551) | 1.069 (0.805;1.334) | -0.039 (-0.058;-0.019) |
| Framingham 1991 Stroke | [75, 105] | 0.526 (0.519;0.533) | 1.117 (0.603;1.632) | 0.076 (0.054;0.098) | 0.534 (0.527;0.541) | 1.132 (0.697;1.566) | 0.057 (0.035;0.079) |

| | | | | | | | |
|---|---|---|---|---|---|---|---|
| | [18, 50) | 0.63 (0.621;0.639) | 1.138 (1.047;1.229) | 0.011 (-0.023;0.044) | 0.62 (0.612;0.628) | 1.158 (1.067;1.249) | 0.009 (-0.022;0.04) |
| | [50, 65) | 0.585 (0.58;0.59) | 1.011 (0.939;1.083) | -0.021 (-0.039;-0.002) | 0.589 (0.584;0.594) | 1.064 (0.997;1.131) | -0.014 (-0.031;0.004) |
| | [65, 75) | 0.567 (0.561;0.574) | 1.007 (0.893;1.121) | -0.035 (-0.055;-0.015) | 0.572 (0.566;0.578) | 1.083 (0.972;1.194) | -0.035 (-0.054;-0.015) |
| CHS Advanced | [75, 105] | 0.533 (0.526;0.539) | 0.766 (0.538;0.994) | 0.077 (0.054;0.099) | 0.551 (0.544;0.557) | 1.16 (0.936;1.384) | 0.058 (0.036;0.08) |
| | [18, 50) | 0.629 (0.62;0.638) | 1.139 (1.045;1.233) | 0.011 (-0.023;0.044) | 0.619 (0.61;0.627) | 1.156 (1.063;1.25) | 0.009 (-0.022;0.04) |
| | [50, 65) | 0.587 (0.582;0.592) | 0.997 (0.922;1.072) | -0.02 (-0.039;-0.001) | 0.59 (0.585;0.595) | 1.051 (0.981;1.12) | -0.013 (-0.031;0.005) |
| | [65, 75) | 0.569 (0.562;0.575) | 1.008 (0.884;1.131) | -0.034 (-0.054;-0.014) | 0.571 (0.565;0.577) | 1.072 (0.951;1.193) | -0.034 (-0.053;-0.014) |
| CHS Basic | [75, 105] | 0.536 (0.53;0.543) | 0.799 (0.584;1.014) | 0.077 (0.055;0.099) | 0.552 (0.546;0.559) | 1.144 (0.913;1.374) | 0.058 (0.036;0.08) |
| | [18, 50) | 0.62 (0.61;0.631) | 1.235 (1.018;1.451) | 0.008 (-0.025;0.042) | 0.61 (0.601;0.62) | 1.198 (1.003;1.394) | 0.007 (-0.024;0.039) |
| | [50, 65) | 0.542 (0.535;0.548) | 0.919 (0.525;1.314) | -0.021 (-0.04;-0.003) | 0.542 (0.535;0.549) | 0.974 (0.555;1.394) | -0.014 (-0.032;0.004) |
| | [65, 75) | 0.509 (0.491;0.526) | -0.452 (-10.744;9.839) | -0.038 (-0.058;-0.018) | 0.512 (0.506;0.518) | 2.436 (-2.858;7.731) | -0.038 (-0.057;-0.019) |
| DARTS | [75, 105] | 0.502 (0.489;0.515) | 0.477 (-3.223;4.177) | 0.077 (0.054;0.099) | 0.502 (0.488;0.515) | -0.487 (-5.277;4.304) | 0.058 (0.036;0.079) |
| | [18, 50) | 0.632 (0.623;0.642) | 1.13 (0.964;1.296) | 0.018 (-0.016;0.052) | 0.622 (0.613;0.63) | 1.192 (1.024;1.36) | 0.015 (-0.016;0.046) |
| | [50, 65) | 0.566 (0.56;0.571) | 0.959 (0.806;1.111) | -0.02 (-0.038;-0.001) | 0.567 (0.562;0.573) | 0.947 (0.805;1.089) | -0.012 (-0.03;0.006) |
| | [65, 75) | 0.534 (0.527;0.541) | 0.972 (0.685;1.259) | -0.041 (-0.061;-0.021) | 0.535 (0.529;0.542) | 1.163 (0.8;1.526) | -0.04 (-0.059;-0.021) |
| Finrisk CHD | [75, 105] | 0.509 (0.501;0.516) | 0.807 (-0.498;2.112) | 0.076 (0.054;0.098) | 0.521 (0.514;0.528) | 0.923 (0.379;1.467) | 0.057 (0.035;0.078) |
| | [18, 50) | 0.63 (0.621;0.64) | 1.052 (0.91;1.194) | 0.021 (-0.013;0.054) | 0.622 (0.613;0.631) | 1.103 (0.955;1.25) | 0.017 (-0.014;0.048) |
| | [50, 65) | 0.581 (0.575;0.586) | 0.989 (0.869;1.11) | -0.02 (-0.038;-0.001) | 0.583 (0.578;0.588) | 0.987 (0.873;1.101) | -0.012 (-0.03;0.005) |
| | [65, 75) | 0.565 (0.559;0.572) | 1.075 (0.925;1.225) | -0.042 (-0.063;-0.022) | 0.563 (0.557;0.569) | 1.194 (1.007;1.381) | -0.042 (-0.061;-0.022) |
| Finrisk CVD | [75, 105] | 0.54 (0.533;0.547) | 0.869 (0.649;1.089) | 0.076 (0.054;0.098) | 0.549 (0.542;0.556) | 0.938 (0.726;1.15) | 0.057 (0.035;0.079) |
| | [18, 50) | 0.593 (0.584;0.602) | 0.88 (0.771;0.99) | 0.014 (-0.019;0.048) | 0.59 (0.581;0.599) | 0.942 (0.812;1.072) | 0.012 (-0.019;0.043) |
| | [50, 65) | 0.591 (0.585;0.596) | 1.05 (0.961;1.139) | -0.021 (-0.039;-0.002) | 0.594 (0.589;0.599) | 1.051 (0.965;1.138) | -0.013 (-0.031;0.004) |
| Finrisk Stroke | [65, 75) | 0.585 (0.579;0.591) | 1.14 (1.039;1.241) | -0.042 (-0.062;-0.022) | 0.581 (0.575;0.586) | 1.227 (1.11;1.343) | -0.041 (-0.061;-0.022) |

| | | | | | | | |
|---|---|---|---|---|---|---|---|
| | [75, 105] | 0.557 (0.55;0.563) | 0.903 (0.767;1.039) | 0.077 (0.055;0.099) | 0.559 (0.552;0.566) | 0.952 (0.809;1.094) | 0.058 (0.036;0.08) |
| | [18, 50) | 0.631 (0.621;0.64) | 1.073 (0.947;1.198) | 0.022 (-0.012;0.056) | 0.624 (0.615;0.634) | 1.106 (0.963;1.249) | 0.02 (-0.012;0.051) |
| | [50, 65) | 0.568 (0.561;0.574) | 1.161 (0.92;1.402) | -0.021 (-0.04;-0.003) | 0.572 (0.566;0.578) | 1.127 (0.926;1.328) | -0.014 (-0.032;0.003) |
| | [65, 75) | 0.539 (0.532;0.546) | 1.219 (0.913;1.525) | -0.04 (-0.059;-0.02) | 0.545 (0.538;0.552) | 1.311 (0.918;1.705) | -0.04 (-0.059;-0.021) |
| Framingham 1998 | [75, 105] | 0.516 (0.509;0.522) | 0.924 (0.16;1.687) | 0.076 (0.054;0.098) | 0.533 (0.526;0.54) | 1.181 (0.742;1.62) | 0.057 (0.035;0.079) |
| | [18, 50) | 0.658 (0.648;0.668) | 1.053 (0.906;1.199) | 0.022 (-0.012;0.056) | 0.646 (0.638;0.655) | 1.049 (0.916;1.183) | 0.02 (-0.012;0.051) |
| | [50, 65) | 0.605 (0.6;0.61) | 0.984 (0.863;1.104) | -0.02 (-0.039;-0.002) | 0.603 (0.598;0.608) | 0.985 (0.863;1.106) | -0.013 (-0.031;0.005) |
| | [65, 75) | 0.562 (0.556;0.569) | 1.094 (0.844;1.344) | -0.04 (-0.06;-0.019) | 0.562 (0.556;0.568) | 1.211 (0.889;1.533) | -0.039 (-0.059;-0.02) |
| QRISK 2 | [75, 105] | 0.514 (0.507;0.521) | 0.402 (0.077;0.728) | 0.075 (0.053;0.098) | 0.527 (0.52;0.534) | 0.639 (0.416;0.863) | 0.056 (0.034;0.078) |
| | [18, 50) | 0.654 (0.644;0.663) | 1.051 (0.901;1.201) | 0.024 (-0.01;0.058) | 0.643 (0.634;0.652) | 1.04 (0.907;1.174) | 0.022 (-0.01;0.054) |
| | [50, 65) | 0.594 (0.589;0.6) | 1.02 (0.881;1.16) | -0.02 (-0.039;-0.001) | 0.594 (0.589;0.599) | 0.989 (0.857;1.12) | -0.013 (-0.031;0.005) |
| QRISK 3 | [65, 75) | 0.553 (0.546;0.559) | 1.069 (0.791;1.347) | -0.04 (-0.06;-0.019) | 0.554 (0.548;0.56) | 1.132 (0.817;1.448) | -0.04 (-0.059;-0.02) |
| | [75, 105] | 0.512 (0.505;0.519) | 0.506 (-0.005;1.018) | 0.076 (0.054;0.098) | 0.524 (0.517;0.531) | 0.815 (0.441;1.189) | 0.056 (0.035;0.078) |
| | [18, 50) | 0.654 (0.644;0.663) | 1.105 (0.955;1.254) | 0.006 (-0.029;0.041) | 0.638 (0.628;0.647) | 1.099 (0.964;1.235) | 0.005 (-0.027;0.037) |
| | [50, 65) | 0.674 (0.667;0.681) | 0.992 (0.908;1.076) | -0.018 (-0.038;0.003) | 0.659 (0.652;0.665) | 1.019 (0.932;1.107) | -0.01 (-0.029;0.009) |
| | [65, 75) | 0.682 (0.676;0.688) | 1.003 (0.947;1.058) | -0.042 (-0.067;-0.017) | 0.66 (0.655;0.666) | 1.008 (0.95;1.067) | -0.041 (-0.064;-0.018) |
| RECODE | [75, 105] | 0.652 (0.645;0.659) | 0.888 (0.776;0.999) | 0.079 (0.055;0.103) | 0.64 (0.633;0.647) | 0.946 (0.821;1.072) | 0.058 (0.035;0.082) |
| | [18, 50) | 0.632 (0.623;0.642) | 1.039 (0.899;1.18) | 0.018 (-0.016;0.052) | 0.623 (0.614;0.632) | 1.094 (0.974;1.214) | 0.015 (-0.017;0.046) |
| | [50, 65) | 0.57 (0.564;0.576) | 1.027 (0.873;1.18) | -0.02 (-0.038;-0.001) | 0.571 (0.566;0.577) | 0.994 (0.844;1.143) | -0.012 (-0.03;0.006) |
| | [65, 75) | 0.544 (0.538;0.551) | 0.951 (0.736;1.166) | -0.041 (-0.061;-0.021) | 0.543 (0.537;0.549) | 1.068 (0.75;1.386) | -0.041 (-0.06;-0.021) |
| Reynolds Risk | [75, 105] | 0.517 (0.51;0.524) | 0.902 (0.232;1.572) | 0.076 (0.054;0.098) | 0.527 (0.52;0.534) | 0.981 (0.368;1.593) | 0.057 (0.035;0.079) |
| | [18, 50) | 0.645 (0.636;0.654) | 1.236 (1.121;1.351) | 0.015 (-0.019;0.048) | 0.632 (0.623;0.64) | 1.253 (1.137;1.369) | 0.012 (-0.019;0.043) |
| SCORE CHD | [50, 65) | 0.584 (0.578;0.589) | 0.946 (0.841;1.05) | -0.019 (-0.038;0) | 0.583 (0.578;0.588) | 0.939 (0.84;1.039) | -0.012 (-0.029;0.006) |

| | | | | | | | |
|---|---|---|---|---|---|---|---|
| | [65, 75) | 0.547 (0.541;0.553) | 1.045 (0.826;1.265) | -0.04 (-0.06;-0.02) | 0.547 (0.541;0.553) | 1.188 (0.874;1.502) | -0.04 (-0.059;-0.02) |
| | [75, 105] | 0.512 (0.506;0.519) | 0.933 (0.178;1.689) | 0.075 (0.053;0.097) | 0.525 (0.518;0.531) | 0.973 (0.569;1.377) | 0.056 (0.034;0.078) |
| | [18, 50) | 0.649 (0.64;0.658) | 1.205 (1.101;1.309) | 0.016 (-0.018;0.05) | 0.636 (0.628;0.645) | 1.221 (1.113;1.328) | 0.013 (-0.018;0.045) |
| | [50, 65) | 0.587 (0.582;0.593) | 0.985 (0.88;1.09) | -0.019 (-0.038;-0.001) | 0.589 (0.583;0.594) | 0.971 (0.873;1.07) | -0.012 (-0.03;0.005) |
| | [65, 75) | 0.549 (0.542;0.555) | 1.024 (0.822;1.226) | -0.042 (-0.062;-0.021) | 0.551 (0.545;0.557) | 1.158 (0.893;1.423) | -0.041 (-0.06;-0.022) |
| SCORE CVD | [75, 105] | 0.515 (0.509;0.522) | 0.828 (0.373;1.283) | 0.075 (0.053;0.097) | 0.53 (0.523;0.536) | 1.012 (0.681;1.343) | 0.056 (0.033;0.078) |
| | [18, 50) | 0.602 (0.592;0.612) | 1.191 (0.911;1.471) | 0.009 (-0.024;0.043) | 0.591 (0.582;0.601) | 1.214 (0.982;1.445) | 0.007 (-0.024;0.039) |
| | [50, 65) | 0.558 (0.551;0.564) | 0.96 (0.768;1.151) | -0.019 (-0.038;0) | 0.556 (0.55;0.563) | 0.97 (0.782;1.158) | -0.012 (-0.03;0.006) |
| | [65, 75) | 0.542 (0.535;0.549) | 1.046 (0.782;1.309) | -0.04 (-0.06;-0.02) | 0.538 (0.531;0.544) | 1.173 (0.818;1.529) | -0.039 (-0.058;-0.02) |
| UKPDS 56 | [75, 105] | 0.522 (0.514;0.529) | 0.871 (0.367;1.375) | 0.076 (0.054;0.099) | 0.527 (0.52;0.534) | 0.84 (0.503;1.178) | 0.057 (0.035;0.079) |
| | [18, 50) | 0.58 (0.57;0.59) | 0.92 (0.744;1.095) | 0.01 (-0.024;0.044) | 0.583 (0.573;0.592) | 0.88 (0.732;1.028) | 0.01 (-0.022;0.042) |
| | [50, 65) | 0.531 (0.525;0.537) | 2.013 (0.394;3.632) | -0.021 (-0.039;-0.003) | 0.544 (0.539;0.55) | 1.275 (0.75;1.8) | -0.014 (-0.031;0.004) |
| | [65, 75) | 0.514 (0.506;0.521) | 0.628 (0.006;1.249) | -0.039 (-0.059;-0.019) | 0.527 (0.52;0.535) | 0.918 (0.212;1.624) | -0.04 (-0.06;-0.021) |
| UKPDS 68 C-HF | [75, 105] | 0.507 (0.498;0.515) | 0.609 (-0.763;1.98) | 0.076 (0.054;0.098) | 0.528 (0.52;0.537) | 1.379 (0.117;2.641) | 0.057 (0.035;0.079) |
| | [18, 50) | 0.58 (0.57;0.59) | 0.92 (0.744;1.095) | 0.01 (-0.024;0.044) | 0.583 (0.573;0.592) | 0.88 (0.732;1.028) | 0.01 (-0.022;0.042) |
| | [50, 65) | 0.531 (0.525;0.537) | 2.013 (0.394;3.632) | -0.021 (-0.039;-0.003) | 0.544 (0.539;0.55) | 1.275 (0.75;1.8) | -0.014 (-0.031;0.004) |
| | [65, 75) | 0.514 (0.506;0.521) | 0.628 (0.006;1.249) | -0.039 (-0.059;-0.019) | 0.527 (0.52;0.535) | 0.918 (0.212;1.624) | -0.04 (-0.06;-0.021) |
| UKPDS 68 Stroke | [75, 105] | 0.507 (0.498;0.515) | 0.609 (-0.763;1.98) | 0.076 (0.054;0.098) | 0.528 (0.52;0.537) | 1.379 (0.117;2.641) | 0.057 (0.035;0.079) |
| | [18, 50) | 0.58 (0.57;0.591) | 1.052 (0.761;1.344) | 0.01 (-0.024;0.044) | 0.582 (0.573;0.592) | 0.959 (0.758;1.16) | 0.011 (-0.021;0.043) |
| | [50, 65) | 0.528 (0.52;0.536) | 1.286 (0.867;1.706) | -0.021 (-0.04;-0.003) | 0.541 (0.533;0.548) | 1.082 (0.826;1.338) | -0.014 (-0.032;0.004) |
| | [65, 75) | 0.507 (0.498;0.515) | 0.474 (-0.085;1.034) | -0.038 (-0.058;-0.018) | 0.522 (0.514;0.53) | 1.137 (0.365;1.908) | -0.038 (-0.057;-0.019) |
| UKPDS 82 C-HF | [75, 105] | 0.497 (0.489;0.505) | -0.429 (-1.748;0.891) | 0.076 (0.054;0.098) | 0.519 (0.51;0.527) | 0.874 (0.32;1.429) | 0.057 (0.035;0.079) |
| UKPDS 82 CHD | [18, 50) | 0.557 (0.546;0.567) | 0.822 (0.642;1.003) | 0.006 (-0.027;0.04) | 0.555 (0.545;0.565) | 0.745 (0.603;0.887) | 0.006 (-0.026;0.037) |

| | | | | | | | |
|---|---|---|---|---|---|---|---|
| | [50, 65) | 0.519 (0.51;0.527) | 1.925 (0.693;3.156) | -0.022 (-0.04;-0.003) | 0.524 (0.515;0.533) | 1.362 (0.908;1.816) | -0.015 (-0.032;0.003) |
| | [65, 75) | 0.513 (0.504;0.521) | 0.713 (0.459;0.966) | -0.038 (-0.058;-0.017) | 0.52 (0.512;0.528) | 0.951 (0.709;1.193) | -0.038 (-0.057;-0.018) |
| | [75, 105] | 0.515 (0.508;0.523) | 0.779 (0.49;1.069) | 0.074 (0.052;0.096) | 0.531 (0.524;0.538) | 1.297 (0.942;1.652) | 0.055 (0.033;0.077) |

**Appendix Table 18 Discrimination (c-statistic) and calibration (calibration-in-the-large (CIL) and calibration slope (CS)) for all included risk scores against CVD and CVD + AF + HF outcomes for the imputed dataset stratified by statins usage at the baseline (statin naïve vs statin users).**

| Risk Score Name | Statins usage at the baseline (statin naïve vs statin users) | CVD | | | CVD + AF + HF | | |
|---|---|---|---|---|---|---|---|
| | | C-stat | CS | CIL | C-stat | CS | CIL |
| ASCVD | statin naïve | 0.692 (0.688;0.695) | 1.022 (0.989;1.055) | 0.026 (0.012;0.041) | 0.713 (0.71;0.716) | 1.011 (0.983;1.038) | 0.025 (0.011;0.039) |
| | statin users | 0.623 (0.619;0.628) | 1.002 (0.949;1.054) | -0.024 (-0.042;-0.007) | 0.643 (0.639;0.648) | 0.955 (0.915;0.996) | -0.025 (-0.042;-0.008) |
| Framingham 1991 fatal CHD | statin naïve | 0.659 (0.655;0.663) | 1.107 (1.054;1.16) | 0.028 (0.014;0.043) | 0.672 (0.668;0.675) | 1.06 (1.024;1.095) | 0.026 (0.013;0.04) |
| | statin users | 0.589 (0.584;0.594) | 1.02 (0.923;1.118) | -0.02 (-0.038;-0.003) | 0.598 (0.593;0.603) | 1.006 (0.92;1.093) | -0.021 (-0.038;-0.004) |
| Framingham 1991 CVD | statin naïve | 0.644 (0.64;0.649) | 1.111 (1.056;1.167) | 0.028 (0.013;0.042) | 0.652 (0.647;0.656) | 1.061 (1.023;1.099) | 0.025 (0.012;0.039) |
| | statin users | 0.593 (0.588;0.598) | 1.039 (0.954;1.123) | -0.021 (-0.038;-0.003) | 0.597 (0.592;0.602) | 1.043 (0.967;1.119) | -0.021 (-0.038;-0.004) |
| Framingham 1991 Stroke | statin naïve | 0.668 (0.663;0.672) | 1.086 (1.039;1.132) | 0.03 (0.016;0.045) | 0.684 (0.68;0.688) | 1.05 (1.018;1.083) | 0.029 (0.015;0.043) |
| | statin users | 0.602 (0.597;0.607) | 1.027 (0.964;1.09) | -0.021 (-0.039;-0.004) | 0.616 (0.612;0.621) | 1.04 (0.989;1.09) | -0.022 (-0.039;-0.005) |
| CHS Advanced | statin naïve | 0.693 (0.69;0.696) | 1.031 (1.007;1.055) | 0.025 (0.01;0.039) | 0.72 (0.716;0.723) | 1.02 (1;1.041) | 0.022 (0.009;0.036) |
| | statin users | 0.626 (0.622;0.631) | 0.988 (0.949;1.028) | -0.021 (-0.038;-0.003) | 0.649 (0.645;0.654) | 0.95 (0.918;0.982) | -0.021 (-0.038;-0.004) |
| CHS Basic | statin naïve | 0.693 (0.689;0.696) | 1.033 (1.007;1.058) | 0.024 (0.01;0.038) | 0.719 (0.716;0.722) | 1.021 (0.999;1.043) | 0.021 (0.008;0.035) |
| | statin users | 0.628 (0.624;0.633) | 0.994 (0.954;1.033) | -0.021 (-0.038;-0.003) | 0.651 (0.646;0.655) | 0.955 (0.923;0.987) | -0.021 (-0.038;-0.003) |
| DARTS | statin naïve | 0.671 (0.667;0.675) | 1.063 (1.017;1.109) | 0.026 (0.011;0.04) | 0.689 (0.686;0.693) | 1.048 (1.008;1.087) | 0.024 (0.009;0.038) |
| | statin users | 0.579 (0.573;0.584) | 0.953 (0.862;1.045) | -0.026 (-0.044;-0.009) | 0.594 (0.589;0.599) | 0.896 (0.83;0.962) | -0.028 (-0.045;-0.011) |
| Finrisk CHD | statin naïve | 0.688 (0.684;0.691) | 1.062 (1.028;1.095) | 0.027 (0.012;0.042) | 0.707 (0.704;0.71) | 1.04 (1.012;1.069) | 0.026 (0.012;0.039) |
| | statin users | 0.599 (0.594;0.604) | 1.006 (0.943;1.07) | -0.024 (-0.041;-0.006) | 0.617 (0.612;0.622) | 0.961 (0.91;1.013) | -0.024 (-0.042;-0.007) |
| Finrisk CVD | statin naïve | 0.696 (0.692;0.7) | 1.045 (1.017;1.074) | 0.028 (0.013;0.042) | 0.716 (0.713;0.72) | 1.024 (1.001;1.048) | 0.026 (0.012;0.04) |
| | statin users | 0.62 (0.616;0.625) | 1.001 (0.954;1.048) | -0.02 (-0.038;-0.003) | 0.639 (0.634;0.643) | 0.956 (0.916;0.996) | -0.02 (-0.038;-0.003) |
| Finrisk Stroke | statin naïve | 0.696 (0.692;0.7) | 1.032 (1.006;1.059) | 0.028 (0.013;0.042) | 0.717 (0.714;0.721) | 1.012 (0.99;1.034) | 0.026 (0.012;0.04) |
| | statin users | 0.632 (0.627;0.636) | 1.001 (0.96;1.041) | -0.016 (-0.034;0.001) | 0.649 (0.645;0.654) | 0.954 (0.919;0.988) | -0.016 (-0.033;0.002) |
| Framingham 1998 | statin naïve | 0.691 (0.687;0.694) | 1.039 (1.007;1.071) | 0.029 (0.015;0.044) | 0.715 (0.712;0.718) | 1.022 (0.999;1.045) | 0.028 (0.014;0.042) |

| | | | | | | | |
|---|---|---|---|---|---|---|---|
| | statin users | 0.609 (0.605;0.614) | 1 (0.952;1.048) | -0.022 (-0.04;-0.005) | 0.632 (0.627;0.636) | 0.994 (0.954;1.033) | -0.023 (-0.04;-0.006) |
| QRISK 2 | statin naïve | 0.687 (0.683;0.69) | 1.041 (0.995;1.087) | 0.027 (0.012;0.041) | 0.707 (0.704;0.711) | 1.022 (0.989;1.056) | 0.025 (0.011;0.039) |
| | statin users | 0.62 (0.615;0.625) | 0.98 (0.927;1.033) | -0.023 (-0.04;-0.005) | 0.636 (0.632;0.641) | 0.963 (0.913;1.013) | -0.023 (-0.04;-0.005) |
| QRISK 3 | statin naïve | 0.685 (0.681;0.689) | 1.04 (0.997;1.083) | 0.027 (0.012;0.042) | 0.705 (0.702;0.708) | 1.017 (0.985;1.049) | 0.025 (0.011;0.04) |
| | statin users | 0.613 (0.608;0.618) | 0.993 (0.935;1.05) | -0.022 (-0.039;-0.004) | 0.63 (0.625;0.634) | 0.957 (0.902;1.012) | -0.022 (-0.039;-0.004) |
| RECODE | statin naïve | 0.709 (0.704;0.713) | 1.077 (1.033;1.121) | 0.026 (0.011;0.041) | 0.717 (0.713;0.721) | 1.066 (1.031;1.101) | 0.023 (0.009;0.037) |
| | statin users | 0.743 (0.738;0.747) | 0.992 (0.955;1.029) | -0.011 (-0.031;0.009) | 0.74 (0.735;0.744) | 0.951 (0.917;0.985) | -0.012 (-0.031;0.008) |
| Reynolds Risk | statin naïve | 0.685 (0.681;0.688) | 1.057 (1.019;1.094) | 0.028 (0.013;0.043) | 0.702 (0.699;0.705) | 1.034 (1.004;1.065) | 0.026 (0.012;0.04) |
| | statin users | 0.602 (0.597;0.607) | 1.01 (0.944;1.077) | -0.022 (-0.04;-0.005) | 0.617 (0.612;0.622) | 0.966 (0.914;1.019) | -0.023 (-0.04;-0.005) |
| SCORE CHD | statin naïve | 0.694 (0.691;0.698) | 1.066 (1.035;1.097) | 0.027 (0.012;0.041) | 0.714 (0.71;0.717) | 1.043 (1.019;1.066) | 0.025 (0.011;0.038) |
| | statin users | 0.607 (0.603;0.612) | 1.028 (0.977;1.08) | -0.024 (-0.041;-0.006) | 0.625 (0.62;0.63) | 0.967 (0.927;1.008) | -0.024 (-0.041;-0.007) |
| SCORE CVD | statin naïve | 0.698 (0.694;0.701) | 1.056 (1.029;1.083) | 0.028 (0.013;0.042) | 0.719 (0.716;0.722) | 1.036 (1.014;1.057) | 0.026 (0.013;0.04) |
| | statin users | 0.614 (0.609;0.619) | 1.015 (0.968;1.062) | -0.023 (-0.041;-0.006) | 0.634 (0.63;0.639) | 0.963 (0.926;1.001) | -0.024 (-0.041;-0.006) |
| UKPDS 56 | statin naïve | 0.659 (0.655;0.664) | 1.079 (1.025;1.134) | 0.024 (0.01;0.039) | 0.67 (0.666;0.673) | 1.058 (1.01;1.106) | 0.022 (0.008;0.035) |
| | statin users | 0.584 (0.578;0.59) | 1.002 (0.913;1.091) | -0.023 (-0.04;-0.006) | 0.591 (0.586;0.597) | 0.929 (0.849;1.009) | -0.023 (-0.04;-0.006) |
| UKPDS 68 C-HF | statin naïve | 0.671 (0.668;0.675) | 1.04 (0.998;1.082) | 0.026 (0.012;0.041) | 0.7 (0.697;0.704) | 1.031 (0.997;1.064) | 0.025 (0.011;0.039) |
| | statin users | 0.598 (0.593;0.603) | 0.933 (0.867;0.998) | -0.021 (-0.039;-0.003) | 0.625 (0.62;0.629) | 0.923 (0.879;0.968) | -0.021 (-0.038;-0.003) |
| UKPDS 68 Stroke | statin naïve | 0.671 (0.668;0.675) | 1.04 (0.998;1.082) | 0.026 (0.012;0.041) | 0.7 (0.697;0.704) | 1.031 (0.997;1.064) | 0.025 (0.011;0.039) |
| | statin users | 0.598 (0.593;0.603) | 0.933 (0.867;0.998) | -0.021 (-0.039;-0.003) | 0.625 (0.62;0.629) | 0.923 (0.879;0.968) | -0.021 (-0.038;-0.003) |
| UKPDS 82 C-HF | statin naïve | 0.658 (0.653;0.663) | 1.021 (0.981;1.061) | 0.022 (0.007;0.037) | 0.687 (0.683;0.691) | 1.013 (0.977;1.048) | 0.019 (0.005;0.034) |
| | statin users | 0.588 (0.583;0.593) | 0.89 (0.803;0.977) | -0.021 (-0.038;-0.003) | 0.614 (0.609;0.619) | 0.911 (0.847;0.976) | -0.021 (-0.038;-0.003) |
| UKPDS 82 CHD | statin naïve | 0.648 (0.643;0.653) | 1.032 (0.978;1.086) | 0.026 (0.012;0.04) | 0.671 (0.666;0.675) | 1.036 (0.993;1.079) | 0.024 (0.01;0.037) |
| | statin users | 0.586 (0.58;0.591) | 1.011 (0.93;1.091) | -0.024 (-0.042;-0.007) | 0.605 (0.6;0.61) | 1.024 (0.955;1.094) | -0.026 (-0.043;-0.008) |

**Appendix Table 19 Discrimination (c-statistic) and calibration (calibration-in-the-large (CIL) and calibration slope (CS)) for all included risk scores against CVD and CVD + AF + HF outcomes for the imputed dataset stratified by CVD history at the baseline (absent vs present).**

| Risk Score Name | CVD history at the baseline (absent vs present) | CVD | | | CVD + AF + HF | | |
|---|---|---|---|---|---|---|---|
| | | C-stat | CS | CIL | C-stat | CS | CIL |
| ASCVD | absent | 0.667 (0.664;0.671) | 0.943 (0.91;0.977) | -0.012 (-0.026;0.002) | 0.689 (0.686;0.693) | 0.95 (0.922;0.979) | -0.006 (-0.019;0.007) |
| | present | 0.52 (0.513;0.528) | 0.277 (0.077;0.477) | 0.01 (-0.014;0.035) | 0.552 (0.544;0.56) | 0.626 (0.51;0.743) | 0.047 (0.021;0.073) |
| Framingham 1991 fatal CHD | absent | 0.643 (0.639;0.647) | 0.946 (0.903;0.989) | -0.009 (-0.023;0.004) | 0.654 (0.65;0.657) | 0.949 (0.914;0.984) | -0.002 (-0.015;0.01) |
| | present | 0.53 (0.522;0.538) | 0.814 (0.534;1.095) | 0.013 (-0.011;0.038) | 0.544 (0.536;0.553) | 0.89 (0.641;1.138) | 0.052 (0.027;0.078) |
| Framingham 1991 CVD | absent | 0.63 (0.626;0.634) | 0.959 (0.918;1) | -0.009 (-0.023;0.004) | 0.636 (0.632;0.639) | 0.958 (0.922;0.995) | -0.002 (-0.014;0.01) |
| | present | 0.538 (0.53;0.546) | 0.769 (0.565;0.974) | 0.015 (-0.01;0.039) | 0.545 (0.537;0.554) | 0.835 (0.615;1.055) | 0.054 (0.028;0.079) |
| Framingham 1991 Stroke | absent | 0.647 (0.643;0.651) | 0.974 (0.929;1.019) | -0.005 (-0.019;0.008) | 0.664 (0.66;0.667) | 0.983 (0.942;1.024) | 0.002 (-0.011;0.014) |
| | present | 0.526 (0.519;0.534) | 0.63 (0.422;0.838) | 0.014 (-0.011;0.038) | 0.545 (0.537;0.553) | 0.828 (0.605;1.052) | 0.053 (0.028;0.079) |
| CHS Advanced | absent | 0.66 (0.656;0.663) | 0.936 (0.912;0.959) | -0.008 (-0.021;0.006) | 0.689 (0.686;0.692) | 0.944 (0.925;0.962) | 0 (-0.012;0.012) |
| | present | 0.513 (0.506;0.52) | 0.364 (0.136;0.592) | 0.011 (-0.013;0.035) | 0.547 (0.54;0.554) | 0.752 (0.627;0.876) | 0.048 (0.023;0.074) |
| CHS Basic | absent | 0.659 (0.656;0.663) | 0.936 (0.912;0.96) | -0.008 (-0.021;0.006) | 0.688 (0.685;0.692) | 0.944 (0.925;0.963) | 0 (-0.013;0.012) |
| | present | 0.515 (0.507;0.522) | 0.406 (0.199;0.612) | 0.011 (-0.013;0.035) | 0.548 (0.54;0.555) | 0.748 (0.626;0.871) | 0.048 (0.022;0.074) |
| DARTS | absent | 0.651 (0.647;0.655) | 0.926 (0.873;0.98) | -0.01 (-0.024;0.004) | 0.669 (0.665;0.672) | 0.932 (0.889;0.975) | -0.002 (-0.016;0.011) |
| | present | 0.516 (0.507;0.524) | 0.39 (0.095;0.685) | 0.013 (-0.011;0.038) | 0.542 (0.533;0.55) | 0.723 (0.47;0.976) | 0.053 (0.027;0.078) |
| Finrisk CHD | absent | 0.665 (0.661;0.668) | 0.955 (0.926;0.984) | -0.01 (-0.024;0.004) | 0.684 (0.681;0.688) | 0.953 (0.927;0.978) | -0.003 (-0.016;0.01) |
| | present | 0.52 (0.513;0.528) | 0.514 (0.296;0.732) | 0.011 (-0.013;0.036) | 0.548 (0.54;0.556) | 0.788 (0.617;0.959) | 0.05 (0.024;0.075) |
| Finrisk CVD | absent | 0.668 (0.665;0.672) | 0.949 (0.923;0.975) | -0.007 (-0.021;0.006) | 0.69 (0.687;0.694) | 0.946 (0.924;0.968) | 0 (-0.013;0.013) |
| | present | 0.527 (0.52;0.535) | 0.65 (0.468;0.831) | 0.011 (-0.014;0.035) | 0.554 (0.547;0.562) | 0.843 (0.7;0.986) | 0.048 (0.023;0.074) |
| Finrisk Stroke | absent | 0.664 (0.661;0.668) | 0.94 (0.915;0.965) | -0.005 (-0.018;0.009) | 0.688 (0.685;0.691) | 0.939 (0.918;0.96) | 0.003 (-0.009;0.016) |
| | present | 0.533 (0.525;0.54) | 0.761 (0.591;0.931) | 0.011 (-0.014;0.035) | 0.556 (0.549;0.564) | 0.898 (0.759;1.037) | 0.049 (0.023;0.074) |

| | | | | | | | |
|---|---|---|---|---|---|---|---|
| Framingham 1998 | absent | 0.663 (0.66;0.667) | 0.943 (0.915;0.971) | -0.009 (-0.023;0.004) | 0.689 (0.686;0.692) | 0.954 (0.93;0.977) | -0.002 (-0.015;0.01) |
| | present | 0.514 (0.507;0.521) | 0.483 (0.211;0.755) | 0.013 (-0.012;0.037) | 0.547 (0.54;0.555) | 0.862 (0.711;1.013) | 0.051 (0.026;0.077) |
| QRISK 2 | absent | 0.663 (0.659;0.666) | 0.932 (0.892;0.973) | -0.014 (-0.028;0) | 0.683 (0.68;0.686) | 0.935 (0.909;0.961) | -0.007 (-0.02;0.006) |
| | present | 0.522 (0.514;0.529) | 0.459 (0.275;0.642) | 0.01 (-0.015;0.034) | 0.549 (0.542;0.557) | 0.671 (0.519;0.822) | 0.047 (0.021;0.072) |
| QRISK 3 | absent | 0.663 (0.66;0.667) | 0.936 (0.891;0.982) | -0.014 (-0.028;0) | 0.683 (0.68;0.686) | 0.935 (0.907;0.962) | -0.007 (-0.02;0.006) |
| | present | 0.522 (0.515;0.53) | 0.521 (0.314;0.728) | 0.009 (-0.016;0.033) | 0.55 (0.542;0.558) | 0.723 (0.552;0.894) | 0.045 (0.019;0.071) |
| RECODE | absent | 0.64 (0.636;0.645) | 0.98 (0.929;1.031) | -0.007 (-0.021;0.008) | 0.657 (0.653;0.661) | 0.963 (0.926;1) | 0.001 (-0.013;0.014) |
| | present | 0.534 (0.527;0.542) | 0.52 (0.294;0.745) | 0.01 (-0.014;0.035) | 0.558 (0.55;0.566) | 0.721 (0.525;0.917) | 0.049 (0.023;0.075) |
| Reynolds Risk | absent | 0.662 (0.658;0.665) | 0.958 (0.922;0.994) | -0.01 (-0.024;0.005) | 0.68 (0.676;0.683) | 0.957 (0.925;0.988) | -0.003 (-0.016;0.011) |
| | present | 0.528 (0.52;0.536) | 0.56 (0.35;0.77) | 0.011 (-0.014;0.035) | 0.552 (0.543;0.56) | 0.754 (0.547;0.961) | 0.049 (0.023;0.075) |
| SCORE CHD | absent | 0.669 (0.665;0.672) | 0.958 (0.928;0.989) | -0.01 (-0.024;0.003) | 0.689 (0.686;0.692) | 0.966 (0.939;0.992) | -0.004 (-0.016;0.009) |
| | present | 0.525 (0.518;0.533) | 0.574 (0.422;0.725) | 0.012 (-0.012;0.036) | 0.552 (0.545;0.56) | 0.749 (0.628;0.87) | 0.051 (0.025;0.076) |
| SCORE CVD | absent | 0.671 (0.667;0.674) | 0.955 (0.927;0.984) | -0.009 (-0.022;0.005) | 0.694 (0.69;0.697) | 0.964 (0.94;0.988) | -0.002 (-0.015;0.01) |
| | present | 0.522 (0.515;0.529) | 0.553 (0.394;0.713) | 0.012 (-0.013;0.036) | 0.552 (0.545;0.56) | 0.765 (0.646;0.885) | 0.05 (0.024;0.076) |
| UKPDS 56 | absent | 0.64 (0.635;0.644) | 0.964 (0.917;1.011) | -0.011 (-0.025;0.004) | 0.649 (0.645;0.652) | 0.941 (0.906;0.976) | -0.003 (-0.017;0.011) |
| | present | 0.533 (0.525;0.541) | 0.647 (0.364;0.929) | 0.01 (-0.014;0.035) | 0.549 (0.54;0.558) | 0.768 (0.511;1.025) | 0.049 (0.023;0.075) |
| UKPDS 68 C-HF | absent | 0.643 (0.64;0.647) | 0.934 (0.898;0.97) | -0.004 (-0.018;0.009) | 0.675 (0.672;0.679) | 0.949 (0.921;0.978) | 0.004 (-0.009;0.017) |
| | present | 0.507 (0.499;0.514) | 0.361 (-0.138;0.859) | 0.012 (-0.012;0.036) | 0.542 (0.534;0.55) | 0.883 (0.676;1.09) | 0.049 (0.023;0.074) |
| UKPDS 68 Stroke | absent | 0.643 (0.64;0.647) | 0.934 (0.898;0.97) | -0.004 (-0.018;0.009) | 0.675 (0.672;0.679) | 0.949 (0.921;0.978) | 0.004 (-0.009;0.017) |
| | present | 0.507 (0.499;0.514) | 0.361 (-0.138;0.859) | 0.012 (-0.012;0.036) | 0.542 (0.534;0.55) | 0.883 (0.676;1.09) | 0.049 (0.023;0.074) |
| UKPDS 82 C-HF | absent | 0.632 (0.627;0.637) | 0.91 (0.861;0.958) | -0.006 (-0.019;0.008) | 0.663 (0.659;0.667) | 0.943 (0.9;0.986) | 0.002 (-0.011;0.015) |
| | present | 0.504 (0.496;0.511) | 0.303 (-0.267;0.874) | 0.012 (-0.012;0.037) | 0.539 (0.531;0.547) | 0.874 (0.589;1.158) | 0.049 (0.023;0.074) |
| UKPDS 82 CHD | absent | 0.618 (0.613;0.623) | 0.958 (0.917;0.999) | -0.006 (-0.02;0.008) | 0.641 (0.637;0.646) | 0.98 (0.951;1.009) | 0.002 (-0.011;0.014) |
| | present | 0.54 (0.533;0.548) | 1.043 (0.782;1.305) | 0.016 (-0.009;0.04) | 0.5 (0.492;0.508) | 0.056 (-2.083;2.194) | 0.052 (0.027;0.078) |

**Appendix Figure 16 Discrimination (c-statistic) for all included risk scores against CVD outcome for the imputed dataset stratified by gender (men vs women), statins (statin naïve vs statin users), CVD history at the baseline (absent vs present), and age subgroups.**

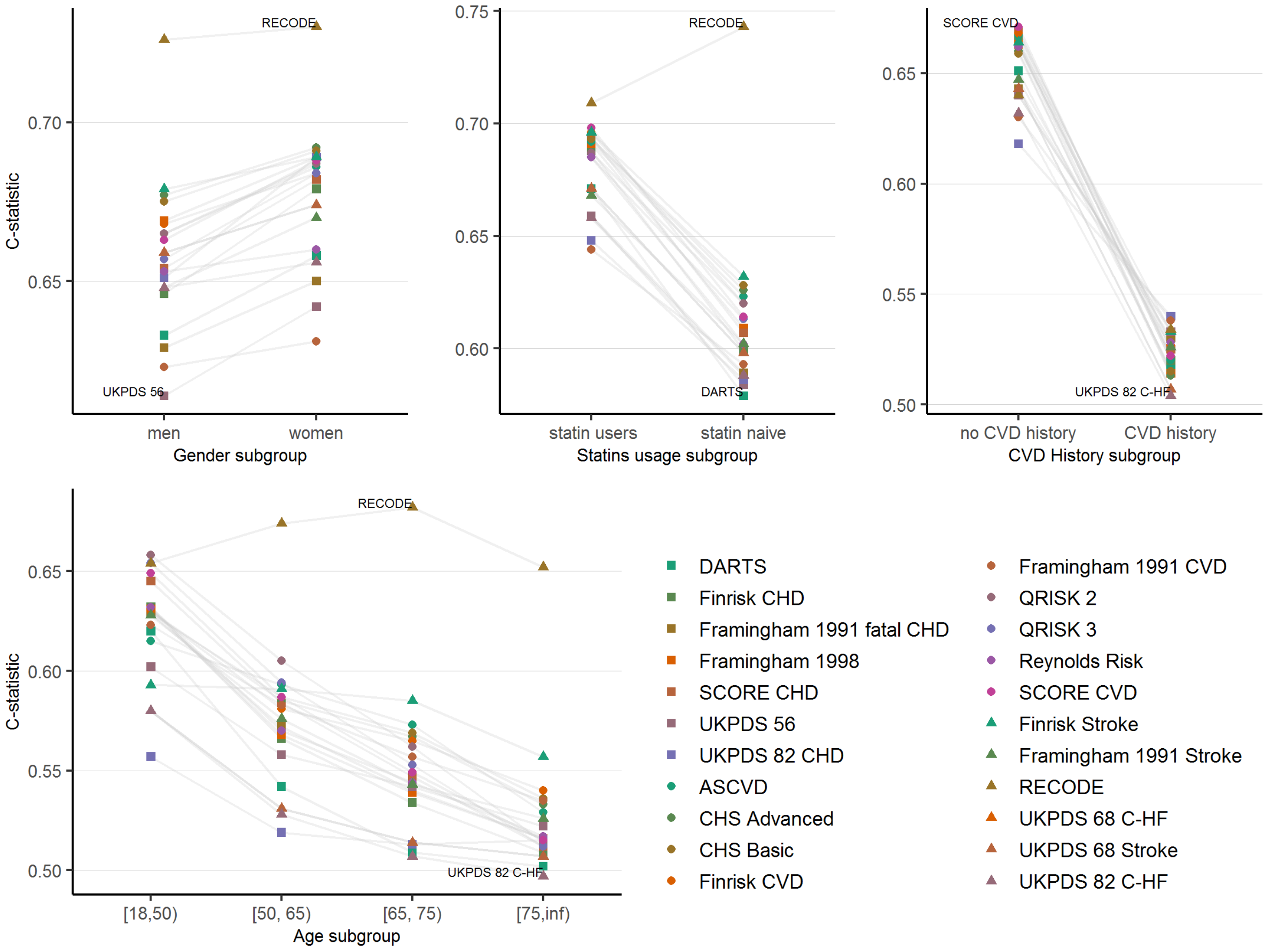

**Appendix Figure 17 Discrimination (c-statistic) for all included risk scores against CVD + AF + HF outcome for the imputed dataset stratified by gender (men vs women), statins (statin naïve vs statin users), CVD history at the baseline (absent vs present), and age subgroups.**

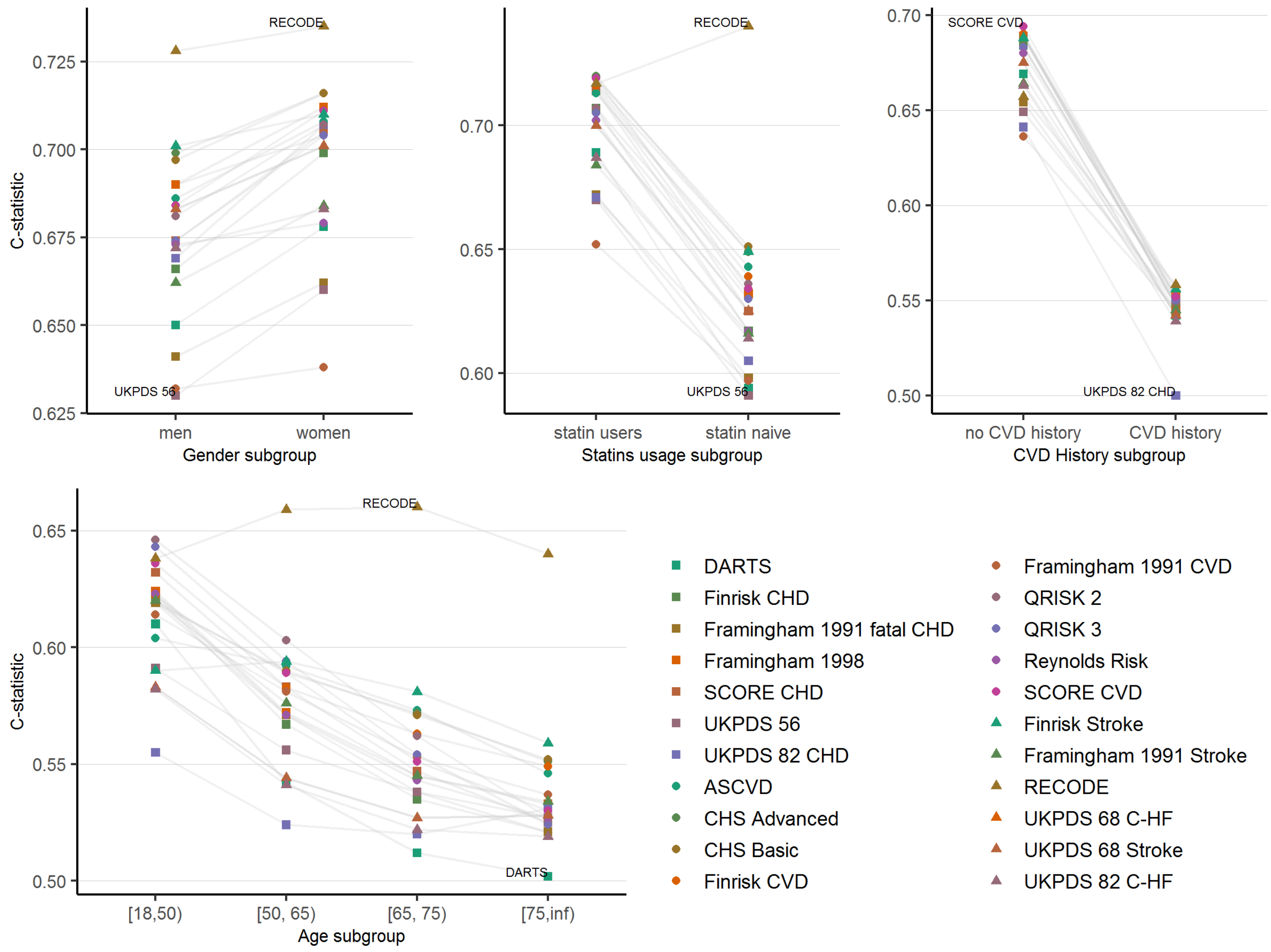

**Appendix Figure 18 Flow diagram of selected articles**

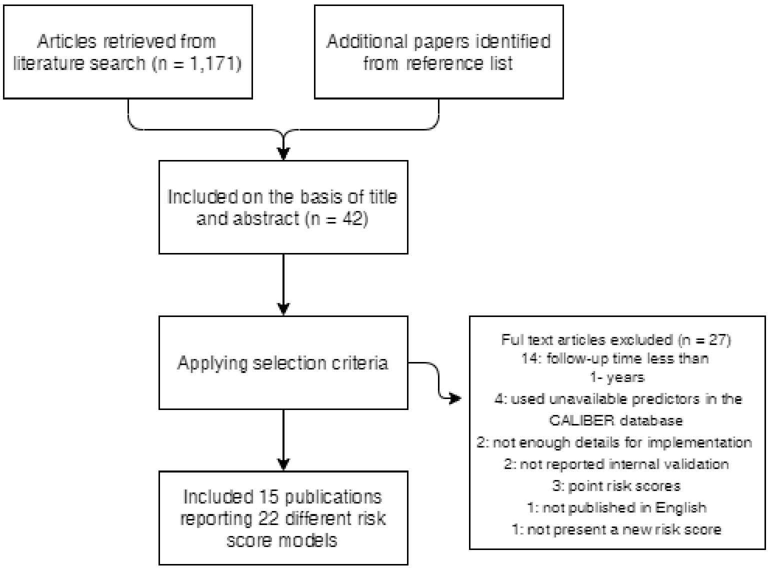
